# Natural history parameters for enteric pathogens to inform modeling studies of diarrhea among children in low-resource settings: Results from the MAL-ED longitudinal birth cohort

**DOI:** 10.1101/2025.05.05.25327027

**Authors:** Avnika B. Amin, Maria Garcia Quesada, Jie Liu, Ritesh Sivakumar, Shane Conyers, Benjamin A. Lopman, Eric Houpt, James A. Platts-Mills, Elizabeth T. Rogawski McQuade

## Abstract

**Background:** Policy decisions may rely on mathematical modelling to predict intervention impacts. Information for key model parameters is limited for most enteric pathogens. To support informed modeling, we characterized incidence, severity, and duration for adenovirus 40/41, astrovirus, *Campylobacter jejuni* and *C. coli, Cryptosporidium*, norovirus GII, rotavirus, sapovirus, *Shigella*, heat-stable enterotoxin-producing enterotoxigenic *Escherichia coli* (ST-ETEC), and typical enteropathogenic *E. coli* (tEPEC) in a longitudinal birth cohort.

**Methods:** We analyzed stool specimens from MAL-ED, a multisite birth cohort with active surveillance of children in South America, southeast Asia, and sub-Saharan Africa. We defined unique infections using longitudinal test results, attributed etiologies to diarrheal episodes, calculated infection rates and disease progression probabilities, and characterized age-based trends.

**Results:** Most pathogens had peak infection rates around 7 to 9 months of age, with incidence gradually decreasing in the second year of life. In contrast, *Cryptosporidium* and ST-ETEC incidence plateaued after 9 months of age instead of declining. Shigella incidence continually increased in the first two years of life. The likelihood of developing diarrhea increased with age for *Shigella* but decreased with age for adenovirus 40/41, *Campylobacter jejuni* and *C. coli*, and tEPEC. The likelihood of attributable diarrhea becoming severe decreased with age.

**Conclusions:** The age at peak infection burden and peak disease burden were not necessarily the same for a given pathogen. Each pathogen evaluated had its own distinct age trends. These results could support informed modelling of impacts of interventions for specific enteric pathogens, particularly in low-resource settings.

## Background

Diarrheal disease is a leading cause of death among children less than five years of age, causing approximately 500,000 deaths annually.(1,2) Rotavirus is the foremost etiology of these deaths despite the availability of safe, effective rotavirus vaccines.(1,2) Other pathogens like *Shigella*, sapovirus, adenovirus types 40 and 41, norovirus, *Campylobacter, Cryptosporidium*, and enterotoxigenic *Escherichia coli* (ETEC) are increasingly recognized as important contributors to under-5 diarrheal disease burden.(3–6) Additional pathogen-specific interventions, like in-development enteric vaccines, and broadly preventive environmental interventions are crucial for curtailing diarrheal disease burden. But having interventions is not enough; interventions must be implemented at scale to achieve measurable impact.

Decision-makers increasingly rely on mathematical models to choose when and how to introduce interventions.(7) With the many competing priorities faced by decision-makers, a convincing investment case requires compelling evidence about an intervention’s projected population-level effects and cost-effectiveness.(8,9) Mathematical models and simulations can provide useful information on these metrics to support decision-making and policies for interventions.(7,10,11) But models require accurate and specific knowledge for the pathogen(s) of interest, including the rate of transmission in the population and the likelihood and duration of different infection and disease states, to accurately predict the impacts of either targeted or broad interventions.(7,12)

The available evidence on key model parameters is limited for most enteric pathogens. As a result, modelers for all of the other enteric pathogens often must adapt information from different data sources of varying quality and comparability that are usually not intended for modelling work.(13) These data sources may suffer heavily from underreporting or other biases, which may not have been relevant for the original research question but heavily impact a model’s accuracy.(14) Longitudinal studies can provide rich information. However, the few with publicly available raw data have study design complexities that make it difficult to derive the desired parameters without completely understanding the primary data collection activities.

Parameterization challenges force modelers to rely on model-based estimation of pathogen characteristics and sensitivity analyses (i.e., where model parameters are varied to evaluate how results may change based on assumptions about parameter values).(12,15) Such uncertainties propagate through model results, affecting interpretation and confidence in decisions about preventive measures. These uncertainties, along with the challenges of communicating them, make it difficult to understand an intervention’s potential.(7)

To support informed modeling of the epidemiology and control of enteric pathogens, we aimed to characterize key parameters using data from a multisite, longitudinal birth cohort study. We describe the top ten contributors to diarrheal disease during the first two years of life: adenovirus 40/41, astrovirus, *Campylobacter jejuni* and *C. coli, Cryptosporidium*, norovirus GII, rotavirus, sapovirus, *Shigella*, heat-stable enterotoxin-producing *Escherichia coli* (ST-ETEC), and typical enteropathogenic *E. coli* (tEPEC). These results will support the accurate parameterization of enteric pathogen models and facilitate assessments of interventions to reduce diarrheal disease, particularly in lower-resource settings.

## Methods

### Study design and participants

We used data from the Etiology, Risk Factors, and Interactions of Enteric Infections and Malnutrition and the Consequences for Child Health and Development (MAL-ED) birth cohort study, which has been previously described.(16) Briefly, children were enrolled from the community within 17 days of birth if their mother was aged ≥16 years, their family intended to stay in the study area for ≥6 months post-enrollment, they were from a singleton pregnancy, they had no other siblings already enrolled, their birthweight or enrollment weight was ≥1500g, they did not have congenital disease or severe neonatal disease. All eight sites (Dhaka, Bangladesh; Vellore, India; Bhaktapur, Nepal; Naushero Feroze, Pakistan; Venda, South Africa; Haydom, Tanzania; Fortaleza, Brazil; and Loreto, Peru) received ethics approval from the appropriate governmental and institutional ethics review boards. Written informed consent was obtained from each child’s parent or guardian.

### Stool sample collection and testing

Stool samples were collected for each diarrheal episode and for monthly non-diarrheal surveillance. Diarrhea was defined as ≥3 loose stools with 24 hours or one stool with visible blood. If >48 hours passed between the occurrence of study-defined diarrhea, they were considered distinct episodes. The number of loose stools per 24 hours, the presence of dehydration and fever, and the duration of vomiting were recorded for each diarrheal episode. Diarrheal severity was calculated using this information as previously described,(17) with a score >6 considered severe.

Stool samples were initially analyzed using a previously described standard protocol.(17,18) Later, available stools from children with complete follow-up through 24 months of age were re-analyzed using customized TaqMan Array Cards (Thermo Fisher, Carlsbad, CA, USA) to detect 29 enteropathogens.(5) Assay validation, nucleic acid extraction, quantitative PCR setup, and quality control have been previously described.(5,19,20)

### Diarrhea etiology

We restricted analyses to the stool samples retested by TaqMan Array Cards and identified pathogen-attributable diarrhea with a previously used algorithm.(21) This algorithm assigns attribution using three steps, with steps evaluated in order from strongest to weakest attribution criteria. The first step requires a pathogen-attributable fraction for the episode (AFe) >0.5 and a tenfold increase in the pathogen quantity from the prior specimen collected in the preceding 60 days. The AFe calculation has been described elsewhere.(5) If no pathogens are attributed in the first step, the second step requires a non-zero AFe and a tenfold increase in pathogen quantity. Finally, if no pathogens have yet been attributed, the third step only requires AFe >0.5. This algorithm considered all tested pathogens in the original study, not only the ten pathogens of interest.

### Analytic datasets

We created separate analytic datasets for each pathogen of interest: adenovirus 40/41, astrovirus, *Campylobacter jejuni* and *C. coli, Cryptosporidium*, norovirus GII, rotavirus, sapovirus, *Shigella*, ST-ETEC (STh or STp), and tEPEC (*bfpA*). Samples missing a test result for the pathogen of interest were excluded from the relevant pathogen-specific dataset. For rotavirus, children in countries that had already introduced rotavirus vaccines (Brazil, Peru, and South Africa) were excluded. For children in the remaining countries, specimens from the 28 days after a rotavirus vaccination were excluded from rotavirus datasets.

We next identified PCR detections likely to represent an active infection, as low-quantity detections may not reflect an established infection, based on the cycle threshold (Ct). Consistent with prior work, we defined positive detections of active infections (i.e., positive detections) as those with Ct ≤30.(22) Pathogens likely to be associated with diarrhea even at lower quantities used slightly higher cutoffs (adenovirus 40/41, Ct ≤30.424, norovirus GII, Ct ≤30.357, rotavirus, Ct ≤32.638; *Shigella*, Ct ≤30.507). Pathogens attributed as the underlying etiology of diarrhea were automatically considered a positive detection regardless of Ct value. We then defined unique infections in two ways to reflect different assumptions. In the first definition (the crude definition), each positive detection was considered a separate infection. If an asymptomatic and symptomatic positive were within seven days of each other, we considered them part of the same infection.

The second definition (the consecutive detections definition) leveraged the longitudinal testing. Two detections were considered consecutive if the child aged by one month or less between detections and there were no negative results in between. All pathogen-attributable diarrheal episodes were considered distinct infections. Detections in the five days preceding the collection of a pathogen-attributable diarrheal stool were considered incubation period shedding, while consecutive detections occurring after the diarrheal stool’s collection were considered post-diarrheal shedding. Incubation and post-diarrheal shedding detections were grouped with the pathogen-attributable diarrheal episode into a single infection.

We then grouped the remaining detections based on consecutive positive detections, where each grouping reflected a single sub-clinical infection (i.e. asymptomatic infections and infections detected during a diarrheal episode that were not considered the underlying etiology). Given the monthly surveillance, we assumed that infections started either at the midpoint between the first positive and the prior negative or, if no test result was available for the month before the first positive, 15 days before the first positive. Similarly, we assumed infections ended either at the midpoint between the last positive and the next negative or, if no test result was available for the month after the last positive, 15 days after the last positive.

### Statistical analysis

We calculated the infection rate and exact 95% confidence intervals (CIs) for each pathogen of interest, comparing results between the two infection definitions. Children contributed 30.5 child-days (i.e., a child-month) if they had at least one tested stool sample in the analytic dataset for a given month of life, minus the duration of infections in each month. When using the crude definition, sub-clinical infections were assumed to last one week. Symptomatic infections were assumed to last for the duration of symptoms plus two additional days, which allowed for the protocol-specified >48 symptom-free hours between diarrheal episodes.

To characterize pathogen natural histories, we calculated the proportion of infections that resulted in severe (as previously defined by score >6) or any pathogen-attributable diarrhea and exact 95% CIs. We also characterized the duration of pathogen-attributable diarrhea and, for the consecutive detections definition, the presumed duration of infection. Infection rates were stratified by 3-month age bands, while proportions and durations were stratified by 6-month age bands to limit uncertainty in the estimates. When using the consecutive detections definition, we further stratified results by infection history (zero or ≥1 prior infection). Finally, we calculated the median age at first infection among the children who had an infection.

To evaluate potential heterogeneity by country, we evaluated age at first infection and rates of infection, pathogen-attributable diarrhea, and severe pathogen-attributable diarrhea by country. Infection rates and age at first infection were calculated as previously described. Time at risk for diarrhea rates was calculated by subtracting the duration of disease plus two days from contributed child-time. This accommodated the protocol-required 48 symptom-free hours for separate diarrheal episodes. Rates were not estimated by age due to low power.

To evaluate the robustness of our results, we conducted three sensitivity analyses. First, we repeated all described analyses using datasets where only AFe was used to determine diarrhea etiology. Using AFe>0.5 is a more established method for etiologic detection, particularly for this dataset.(5) To determine if using 6-month bands for age obscured more nuanced trends, we next repeated the main analyses that stratified by 6-month bands using 3-month bands instead. Finally, we used a cycle threshold <35 to define an infection and repeated the main analyses of age-stratified infection rates, the proportion of infections resulting in attributable diarrhea, age at first infection, and infection rates by site. Code to reproduce the main statistical analysis and sensitivity analyses is available at [to be added].

## Results

The 1,715 children included in non-rotavirus analyses contributed 36,923.6 child-months, with 1,119 children contributing 24,765.0 child-months to rotavirus analyses. Total child-months varied by pathogen and the infection definition, with fewer child-months at risk for infection and fewer unique infections when using the consecutive detections definition (Supplemental Table 1). The number of pathogen-attributable or severe pathogen-attributable diarrheal episodes did not differ between the two infection definitions.

### Infection incidence rates

Rotavirus generally had a lower infection rate across all age groups than the other pathogens (Figure 1, Supplemental Table 2). Most pathogens’ infection rates peaked between 7 and 12 months of age, with incidence gradually decreasing in the second year of life. *Cryptosporidium* and ST-ETEC incidence peaked around the same time, though incidence plateaued afterwards instead of declining. Shigella incidence increased over the first two years of life. Both infection definitions yielded similar age trends, and rates were similar or sometimes higher using the crude infection definition.

**Figure 1.**
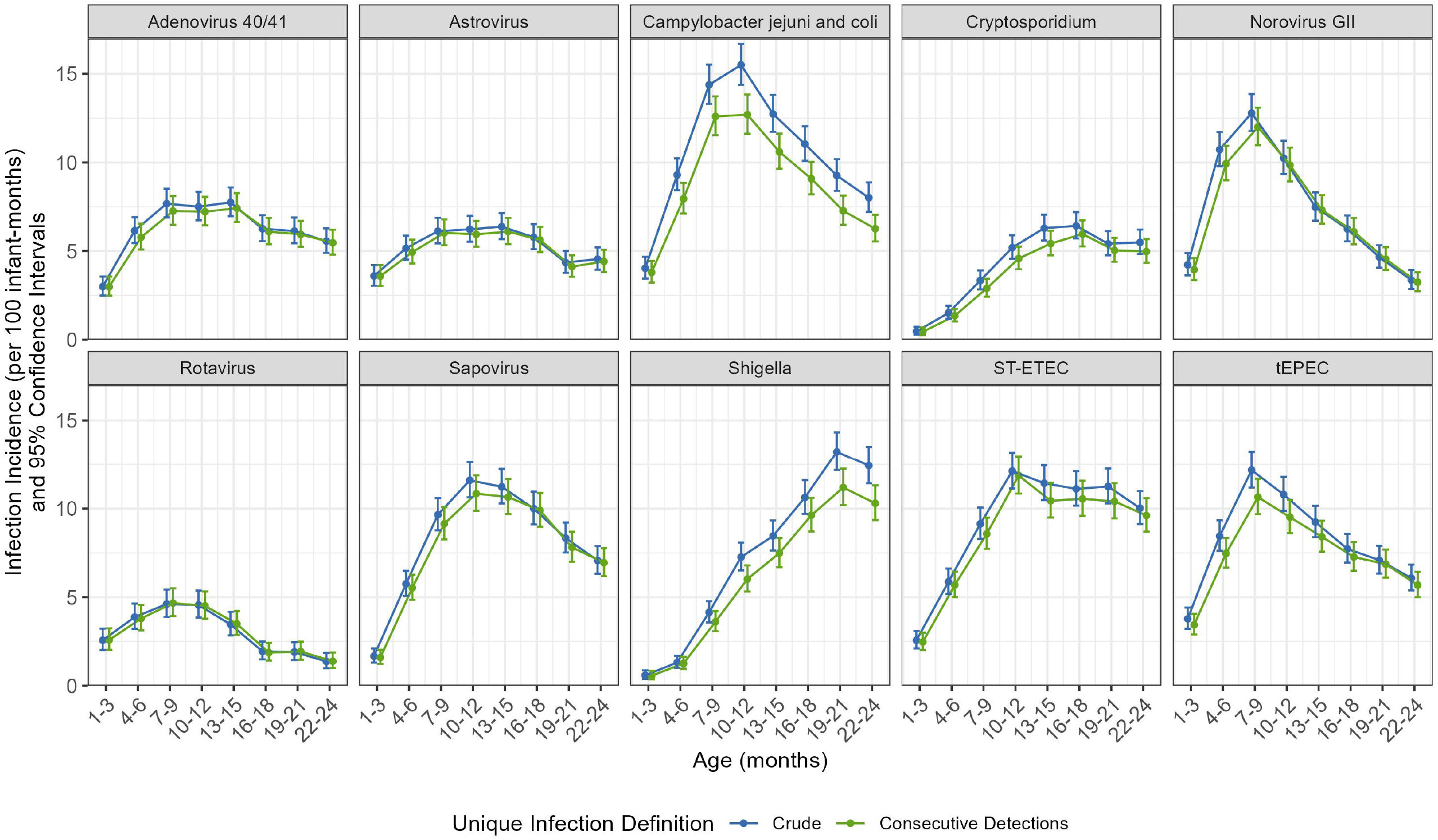
Infection incidence rates per 100 child-months and 95% confidence intervals using algorithm attribution of etiology and different infection definitions (crude or consecutive). I-bars indicate exact 95% confidence intervals. Algorithm attribution considered both the attributable fraction per episode and longitudinal changes in the quantity of pathogen detected. With the crude infection definition, all positives were unique infections unless positive asymptomatic and symptomatic detections were separated by <7 days. The consecutive detections definition considered both consecutive positive detections and time between consecutive positives.

The described age trends persisted after stratifying by infection histories (Supplemental Table 3). For most pathogens, children with at least one prior infection by a given pathogen had similar infection rates as children with no infection history. However, children previously infected with adenovirus 40/41, *Campylobacter jejuni* and *C. coli, Shigella*, or ST-ETEC had higher infection rates than infection-naïve children. 95% CIs for rates in the first few months of life were wider for children with a prior infection when compared to the narrow 95% CIs for infection-naïve children.

First infections by *Campylobacter jejuni* and *C. coli*, norovirus GII, and rotavirus predominantly occurred in the first year of life across all study sites (Figure 2, Supplemental Table 4). *Shigella* and *Cryptosporidium* tended to have first infections in the second year of life, while the other pathogens had more varied median ages at first infection, though first infections were still generally in the first year of life. These findings were consistent regardless of infection definition.

**Figure 2.**
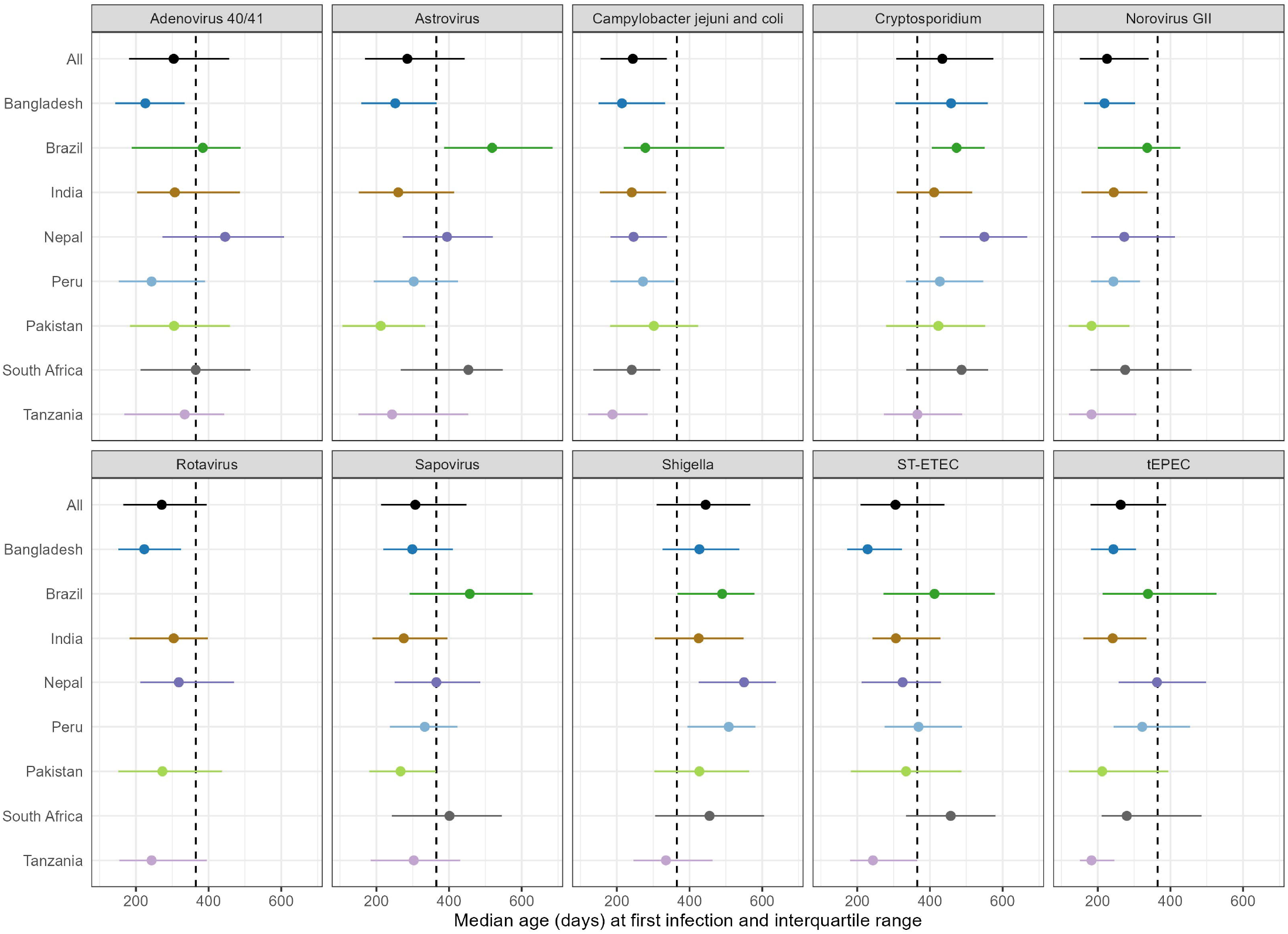
Median and interquartile range for age at first infection, regardless of symptomology, by country using algorithm attribution and the consecutive detections definition. Points denote medians, horizontal lines indicate interquartile ranges, and vertical dashed line indicates one year of age. Brazil, Peru, and South Africa had already introduced rotavirus vaccines and were not included in rotavirus analyses. Algorithm attribution considered both the attributable fraction per episode and longitudinal changes in the quantity of pathogen detected. The consecutive detections definition considered both consecutive positive detections and time between consecutive positives.

### Infection severity and duration

The proportion of infections resulting in pathogen-attributable diarrhea was larger for rotavirus than for any other pathogen across nearly all age groups (Figure 3, Supplemental Table 5). Age did not appear strongly linked to the proportion of astrovirus, *Cryptosporidium, Shigella*, or ST-ETEC infections resulting in pathogen-attributable diarrhea. The probability of infections resulting in diarrhea decreased with age for adenovirus 40/41, *Campylobacter jejuni* and *C. coli*, and tEPEC. Norovirus GII, rotavirus, and sapovirus had the highest attributable diarrhea proportions between 7 and 18 months of age. These trends were apparent with both infection definitions, although the actual proportions were higher with the consecutive detections definition.

**Figure 3.**
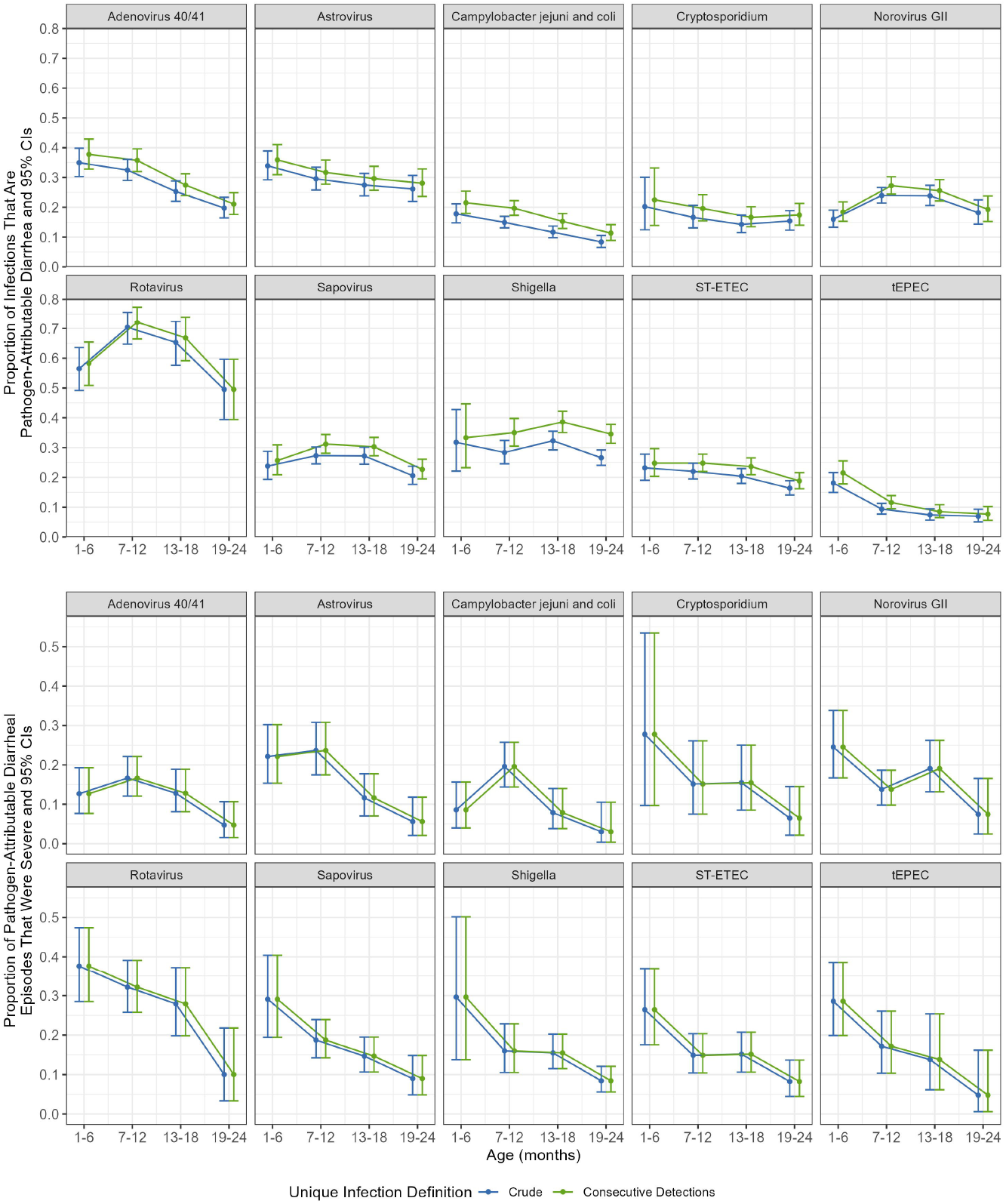
Proportion of infections that resulted in diarrhea (top) and proportion of pathogen-attributable diarrheal episodes that were severe (bottom) using algorithm attribution of etiology and different infection definitions (crude or consecutive). I-bars indicate exact 95% confidence intervals. Algorithm-based attribution considered both the attributable fraction per episode and longitudinal changes in the quantity of pathogen detected. With the crude infection definition, all positives were unique infections unless positive asymptomatic and symptomatic detections were separated by <7 days. The consecutive detections definition considered both consecutive positive detections and time between consecutive positives.

When further stratifying by infection history, pathogen-specific age trends were generally similar to unstratified trends (Supplemental Table 6). Exceptions included diarrhea progression probabilities for norovirus GII and sapovirus among children with prior infections, where the probability of diarrhea appeared to decrease with age. Prior infections were associated with higher diarrhea probabilities for adenovirus 40/41, astrovirus, sapovirus, and ST-ETEC across most age groups. However, infection history did not appear to affect diarrhea probabilities for other pathogens.

For most pathogens, the proportion of pathogen-attributable diarrheal episodes that were severe decreased with age (Figure 3, Supplemental Table 7). Adenovirus 40/41 and *Campylobacter jejuni* and *C. coli* were exceptions to this trend, with peak proportions between 7 and 12 months of age. Trends did not differ by infection definition, as both definitions resulted in the same numbers of pathogen-attributable and severe pathogen-attributable diarrhea episodes (Supplemental Table 1). When further stratifying by infection history, pathogen-specific age trends were generally similar to the unstratified trends regardless of infection history (Supplemental Table 8). However, 95% CIs for children in the second year of life with no infection history and for children in the first year of life with prior infections were quite wide due to limited episodes of severe diarrhea.

The median duration of infection was roughly a month for most pathogens regardless of the age at infection (Supplemental Table 9). Infection history did not appear to affect these trends, though duration tended to skew shorter among children with prior infections (Supplemental Table 10).

The duration of diarrheal symptoms was generally consistent across age groups regardless of the attributed pathogen (Figure 4, Supplemental Table 11). The variation in diarrhea duration slightly decreased as age increased. Children with prior infections tended to have a shorter duration of diarrhea than infection-naïve children (Supplemental Table 12).

**Figure 4.**
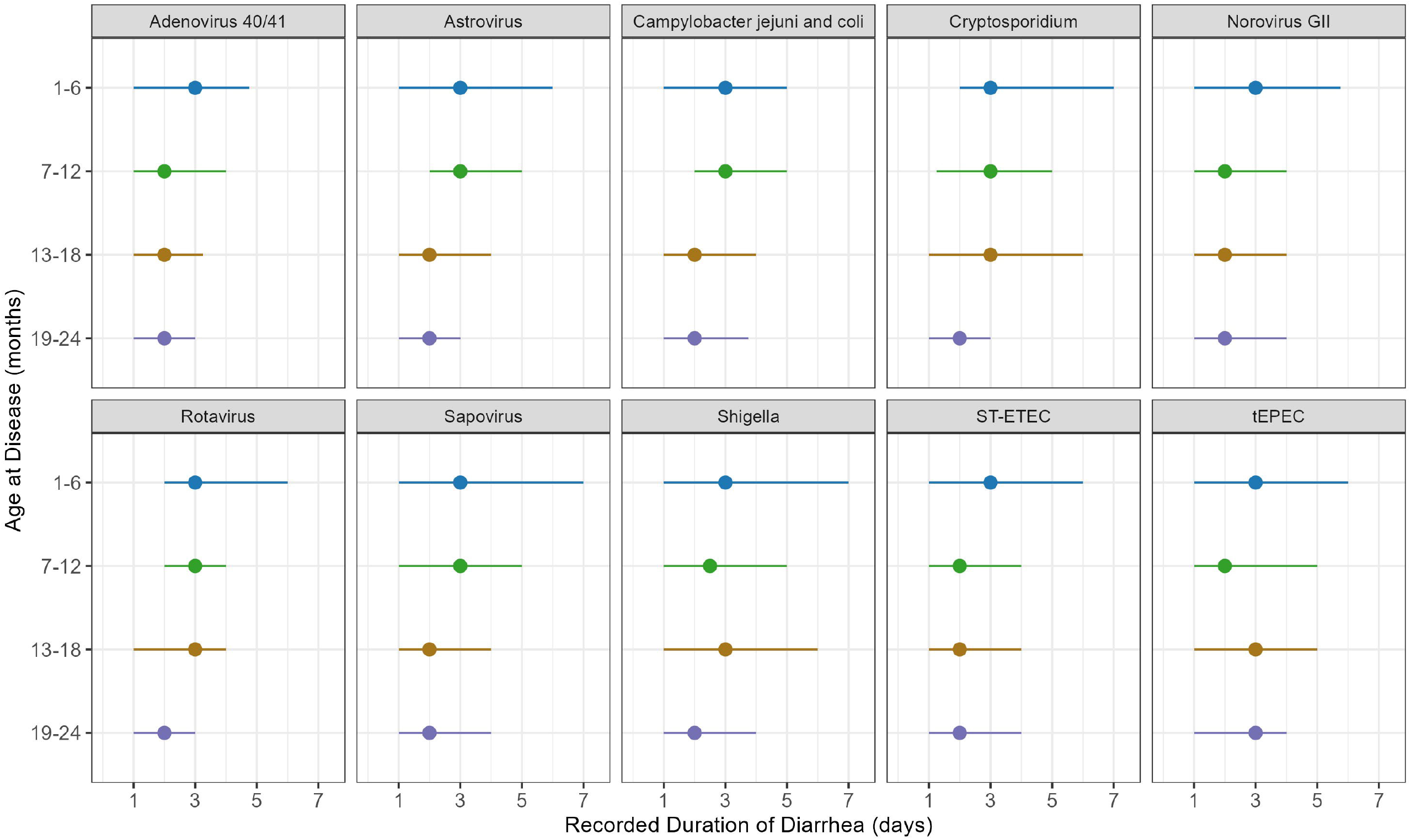
Median and interquartile range for duration of pathogen-attributable diarrhea using algorithm attribution of etiology. The points denote medians, while the lines indicate the interquartile ranges. Algorithm-based attribution considered both the attributable fraction per episode and longitudinal changes in the quantity of pathogen detected. The consecutive detections definition considered both consecutive positive detections and time between consecutive positives. Because the recorded start and end of symptoms are consistent across infection definitions, they are not separately reported.

### Country-specific incidence rates and age at first infection

Infection rates varied by study site for each pathogen except rotavirus, which had similar infection rates across sites (Figure 5, Supplemental Table 13). Bangladesh and Peru usually had the highest infection rates. Infection rates were usually slightly higher when using the crude infection definition instead of the consecutive detections definition.

**Figure 5.**
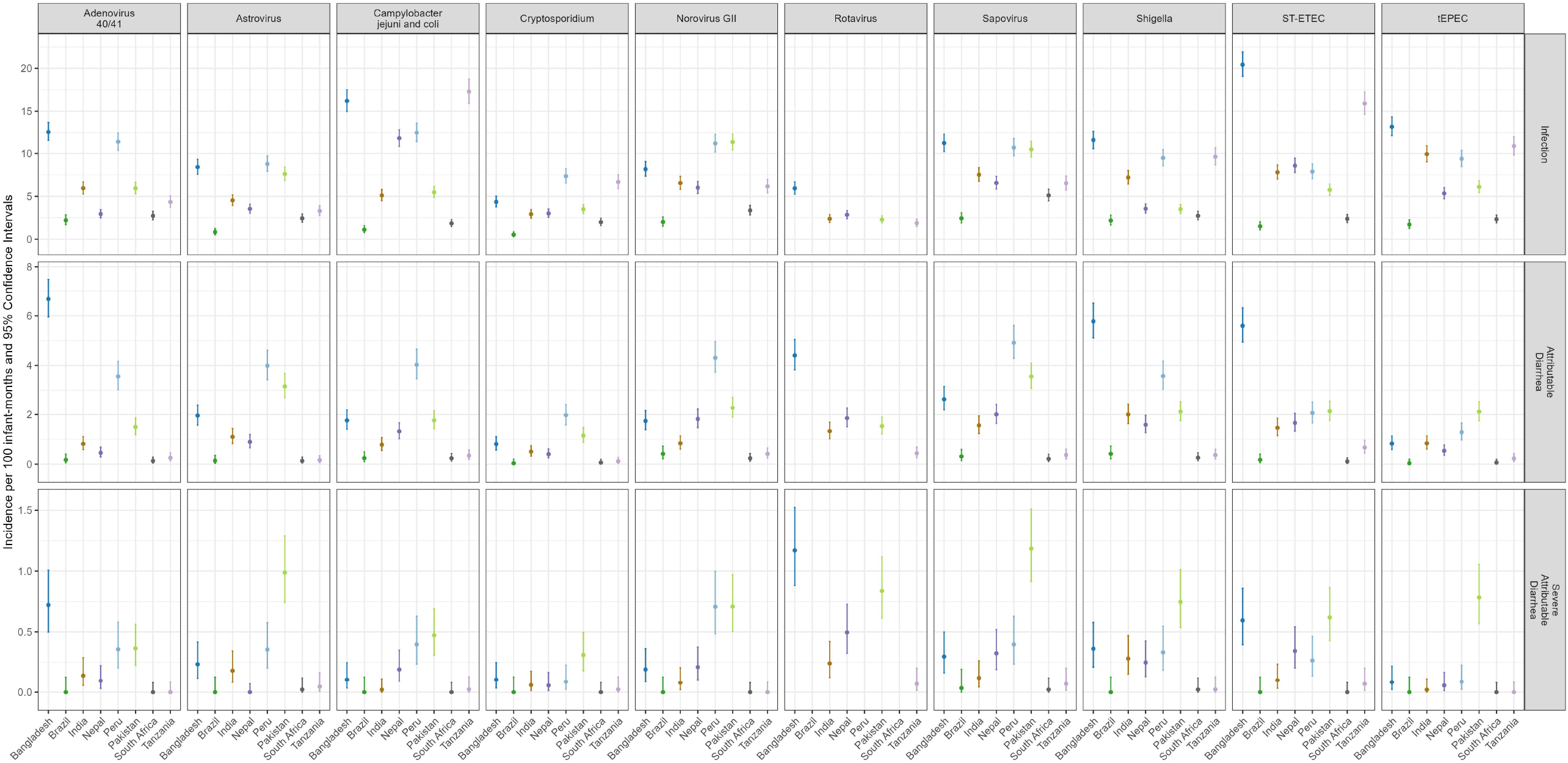
Incidence rates of infection (top), pathogen-attributable diarrhea (middle), and severe (score >6) pathogen-attributable diarrhea (bottom) per 100 child-months and 95% confidence intervals for each study site using algorithm attribution of etiology and the consecutive detections definition for unique infections. I-bars indicate exact 95% confidence intervals. Algorithm attribution considered both the attributable fraction per episode and longitudinal changes in the quantity of pathogen detected. The consecutive detections definition considered both consecutive positive detections and time between consecutive positives.

Rates of pathogen-attributable diarrhea (Figure 5, Supplemental Table 14) and severe pathogen-attributable diarrhea (Figure 5, Supplemental Table 15) varied substantially by site for all pathogens except *Cryptosporidium* and tEPEC. Bangladesh, Peru, and Pakistan generally had the highest rates of both pathogen-attributable diarrhea and severe pathogen-attributable diarrhea across different pathogens. Infection definition did not impact either of the observed rates.

### Sensitivity analyses

In the sensitivity analysis using the more established AFe >0.5 to attribute etiology, the number of pathogen-attributable and severe pathogen-attributable diarrheal episodes substantially decreased for all pathogens except rotavirus and *Shigella* (Supplemental Table 1). In general, the described trends for infection incidence, age at first infection, proportion of infections that resulted in diarrhea, proportion of pathogen-attributable diarrheal episodes that were severe, infection duration, and pathogen-attributable diarrhea duration did not substantially change when using only AFe to attribute etiology (Supplemental Tables 2-12). However certain characteristics, like the proportions of infections resulting in diarrhea, were attenuated in magnitude since each pathogen was attributed to fewer episodes of diarrhea. A similar phenomenon occurred for trends in site-specific rates (Supplemental Tables 13-15).

Using 3-month age bands instead of 6-month age bands did not substantially change the observed trends for pathogen-attributable diarrhea and severe pathogen-attributable diarrhea proportions, duration of infection, or duration of pathogen-attributable diarrhea (Supplemental Tables 16-19). While some additional nuance may have been gained, the 95% confidence intervals for the proportions were wider and the interquartile ranges for the durations were broader. This increased imprecision made it difficult to determine if the more refined patterns reflected underlying age trends or were a statistical artifact.

Finally, when defining a positive detection with the less stringent Ct <30, infection rates were higher, and the two infection definitions yielded less similar values (Supplemental Table 20). Proportions of infections resulting in diarrhea decreased in magnitude (Supplemental Table 21). However, age trends for rates and proportions did not substantially change. Country-specific infection rates were higher and inter-country heterogeneity increased (Supplemental Table 22). Finally, the median age at first infection generally decreased slightly both overall and by country (Supplemental Table 23), consistent with the correspondingly higher infection rate.

## Discussion

By characterizing infection rates, median age at first infection, probabilities of progressing to different disease states, and durations of pathogen-attributable diarrhea and infections, this study provides direct estimates of key parameters for enteric pathogen modeling. We demonstrate distinct age trends for pathogen natural histories and characterize heterogeneity by country. Our analyses were robust to different definitions of unique infection episodes, different methods for pathogen attribution, and different age groupings. This supports the generalizability of the results for use in modeling and simulation for pediatric populations in low- and middle-income countries. Some parameters can also be combined to yield other values of interest. For example, if interested in the proportion of infections that are severe, the two progression probabilities can be combined.

Our results can support a variety of modeling efforts, for example to understand pathogen epidemiology, to predict the impacts of climate changes on pathogen burden, and to evaluate the impacts of interventions that specifically or broadly target enteric pathogens. For example, most of the examined pathogens do not yet have a vaccine. Because vaccines are expected to reduce disease burden rather than impact infection risks, accurate models can help project the averted disease burden from vaccine introduction and guide target vaccine profiles required to optimize vaccine impact and cost-effectiveness.(23) Broader interventions, like household-based water filtration, may have varying impacts against different enteric pathogens,(24) so models can project their impacts based on pathogen distributions. They can also evaluate how disease dynamics, like pathogen strain diversity or seasonality, may change in response to an intervention when coupled with other context-specific knowledge.(25) When researchers can use accurate parameters in their models, their results facilitate informed decision-making and set reasonable expectations for an intervention’s impact.

While we conducted this study with the goal of informing enteric pathogen modeling, our findings can also inform relevant timing for interventions. For example, interventions that aim to prevent *Campylobacter jejuni* and *C. coli*, norovirus GII, sapovirus, and tEPEC may have the best impact when delivered early in life, as this is when their infection incidences and probabilities of severe, pathogen-attributable diarrhea are highest. However, given the varied timing of peak incidences and attributable diarrhea probabilities in the first two years of life, non-specific or multi-pathogen preventive efforts may affect different pathogens based on when they affect an infant. Models may inform the features of intervention delivery, like the number and timing of vaccine doses or the timing of booster doses, based on programmatic goals for reducing disease and pathogen burden.(26,27)

This study is subject to several limitations. The results are most applicable to the first two years of life for children in low and middle income countries, so they may not generalize well to other settings. MAL-ED was not powered for heavily-stratified analyses, so we could not further stratify analyses by other relevant factors or adjust analyses for potential confounders. Underlying differences in susceptibility may explain the counterintuitive higher infection incidence among previously-infected children relative to infection-naïve children.(28,29) We could not define independent infections with complete certainty since infection surveillance occurred monthly. We attempted to mitigate this limitation by reporting the two extreme assumptions of the crude and consecutive detections definitions for unique infections. Finally, diarrheal etiology may have been misclassified, which would most affect the accuracy of the probabilities of pathogen-attributable disease states and the duration of pathogen-attributable diarrhea. We attempted to mitigate this issue by examining different methods of pathogen attribution, which revealed similar age-based trends.

## Conclusions

In summary, we describe age-based trends for infection incidence and pathogen natural histories to support accurate parameterization of enteric pathogen models. Our findings provide crucial information about pathogen epidemiology for the top ten contributors to global diarrhea disease among young children. Many of these pathogens have under-described natural histories. We identify pathogen-specific similarities and differences in the timing of peak infection incidence and disease conversion, which may help inform decisions about preventive measures.

## Data Availability

Individual-level data from the original MAL-ED study will not be made publicly available. However, pseudo-data and code for statistical analysis will be available on GitHub.

## Declarations

### Ethics approval and consent to participate

Ethical approval for MAL-ED was obtained from the institutional review boards at the University of Virginia School of Medicine (Charlottesville, VA) (14595), and at each of the participating research sites: Ethical Review Committee, International Centre for Diarrhoeal Disease Research, Bangladesh (Bangladesh); Committee for Ethics in Research, Universidade Federal do Ceara, and National Ethical Research Committee, Health Ministry, Council of National Health (Brazil); Institutional Review Board, Christian Medical College, Vellore, and Health Ministry Screening Committee, Indian Council of Medical Research (India); Institutional Review Board, Institute of Medicine, Tribhuvan University, Ethical Review Board, Nepal Health Research Council, and Institutional Review Board, Walter Reed Army Institute of Research (Nepal); Institutional Review Board, Johns Hopkins University, and PRISMA Ethics Committee; Health Ministry, Loreto (Peru); Ethical Review Committee, Aga Khan University (Pakistan); Health, Safety and Research Ethics Committee, University of Venda, and Department of Health and Social Development, Limpopo Provincial Government (South Africa); and Medical Research Coordinating Committee, National Institute for Medical Research, and Chief Medical Officer, Ministry of Health and Social Welfare (Tanzania). The secondary analysis was conducted after MAL-ED’s completion with de-identified data and was deemed not to meet the definition of human subjects research.

### Clinical trial number

Not applicable

### Consent for publication

Not applicable

### Competing interests

The authors declare they have no competing interests.

### Funding

The original Etiology, Risk Factors and Interactions of Enteric Infections and Malnutrition and the Consequences for Child Health and Development Project (MAL-ED) was supported by the Bill & Melinda Gates Foundation (OPP1131125), the Foundation for the NIH, the National Institutes of Health (AI114888), and the Fogarty International Center.

### Authors’ contributions

ABA led data analysis and writing of the manuscript. MGQ, RS, SC, BAL, and ETRM contributed to data analysis. JL, EH, JAP-M, and ETRM contributed to the data acquisition and design of the original study. ABA and ETRM conceived the secondary data analysis project. MGQ, JL, RS, SC, BAL, EH, JAP-M, and ETRM contributed to interpretation of the results and critical review of the manuscript.

## Acknowledgements

We thank MAL-ED staff and participants for their important contributions to this project.

## Notes

### Competing Interest Statement

The authors have declared no competing interest.

### Author Declarations

Ethical approval for MAL-ED was obtained from the institutional review boards at the University of Virginia School of Medicine (Charlottesville, VA) (14595), and at each of the participating research sites: Ethical Review Committee, International Centre for Diarrhoeal Disease Research, Bangladesh (Bangladesh); Committee for Ethics in Research, Universidade Federal do Ceara, and National Ethical Research Committee, Health Ministry, Council of National Health (Brazil); Institutional Review Board, Christian Medical College, Vellore, and Health Ministry Screening Committee, Indian Council of Medical Research (India); Institutional Review Board, Institute of Medicine, Tribhuvan University, Ethical Review Board, Nepal Heal th Research Council, and Institutional Review Board, Walter Reed Army Institute of Research (Nepal); Institutional Review Board, Johns Hopkins University, and PRISMA Ethics Committee; Health Ministry, Loreto (Peru); Ethical Review Committee, Aga Khan University (Pakistan); Health, Safety and Research Ethics Committee, University of Venda, and Department of Heal th and Social Development, Limpopo Provincial Government (South Africa); and Medical Research Coordinating Committee, National Insti tute for Medical Research, and Chief Medical Officer, Ministry of Health and Social Welfare (Tanzania). The secondary analysis was conducted after MAL-ED's completion with de-identified data and was deemed not to meet the definition of human subjects research.

