## Supplemental Material for "Natural history parameters for enteric pathogens to inform modeling studies of diarrhea among children in low-resource settings: Results from the MAL-ED longitudinal birth cohort"

**Formatted for Lancet Global Health**

Manuscript text: 3,455/3,500 words (main text only)

### Supplemental Material

Supplemental Table 1. Infant-months, infections, pathogen-attributable diarrheal episodes, and severe pathogen-attributable diarrheal episodes included in analysis with different approaches to etiologic attribution (algorithm or AFe) and infection definition (crude or consecutive).

Supplemental Table 2. Infection incidence rates per 100 infant-months and exact 95% confidence intervals (95% CIs) by 3-month age bands using different approaches to etiologic attribution (algorithm or AFe) and infection definition (crude or consecutive detections).

Supplemental Table 3. Infection incidence rates per 100 infant-months and exact 95% confidence intervals (95% CIs) by 3-month age bands and prior infection history using different approaches to etiologic attribution (algorithm or AFe) and the consecutive detections definition for unique infections.

Supplemental Table 4. Median and interquartile range for age (in days) at first infection, regardless of symptomology, by country.

Supplemental Table 5. Proportion of infections that were pathogen-attributable diarrhea and exact 95% confidence intervals (95% CIs) by 6-month age bands using different approaches to etiologic attribution (algorithm or AFe) and infection definition (crude or consecutive detections).

Supplemental Table 6. Proportion of infections that were pathogen-attributable diarrhea and exact 95% confidence intervals (95% CIs) by 6-month age bands and infection history using different approaches to etiologic attribution (algorithm or AFe) and the consecutive detections definition for unique infections.

Supplemental Table 7. Proportion of pathogen-attributable diarrheal episodes that were severe (score >6) and exact 95% confidence intervals (95% CIs) by 6-month age bands using different approaches to etiologic attribution (algorithm or AFe).

Supplemental Table 8. Proportion of pathogen-attributable diarrheal episodes that were severe (score >6) and exact 95% confidence intervals (95% CIs) by 6-month age bands and infection history using different approaches to etiologic attribution (algorithm or AFe) and the consecutive detections definition for unique infections.

Supplemental Table 9. Median and interquartile range (in days) for duration of infection by 6-month age bands using different approaches to etiologic attribution (algorithm or AFe) and the consecutive detections definition for unique infections.

Supplemental Table 10. Median and interquartile range (in days) for duration of infection by 6-month age bands and infection history using different approaches to etiologic attribution (algorithm or AFe) and the consecutive detections definition for unique infections.

Supplemental Table 11. Median and interquartile range (in days) for duration of pathogen-attributable diarrhea by 6-month age bands using different approaches to etiologic attribution (algorithm or AFe).

Supplemental Table 12. Median and interquartile range (in days) for duration of pathogen-attributable diarrhea by 6-month age bands and infection history using different approaches to etiologic attribution (algorithm or AFe).

Supplemental Table 13. Infection incidence rates per 100 infant-months and exact 95% confidence intervals (95% CIs) by country using different approaches to etiologic attribution (algorithm or AFe) and infection definition (crude or consecutive detections).

Supplemental Table 14. Pathogen-attributable diarrhea incidence rates per 100 infant-months and exact 95% confidence intervals (95% CIs) by country using different approaches to etiologic attribution (algorithm or AFe).

Supplemental Table 15. Severe (score >6) pathogen-attributable diarrhea incidence rates per 100 infant-months and exact 95% confidence intervals (95% CIs) by country using different approaches to etiologic attribution (algorithm or AFe).

Supplemental Table 16. Proportion of infections that were pathogen-attributable diarrhea and proportion of pathogen-attributable diarrheal episodes that were severe (score >6) and exact 95% confidence intervals (95% CIs) by 3-month age bands using algorithm attribution and different approaches to infection definition (crude or consecutive detections).

Supplemental Table 17. Proportion of infections that were pathogen-attributable diarrhea and proportion of pathogen-attributable diarrheal episodes that were severe (score >6) and exact 95% confidence intervals (95% CIs) by 3-month age bands and infection history using algorithm attribution and the consecutive detections definition for unique infections.

Supplemental Table 18. Median and interquartile range for duration of infection by 3-month age bands (unstratified or stratified by infection history) using algorithm attribution and the consecutive detections definition for unique infections.

Supplemental Table 19. Median and interquartile range for duration of pathogen-attributable diarrhea by 3-month age bands (unstratified or stratified by infection history) using algorithm attribution.

Supplemental Table 20. Infection incidence rates per 100 infant-months and exact 95% confidence intervals (95% CIs) by 3-month age bands using Ct <30 to define a positive detection, algorithm attribution, and different infection definitions (crude or consecutive detections).

Supplemental Table 21. Proportion of infections that were pathogen-attributable diarrhea and exact 95% confidence intervals (95% CIs) by 6-month age bands using Ct <30 to define a positive detection, algorithm attribution and different infection definitions (crude or consecutive detections).

Supplemental Table 22. Infection incidence rates per 100 infant-months and exact 95% confidence intervals (95% CIs) by country using Ct <30 to define a positive detection, algorithm attribution and different infection definitions (crude or consecutive detections).

Supplemental Table 23. Median and interquartile range for age at first infection, regardless of symptomology, by country using Ct <30 to define a positive detection.

**Supplemental Table 1. Infant-months, infections, pathogen-attributable diarrheal episodes, and severe pathogen-attributable diarrheal episodes included in analysis with different approaches to etiologic attribution (algorithm or AFe) and infection definition (crude or consecutive).** With pathogen-attributable fraction (AFe) attribution, a pathogen was attributed to a diarrheal episode with AFe >0.5. Algorithm attribution considered both AFe and longitudinal changes in the quantity of pathogen detected. With the crude infection definition, all positives were unique infections unless positive asymptomatic and symptomatic detections were separated by <7 days. The consecutive detections definition considered both consecutive positive detections and time between consecutive positives.

| Pathogen (number of infants) | Outcome | Number of outcomes (infant-months) | | | |
|---|---|---|---|---|---|
| | | Algorithm attribution | | AFe attribution | |
| | | Crude | Consecutive detections | Crude | Consecutive detections |
| Adenovirus 40/41 (n=1,715) | Infections | 2,293 (36,463.0) | 2,114 (34,975.5) | 2,168 (36,481.0) | 2,137 (34,954.6) |
| | Pathogen-Attributable Diarrheal Episodes | 640 (36,788.2) | 640 (36,788.2) | 411 (36,826.7) | 411 (36,826.7) |
| | Severe Pathogen-Attributable Diarrheal Episodes | 82 (36,871.5) | 82 (36,871.5) | 61 (36,878.7) | 61 (36,878.7) |
| Astrovirus (n=1,715) | Infections | 1,931 (36,501.1) | 1,802 (35,276.1) | 1,831 (36,523.1) | 1,714 (35,344.8) |
| | Pathogen-Attributable Diarrheal Episodes | 562 (36,770.4) | 562 (36,770.4) | 308 (36,822.7) | 308 (36,822.7) |
| | Severe Pathogen-Attributable Diarrheal Episodes | 93 (36,842.2) | 93 (36,842.2) | 46 (36,862.2) | 46 (36,862.2) |
| Campylobacter jejuni and coli (n=1,715) | Infections | 3,831 (36,158.3) | 2,938 (33,524.7) | 3,755 (36,171.4) | 2,854 (33,576.1) |
| | Pathogen-Attributable Diarrheal Episodes | 502 (36,813.2) | 502 (36,813.2) | 162 (36,878.2) | 162 (36,878.2) |
| | Severe Pathogen-Attributable Diarrheal Episodes | 61 (36,886.0) | 61 (36,886.0) | 12 (36,903.2) | 12 (36,903.2) |
| Cryptosporidium (n=1,715) | Infections | 1,575 (36,586.5) | 1,364 (35,513.8) | 1,559 (36,588.0) | 1,348 (35,524.2) |
| | Pathogen-Attributable Diarrheal Episodes | 245 (36,848.2) | 245 (36,848.2) | 119 (36,871.2) | 119 (36,871.2) |
| | Severe Pathogen-Attributable Diarrheal Episodes | 33 (36,882.2) | 33 (36,882.2) | 16 (36,888.3) | 16 (36,888.3) |
| Norovirus GII (n=1,715) | Infections | 2,728 (36,424.5) | 2,449 (34,602.1) | 2,682 (36,433.6) | 2,405 (34,636.7) |
| | Pathogen-Attributable Diarrheal Episodes | 579 (36,847.2) | 579 (36,847.2) | 296 (36,903.0) | 296 (36,903.0) |
| | Severe Pathogen-Attributable Diarrheal Episodes | 95 (36,923.6) | 95 (36,923.6) | 40 (36,940.9) | 40 (36,940.9) |
| Rotavirus (n=1,119) | Infections | 751 (24,661.9) | 734 (24,228.0) | 761 (24,662.0) | 750 (24,224.6) |
| | Pathogen-Attributable Diarrheal Episodes | 472 (24,716.8) | 472 (24,716.8) | 486 (24,716.1) | 486 (24,716.1) |

| | | | | | |
|---|---|---|---|---|---|
| | Severe Pathogen-Attributable Diarrheal Episodes | 142 (24,765.0) | 142 (24,765.0) | 142 (24,765.8) | 142 (24,765.8) |
| Sapovirus (n=1,715) | Infections | 2,993 (36,366.4) | 2,683 (34,372.3) | 2,941 (36,381.0) | 2,654 (34,406.4) |
| | Pathogen-Attributable Diarrheal Episodes | 757 (36,806.2) | 757 (36,806.2) | 542 (36,852.9) | 542 (36,852.9) |
| | Severe Pathogen-Attributable Diarrheal Episodes | 125 (36,903.7) | 125 (36,903.7) | 67 (36,925.3) | 67 (36,925.3) |
| Shigella (n=1,715) | Infections | 2,656 (36,420.1) | 2,139 (34,632.4) | 2,609 (36,431.1) | 2,149 (34,664.8) |
| | Pathogen-Attributable Diarrheal Episodes | 770 (36,791.1) | 770 (36,791.1) | 755 (36,795.9) | 755 (36,795.9) |
| | Severe Pathogen-Attributable Diarrheal Episodes | 102 (36,897.6) | 102 (36,897.6) | 90 (36,901.2) | 90 (36,901.2) |
| ST-ETEC (n=1,715) | Infections | 3,352 (36,233.6) | 2,952 (33,949.7) | 3,285 (36,245.3) | 2,912 (33,993.1) |
| | Pathogen-Attributable Diarrheal Episodes | 671 (36,761.0) | 671 (36,761.0) | 452 (36,802.7) | 452 (36,802.7) |
| | Severe Pathogen-Attributable Diarrheal Episodes | 100 (36,846.9) | 100 (36,846.9) | 62 (36,860.6) | 62 (36,860.6) |
| tEPEC (n=1,715) | Infections | 2,984 (36,339.7) | 2,542 (34,257.5) | 2,959 (36,349.7) | 2,510 (34,275.2) |
| | Pathogen-Attributable Diarrheal Episodes | 297 (36,868.2) | 297 (36,868.5) | 31 (36,925.7) | 31 (36,925.7) |
| | Severe Pathogen-Attributable Diarrheal Episodes | 55 (36,906.9) | 55 (36,906.9) | 8 (36,929.4) | 8 (36,929.4) |

**Supplemental Table 2. Infection incidence rates per 100 infant-months and exact 95% confidence intervals (95% CIs) by 3-month age bands using different approaches to etiologic attribution (algorithm or AFe) and infection definition (crude or consecutive detections).** With pathogen-attributable fraction (AFe) attribution, a pathogen was attributed to a diarrheal episode with AFe >0.5. Algorithm attribution considered both AFe and longitudinal changes in the quantity of pathogen detected. With the crude infection definition, all positives were unique infections unless positive asymptomatic and symptomatic detections were separated by <7 days. The consecutive detections definition considered both consecutive positive detections and time between consecutive positives. Italicized column headers correspond to Figure 1 data.

| Pathogen | Age (months) | Algorithm attribution | | | | AFe attribution | | | |
|---|---|---|---|---|---|---|---|---|---|
| | | Crude | | *Consecutive detections* | | Crude | | Consecutive detections | |
| | | Infections (Infant-months) | Incidence (95% CI) | *Infections (Infant-months)* | *Incidence (95% CI)* | Infections (Infant-months) | Incidence (95% CI) | Infections (Infant-months) | Incidence (95% CI) |
| Adenovirus 40/41 | 1-3 | 127 (4,245.1) | 2.99 (2.49,3.56) | 124 (4,153.5) | 2.99 (2.48,3.56) | 115 (4,246.7) | 2.71 (2.24,3.25) | 124 (4,143.2) | 2.99 (2.49,3.57) |
| | 4-6 | 279 (4,535.8) | 6.15 (5.45,6.92) | 252 (4,360.3) | 5.78 (5.09,6.54) | 261 (4,539.6) | 5.75 (5.07,6.49) | 255 (4,352.5) | 5.86 (5.16,6.62) |
| | 7-9 | 355 (4,623.3) | 7.68 (6.90,8.52) | 320 (4,407.4) | 7.26 (6.49,8.10) | 323 (4,628.1) | 6.98 (6.24,7.78) | 326 (4,407.6) | 7.40 (6.62,8.24) |
| | 10-12 | 347 (4,624.8) | 7.50 (6.73,8.34) | 318 (4,402.5) | 7.22 (6.45,8.06) | 329 (4,627.1) | 7.11 (6.36,7.92) | 322 (4,402.8) | 7.31 (6.54,8.16) |
| | 13-15 | 359 (4,631.9) | 7.75 (6.97,8.60) | 327 (4,406.5) | 7.42 (6.64,8.27) | 346 (4,633.7) | 7.47 (6.70,8.30) | 332 (4,403.2) | 7.54 (6.75,8.40) |
| | 16-18 | 289 (4,616.8) | 6.26 (5.56,7.02) | 270 (4,427.9) | 6.10 (5.39,6.87) | 276 (4,618.7) | 5.98 (5.29,6.72) | 272 (4,427.8) | 6.14 (5.43,6.92) |
| | 19-21 | 281 (4,581.6) | 6.13 (5.44,6.89) | 261 (4,391.9) | 5.94 (5.24,6.71) | 272 (4,582.8) | 5.94 (5.25,6.68) | 263 (4,391.9) | 5.99 (5.29,6.76) |
| | 22-24 | 256 (4,603.5) | 5.56 (4.90,6.29) | 242 (4,425.5) | 5.47 (4.80,6.20) | 246 (4,604.3) | 5.34 (4.70,6.05) | 243 (4,425.5) | 5.49 (4.82,6.23) |
| Astrovirus | 1-3 | 152 (4,232.3) | 3.59 (3.04,4.21) | 148 (4,128.2) | 3.59 (3.03,4.21) | 133 (4,238.1) | 3.14 (2.63,3.72) | 128 (4,145.9) | 3.09 (2.58,3.67) |
| | 4-6 | 234 (4,535.7) | 5.16 (4.52,5.86) | 217 (4,394.3) | 4.94 (4.30,5.64) | 223 (4,538.7) | 4.91 (4.29,5.60) | 208 (4,399.5) | 4.73 (4.11,5.42) |
| | 7-9 | 284 (4,639.6) | 6.12 (5.43,6.88) | 269 (4,464.0) | 6.03 (5.33,6.79) | 269 (4,644.7) | 5.79 (5.12,6.53) | 255 (4,474.7) | 5.70 (5.02,6.44) |
| | 10-12 | 288 (4,624.5) | 6.23 (5.53,6.99) | 264 (4,441.1) | 5.94 (5.25,6.71) | 279 (4,626.5) | 6.03 (5.34,6.78) | 260 (4,445.3) | 5.85 (5.16,6.60) |
| | 13-15 | 297 (4,655.7) | 6.38 (5.67,7.15) | 273 (4,474.8) | 6.10 (5.40,6.87) | 283 (4,658.1) | 6.08 (5.39,6.83) | 261 (4,483.8) | 5.82 (5.14,6.57) |
| | 16-18 | 267 (4,619.8) | 5.78 (5.11,6.52) | 250 (4,445.1) | 5.62 (4.95,6.37) | 256 (4,621.1) | 5.54 (4.88,6.26) | 239 (4,453.4) | 5.37 (4.71,6.09) |
| | 19-21 | 200 (4,594.2) | 4.35 (3.77,5.00) | 184 (4,466.3) | 4.12 (3.55,4.76) | 195 (4,594.5) | 4.24 (3.67,4.88) | 181 (4,468.8) | 4.05 (3.48,4.69) |
| | 22-24 | 209 (4,599.2) | 4.54 (3.95,5.20) | 197 (4,462.2) | 4.41 (3.82,5.08) | 193 (4,601.4) | 4.19 (3.62,4.83) | 182 (4,473.3) | 4.07 (3.50,4.70) |
| *Campylobacter jejuni* and *coli* | 1-3 | 171 (4,244.4) | 4.03 (3.45,4.68) | 155 (4,084.0) | 3.80 (3.22,4.44) | 165 (4,245.5) | 3.89 (3.32,4.53) | 149 (4,083.5) | 3.65 (3.09,4.28) |

| | | | | | | | | | |
|---|---|---|---|---|---|---|---|---|---|
| | 4-6 | 419<br>(4,505.2) | 9.30<br>(8.43,10.24) | 333<br>(4,188.7) | 7.95<br>(7.12,8.85) | 409<br>(4,506.7) | 9.08<br>(8.22,10.00) | 319<br>(4,201.1) | 7.59<br>(6.78,8.47) |
| | 7-9 | 658<br>(4,574.8) | 14.38<br>(13.31,15.53) | 517<br>(4,103.4) | 12.60<br>(11.54,13.73) | 645<br>(4,580.2) | 14.08<br>(13.02,15.21) | 503<br>(4,110.5) | 12.24<br>(11.19,13.35) |
| | 10-12 | 706<br>(4,553.0) | 15.51<br>(14.38,16.69) | 520<br>(4,096.4) | 12.69<br>(11.63,13.83) | 693<br>(4,555.4) | 15.21<br>(14.10,16.39) | 507<br>(4,105.4) | 12.35<br>(11.30,13.47) |
| | 13-15 | 586<br>(4,599.6) | 12.74<br>(11.73,13.81) | 446<br>(4,206.7) | 10.60<br>(9.64,11.63) | 574<br>(4,600.8) | 12.48<br>(11.48,13.54) | 433<br>(4,213.6) | 10.28<br>(9.33,11.29) |
| | 16-18 | 504<br>(4,567.1) | 11.04<br>(10.09,12.04) | 385<br>(4,237.7) | 9.09<br>(8.20,10.04) | 492<br>(4,568.0) | 10.77<br>(9.84,11.77) | 372<br>(4,248.6) | 8.76<br>(7.89,9.69) |
| | 19-21 | 421<br>(4,545.4) | 9.26<br>(8.40,10.19) | 309<br>(4,248.9) | 7.27<br>(6.48,8.13) | 417<br>(4,545.8) | 9.17<br>(8.31,10.10) | 305<br>(4,250.6) | 7.18<br>(6.39,8.03) |
| | 22-24 | 366<br>(4,568.8) | 8.01<br>(7.21,8.88) | 273<br>(4,359.0) | 6.26<br>(5.54,7.05) | 360<br>(4,568.8) | 7.88<br>(7.09,8.74) | 266<br>(4,362.8) | 6.10<br>(5.39,6.88) |
| *Cryptosporidium* | 1-3 | 20<br>(4,266.5) | 0.47<br>(0.29,0.72) | 19<br>(4,250.3) | 0.45<br>(0.27,0.70) | 20<br>(4,266.4) | 0.47<br>(0.29,0.72) | 19<br>(4,250.3) | 0.45<br>(0.27,0.70) |
| | 4-6 | 69<br>(4,574.0) | 1.51<br>(1.17,1.91) | 61<br>(4,523.1) | 1.35<br>(1.03,1.73) | 69<br>(4,574.6) | 1.51<br>(1.17,1.91) | 61<br>(4,523.1) | 1.35<br>(1.03,1.73) |
| | 7-9 | 156<br>(4,670.8) | 3.34<br>(2.84,3.91) | 132<br>(4,559.3) | 2.90<br>(2.42,3.43) | 156<br>(4,671.1) | 3.34<br>(2.84,3.91) | 133<br>(4,559.3) | 2.92<br>(2.44,3.46) |
| | 10-12 | 241<br>(4,640.0) | 5.19<br>(4.56,5.89) | 205<br>(4,477.3) | 4.58<br>(3.97,5.25) | 239<br>(4,639.6) | 5.15<br>(4.52,5.85) | 203<br>(4,479.1) | 4.53<br>(3.93,5.20) |
| | 13-15 | 293<br>(4,655.7) | 6.29<br>(5.59,7.06) | 242<br>(4,463.9) | 5.42<br>(4.76,6.15) | 288<br>(4,657.0) | 6.18<br>(5.49,6.94) | 238<br>(4,466.0) | 5.33<br>(4.67,6.05) |
| | 16-18 | 296<br>(4,607.7) | 6.42<br>(5.71,7.20) | 263<br>(4,409.1) | 5.96<br>(5.27,6.73) | 292<br>(4,608.3) | 6.34<br>(5.63,7.11) | 258<br>(4,412.1) | 5.85<br>(5.16,6.61) |
| | 19-21 | 248<br>(4,581.7) | 5.41<br>(4.76,6.13) | 222<br>(4,406.7) | 5.04<br>(4.40,5.75) | 245<br>(4,581.6) | 5.35<br>(4.70,6.06) | 218<br>(4,408.7) | 4.94<br>(4.31,5.65) |
| | 22-24 | 252<br>(4,590.3) | 5.49<br>(4.83,6.21) | 220<br>(4,423.9) | 4.97<br>(4.34,5.68) | 250<br>(4,589.4) | 5.45<br>(4.79,6.17) | 218<br>(4,425.5) | 4.93<br>(4.29,5.63) |
| Norovirus GII | 1-3 | 179<br>(4,240.2) | 4.22<br>(3.63,4.89) | 162<br>(4,101.0) | 3.95<br>(3.37,4.61) | 176<br>(4,240.6) | 4.15<br>(3.56,4.81) | 158<br>(4,104.0) | 3.85<br>(3.27,4.50) |
| | 4-6 | 483<br>(4,504.7) | 10.72<br>(9.79,11.72) | 414<br>(4,168.1) | 9.93<br>(9.00,10.94) | 478<br>(4,506.6) | 10.61<br>(9.68,11.60) | 407<br>(4,172.6) | 9.75<br>(8.83,10.75) |
| | 7-9 | 588<br>(4,597.8) | 12.79<br>(11.78,13.87) | 507<br>(4,224.3) | 12.00<br>(10.98,13.09) | 580<br>(4,600.7) | 12.61<br>(11.60,13.68) | 501<br>(4,229.7) | 11.84<br>(10.83,12.93) |
| | 10-12 | 472<br>(4,604.6) | 10.25<br>(9.35,11.22) | 424<br>(4,301.8) | 9.86<br>(8.94,10.84) | 467<br>(4,605.4) | 10.14<br>(9.24,11.10) | 421<br>(4,307.1) | 9.77<br>(8.86,10.75) |
| | 13-15 | 348<br>(4,650.6) | 7.48<br>(6.72,8.31) | 324<br>(4,425.5) | 7.32<br>(6.55,8.16) | 342<br>(4,651.8) | 7.35<br>(6.59,8.17) | 319<br>(4,428.9) | 7.20<br>(6.43,8.04) |
| | 16-18 | 289<br>(4,626.4) | 6.25<br>(5.55,7.01) | 270<br>(4,430.6) | 6.09<br>(5.39,6.87) | 279<br>(4,627.9) | 6.03<br>(5.34,6.78) | 260<br>(4,437.2) | 5.86<br>(5.17,6.62) |
| | 19-21 | 214<br>(4,591.4) | 4.66<br>(4.06,5.33) | 202<br>(4,446.0) | 4.54<br>(3.94,5.22) | 207<br>(4,591.8) | 4.51<br>(3.91,5.17) | 195<br>(4,450.6) | 4.38<br>(3.79,5.04) |
| | 22-24 | 155<br>(4,608.8) | 3.36<br>(2.85,3.94) | 146<br>(4,504.8) | 3.24<br>(2.74,3.81) | 153<br>(4,608.8) | 3.32<br>(2.81,3.89) | 144<br>(4,506.7) | 3.20<br>(2.69,3.76) |

| | | | | | | | | | |
|---|---|---|---|---|---|---|---|---|---|
| Rotavirus | 1-3 | 74 (2,881.2) | 2.57 (2.02,3.22) | 73 (2,828.7) | 2.58 (2.02,3.24) | 69 (2,883.4) | 2.39 (1.86,3.03) | 69 (2,835.0) | 2.43 (1.89,3.08) |
| | 4-6 | 119 (3,071.0) | 3.87 (3.21,4.64) | 114 (3,003.8) | 3.80 (3.13,4.56) | 118 (3,070.9) | 3.84 (3.18,4.60) | 115 (3,003.5) | 3.83 (3.16,4.60) |
| | 7-9 | 144 (3,124.1) | 4.61 (3.89,5.43) | 142 (3,043.9) | 4.67 (3.93,5.50) | 147 (3,124.0) | 4.71 (3.98,5.53) | 146 (3,042.2) | 4.80 (4.05,5.64) |
| | 10-12 | 143 (3,137.5) | 4.56 (3.84,5.37) | 138 (3,061.7) | 4.51 (3.79,5.33) | 144 (3,137.4) | 4.59 (3.87,5.40) | 140 (3,060.8) | 4.57 (3.85,5.40) |
| | 13-15 | 109 (3,151.4) | 3.46 (2.84,4.17) | 108 (3,092.2) | 3.49 (2.87,4.22) | 114 (3,150.5) | 3.62 (2.98,4.35) | 113 (3,090.5) | 3.66 (3.01,4.40) |
| | 16-18 | 61 (3,136.4) | 1.94 (1.49,2.50) | 58 (3,102.5) | 1.87 (1.42,2.42) | 63 (3,136.2) | 2.01 (1.54,2.57) | 61 (3,100.4) | 1.97 (1.50,2.53) |
| | 19-21 | 59 (3,097.0) | 1.91 (1.45,2.46) | 59 (3,058.9) | 1.93 (1.47,2.49) | 61 (3,096.7) | 1.97 (1.51,2.53) | 61 (3,057.7) | 1.99 (1.53,2.56) |
| | 22-24 | 42 (3,063.3) | 1.37 (0.99,1.85) | 42 (3,036.3) | 1.38 (1.00,1.87) | 45 (3,062.9) | 1.47 (1.07,1.97) | 45 (3,034.5) | 1.48 (1.08,1.98) |
| Sapovirus | 1-3 | 71 (4,256.4) | 1.67 (1.30,2.10) | 67 (4,201.8) | 1.59 (1.24,2.03) | 68 (4,259.3) | 1.60 (1.24,2.02) | 64 (4,203.9) | 1.52 (1.17,1.94) |
| | 4-6 | 261 (4,542.7) | 5.75 (5.07,6.49) | 241 (4,362.4) | 5.52 (4.85,6.27) | 257 (4,545.3) | 5.65 (4.98,6.39) | 235 (4,365.2) | 5.38 (4.72,6.12) |
| | 7-9 | 446 (4,620.5) | 9.65 (8.78,10.59) | 396 (4,331.7) | 9.14 (8.26,10.09) | 437 (4,624.2) | 9.45 (8.58,10.38) | 386 (4,336.5) | 8.90 (8.04,9.83) |
| | 10-12 | 532 (4,585.6) | 11.60 (10.64,12.63) | 460 (4,241.7) | 10.84 (9.88,11.88) | 528 (4,587.8) | 11.51 (10.55,12.53) | 462 (4,244.5) | 10.88 (9.91,11.92) |
| | 13-15 | 519 (4,620.4) | 11.23 (10.29,12.24) | 456 (4,281.3) | 10.65 (9.70,11.68) | 510 (4,621.5) | 11.04 (10.10,12.04) | 449 (4,287.1) | 10.47 (9.53,11.49) |
| | 16-18 | 459 (4,588.6) | 10.00 (9.11,10.96) | 423 (4,275.6) | 9.89 (8.97,10.88) | 450 (4,589.8) | 9.80 (8.92,10.75) | 420 (4,282.9) | 9.81 (8.89,10.79) |
| | 19-21 | 381 (4,570.6) | 8.34 (7.52,9.22) | 337 (4,315.1) | 7.81 (7.00,8.69) | 374 (4,571.2) | 8.18 (7.37,9.05) | 338 (4,321.2) | 7.82 (7.01,8.70) |
| | 22-24 | 324 (4,581.5) | 7.07 (6.32,7.89) | 303 (4,362.7) | 6.95 (6.19,7.77) | 317 (4,581.8) | 6.92 (6.18,7.72) | 300 (4,365.0) | 6.87 (6.12,7.70) |
| *Shigella* | 1-3 | 25 (4,267.0) | 0.59 (0.38,0.86) | 24 (4,248.7) | 0.56 (0.36,0.84) | 23 (4,267.4) | 0.54 (0.34,0.81) | 23 (4,250.0) | 0.54 (0.34,0.81) |
| | 4-6 | 60 (4,583.1) | 1.31 (1.00,1.69) | 57 (4,531.7) | 1.26 (0.95,1.63) | 56 (4,583.8) | 1.22 (0.92,1.59) | 54 (4,535.1) | 1.19 (0.89,1.55) |
| | 7-9 | 193 (4,657.4) | 4.14 (3.58,4.77) | 163 (4,511.0) | 3.61 (3.08,4.21) | 191 (4,657.6) | 4.10 (3.54,4.73) | 163 (4,513.5) | 3.61 (3.08,4.21) |
| | 10-12 | 336 (4,627.0) | 7.26 (6.51,8.08) | 265 (4,398.0) | 6.03 (5.32,6.80) | 329 (4,628.9) | 7.11 (6.36,7.92) | 263 (4,403.3) | 5.97 (5.27,6.74) |
| | 13-15 | 392 (4,637.8) | 8.45 (7.64,9.33) | 328 (4,380.2) | 7.49 (6.70,8.34) | 386 (4,639.2) | 8.32 (7.51,9.19) | 328 (4,385.1) | 7.48 (6.69,8.33) |
| | 16-18 | 487 (4,582.9) | 10.63 (9.70,11.61) | 408 (4,240.1) | 9.62 (8.71,10.60) | 476 (4,586.5) | 10.38 (9.47,11.35) | 406 (4,251.9) | 9.55 (8.64,10.52) |
| | 19-21 | 599 (4,528.9) | 13.23 (12.19,14.33) | 461 (4,118.9) | 11.19 (10.19,12.26) | 590 (4,530.5) | 13.02 (11.99,14.12) | 471 (4,126.4) | 11.41 (10.41,12.49) |

| | | | | | | | | | |
|---|---|---|---|---|---|---|---|---|---|
| | 22-24 | 564<br>(4,536.0) | 12.43<br>(11.43,13.50) | 433<br>(4,203.7) | 10.30<br>(9.35,11.32) | 558<br>(4,537.4) | 12.30<br>(11.30,13.36) | 441<br>(4,199.6) | 10.50<br>(9.54,11.53) |
| ST-ETEC | 1-3 | 109<br>(4,250.1) | 2.56<br>(2.11,3.09) | 103<br>(4,168.8) | 2.47<br>(2.02,3.00) | 100<br>(4,252.9) | 2.35<br>(1.91,2.86) | 94<br>(4,174.6) | 2.25<br>(1.82,2.76) |
| | 4-6 | 266<br>(4,535.6) | 5.86<br>(5.18,6.61) | 248<br>(4,360.1) | 5.69<br>(5.00,6.44) | 256<br>(4,539.0) | 5.64<br>(4.97,6.37) | 238<br>(4,366.6) | 5.45<br>(4.78,6.19) |
| | 7-9 | 422<br>(4,617.2) | 9.14<br>(8.29,10.05) | 371<br>(4,328.0) | 8.57<br>(7.72,9.49) | 412<br>(4,618.1) | 8.92<br>(8.08,9.83) | 363<br>(4,333.4) | 8.38<br>(7.54,9.28) |
| | 10-12 | 555<br>(4,577.7) | 12.12<br>(11.14,13.18) | 496<br>(4,179.2) | 11.87<br>(10.85,12.96) | 551<br>(4,578.0) | 12.04<br>(11.05,13.08) | 493<br>(4,184.9) | 11.78<br>(10.76,12.87) |
| | 13-15 | 527<br>(4,608.1) | 11.44<br>(10.48,12.46) | 446<br>(4,270.6) | 10.44<br>(9.50,11.46) | 520<br>(4,609.3) | 11.28<br>(10.33,12.29) | 444<br>(4,274.4) | 10.39<br>(9.44,11.40) |
| | 16-18 | 508<br>(4,571.6) | 11.11<br>(10.17,12.12) | 447<br>(4,237.1) | 10.55<br>(9.59,11.57) | 495<br>(4,572.1) | 10.83<br>(9.89,11.82) | 440<br>(4,245.6) | 10.36<br>(9.42,11.38) |
| | 19-21 | 509<br>(4,525.0) | 11.25<br>(10.29,12.27) | 434<br>(4,170.0) | 10.41<br>(9.45,11.43) | 501<br>(4,526.7) | 11.07<br>(10.12,12.08) | 433<br>(4,175.5) | 10.37<br>(9.42,11.39) |
| | 22-24 | 456<br>(4,548.4) | 10.03<br>(9.13,10.99) | 407<br>(4,235.9) | 9.61<br>(8.70,10.59) | 450<br>(4,549.2) | 9.89<br>(9.00,10.85) | 407<br>(4,238.1) | 9.60<br>(8.69,10.58) |
| tEPEC | 1-3 | 160<br>(4,235.3) | 3.78<br>(3.22,4.41) | 141<br>(4,104.3) | 3.44<br>(2.89,4.05) | 154<br>(4,238.1) | 3.63<br>(3.08,4.26) | 135<br>(4,106.0) | 3.29<br>(2.76,3.89) |
| | 4-6 | 381<br>(4,511.5) | 8.45<br>(7.62,9.34) | 315<br>(4,217.6) | 7.47<br>(6.67,8.34) | 381<br>(4,515.4) | 8.44<br>(7.61,9.33) | 311<br>(4,219.9) | 7.37<br>(6.57,8.24) |
| | 7-9 | 560<br>(4,599.2) | 12.18<br>(11.19,13.23) | 449<br>(4,214.6) | 10.65<br>(9.69,11.69) | 555<br>(4,600.9) | 12.06<br>(11.08,13.11) | 441<br>(4,217.2) | 10.46<br>(9.50,11.48) |
| | 10-12 | 496<br>(4,593.7) | 10.80<br>(9.87,11.79) | 406<br>(4,266.8) | 9.52<br>(8.61,10.49) | 493<br>(4,594.0) | 10.73<br>(9.80,11.72) | 403<br>(4,268.9) | 9.44<br>(8.54,10.41) |
| | 13-15 | 428<br>(4,632.2) | 9.24<br>(8.38,10.16) | 366<br>(4,350.7) | 8.41<br>(7.57,9.32) | 426<br>(4,632.3) | 9.20<br>(8.34,10.11) | 364<br>(4,352.0) | 8.36<br>(7.53,9.27) |
| | 16-18 | 356<br>(4,607.0) | 7.73<br>(6.95,8.57) | 317<br>(4,362.4) | 7.27<br>(6.49,8.11) | 351<br>(4,608.0) | 7.62<br>(6.84,8.46) | 312<br>(4,367.2) | 7.14<br>(6.37,7.98) |
| | 19-21 | 324<br>(4,572.3) | 7.09<br>(6.34,7.90) | 298<br>(4,341.9) | 6.86<br>(6.11,7.69) | 321<br>(4,572.6) | 7.02<br>(6.27,7.83) | 294<br>(4,345.2) | 6.77<br>(6.01,7.59) |
| | 22-24 | 279<br>(4,588.4) | 6.08<br>(5.39,6.84) | 250<br>(4,399.1) | 5.68<br>(5.00,6.43) | 278<br>(4,588.3) | 6.06<br>(5.37,6.81) | 250<br>(4,398.9) | 5.68<br>(5.00,6.43) |

**Supplemental Table 3. Infection incidence rates per 100 infant-months and exact 95% confidence intervals (95% CIs) by 3-month age bands and prior infection history using different approaches to etiologic attribution (algorithm or AFe) and the consecutive detections definition for unique infections.** With pathogen-attributable fraction (AFe) attribution, a pathogen was attributed to a diarrheal episode with AFe >0.5. Algorithm attribution considered both AFe and longitudinal changes in the quantity of pathogen detected.

| Pathogen | Age (months) | Algorithm attribution | | | | AFe attribution | | | |
|---|---|---|---|---|---|---|---|---|---|
| | | No prior infections | | ≥1 prior infection | | No prior infections | | ≥1 prior infection | |
| | | Infections (Infant-months) | Incidence (95% CI) | Infections (Infant-months) | Incidence (95% CI) | Infections (Infant-months) | Incidence (95% CI) | Infections (Infant-months) | Incidence (95% CI) |
| Adenovirus 40/41 | 1-3 | 120 (4,068.9) | 2.95 (2.45,3.53) | 4 (84.6) | 4.73 (1.29,12.10) | 120 (4,069.2) | 2.95 (2.45,3.53) | 4 (74.1) | 5.40 (1.47,13.83) |
| | 4-6 | 214 (3,873.5) | 5.52 (4.81,6.32) | 38 (486.7) | 7.81 (5.52,10.72) | 216 (3,865.7) | 5.59 (4.87,6.38) | 39 (486.8) | 8.01 (5.70,10.95) |
| | 7-9 | 198 (3,368.9) | 5.88 (5.09,6.76) | 122 (1,038.6) | 11.75 (9.76,14.03) | 196 (3,368.3) | 5.82 (5.03,6.69) | 130 (1,039.4) | 12.51 (10.45,14.85) |
| | 10-12 | 182 (2,821.3) | 6.45 (5.55,7.46) | 136 (1,581.2) | 8.60 (7.22,10.17) | 182 (2,821.1) | 6.45 (5.55,7.46) | 140 (1,581.7) | 8.85 (7.45,10.45) |
| | 13-15 | 172 (2,382.0) | 7.22 (6.18,8.38) | 155 (2,024.5) | 7.66 (6.50,8.96) | 172 (2,381.9) | 7.22 (6.18,8.38) | 160 (2,021.3) | 7.92 (6.74,9.24) |
| | 16-18 | 108 (2,001.5) | 5.40 (4.43,6.51) | 162 (2,426.3) | 6.68 (5.69,7.79) | 108 (2,001.5) | 5.40 (4.43,6.51) | 164 (2,426.3) | 6.76 (5.76,7.88) |
| | 19-21 | 87 (1,735.6) | 5.01 (4.01,6.18) | 174 (2,656.3) | 6.55 (5.61,7.60) | 87 (1,735.6) | 5.01 (4.01,6.18) | 176 (2,656.3) | 6.63 (5.68,7.68) |
| | 22-24 | 63 (1,564.8) | 4.03 (3.09,5.15) | 179 (2,860.7) | 6.26 (5.37,7.24) | 63 (1,564.8) | 4.03 (3.09,5.15) | 180 (2,860.7) | 6.29 (5.41,7.28) |
| Astrovirus | 1-3 | 138 (4,030.0) | 3.42 (2.88,4.05) | 10 (98.1) | 10.19 (4.89,18.74) | 123 (4,068.5) | 3.02 (2.51,3.61) | 5 (77.4) | 6.46 (2.10,15.07) |
| | 4-6 | 194 (3,854.1) | 5.03 (4.35,5.79) | 23 (540.2) | 4.26 (2.70,6.39) | 193 (3,899.1) | 4.95 (4.28,5.70) | 15 (500.4) | 3.00 (1.68,4.94) |
| | 7-9 | 204 (3,370.2) | 6.05 (5.25,6.94) | 65 (1,093.8) | 5.94 (4.59,7.57) | 196 (3,428.0) | 5.72 (4.95,6.58) | 59 (1,046.7) | 5.64 (4.29,7.27) |
| | 10-12 | 165 (2,856.3) | 5.78 (4.93,6.73) | 99 (1,584.9) | 6.25 (5.08,7.60) | 167 (2,918.1) | 5.72 (4.89,6.66) | 93 (1,527.2) | 6.09 (4.92,7.46) |
| | 13-15 | 147 (2,426.8) | 6.06 (5.12,7.12) | 126 (2,048.1) | 6.15 (5.12,7.32) | 148 (2,489.2) | 5.95 (5.03,6.98) | 113 (1,994.6) | 5.67 (4.67,6.81) |
| | 16-18 | 96 (2,112.0) | 4.55 (3.68,5.55) | 154 (2,333.1) | 6.60 (5.60,7.73) | 98 (2,165.8) | 4.52 (3.67,5.51) | 141 (2,287.6) | 6.16 (5.19,7.27) |
| | 19-21 | 67 (1,907.8) | 3.51 (2.72,4.46) | 117 (2,558.4) | 4.57 (3.78,5.48) | 72 (1,946.4) | 3.70 (2.89,4.66) | 109 (2,522.5) | 4.32 (3.55,5.21) |
| | 22-24 | 53 (1,747.8) | 3.03 (2.27,3.97) | 144 (2,714.4) | 5.31 (4.47,6.25) | 51 (1,782.5) | 2.86 (2.13,3.76) | 131 (2,690.8) | 4.87 (4.07,5.78) |
| *Campylobacter jejuni* and *coli* | 1-3 | 153 (3,997.8) | 3.83 (3.24,4.48) | 2 (86.1) | 2.32 (0.28,8.39) | 147 (4,002.9) | 3.67 (3.10,4.32) | 2 (80.6) | 2.48 (0.30,8.96) |
| | 4-6 | 273 (3,674.9) | 7.43 (6.57,8.36) | 60 (513.9) | 11.68 (8.91,15.03) | 265 (3,708.5) | 7.15 (6.31,8.06) | 54 (492.6) | 10.96 (8.24,14.30) |

| | | | | | | | | | |
|---|---|---|---|---|---|---|---|---|---|
| | 7-9 | 331<br>(2,878.7) | 11.50<br>(10.29,12.81) | 186<br>(1,224.7) | 15.19<br>(13.08,17.53) | 326<br>(2,927.3) | 11.14<br>(9.96,12.41) | 177<br>(1,183.2) | 14.96<br>(12.84,17.33) |
| | 10-12 | 203<br>(2,166.8) | 9.37<br>(8.12,10.75) | 317<br>(1,929.6) | 16.43<br>(14.67,18.34) | 202<br>(2,220.0) | 9.10<br>(7.89,10.44) | 305<br>(1,885.3) | 16.18<br>(14.41,18.10) |
| | 13-15 | 116<br>(1,751.8) | 6.62<br>(5.47,7.94) | 330<br>(2,454.9) | 13.44<br>(12.03,14.97) | 120<br>(1,802.5) | 6.66<br>(5.52,7.96) | 313<br>(2,411.0) | 12.98<br>(11.58,14.50) |
| | 16-18 | 62<br>(1,509.5) | 4.11<br>(3.15,5.27) | 323<br>(2,728.2) | 11.84<br>(10.58,13.20) | 62<br>(1,559.2) | 3.98<br>(3.05,5.10) | 310<br>(2,689.4) | 11.53<br>(10.28,12.88) |
| | 19-21 | 37<br>(1,390.6) | 2.66<br>(1.87,3.67) | 272<br>(2,858.2) | 9.52<br>(8.42,10.72) | 38<br>(1,427.2) | 2.66<br>(1.88,3.65) | 267<br>(2,823.4) | 9.46<br>(8.36,10.66) |
| | 22-24 | 27<br>(1,332.1) | 2.03<br>(1.34,2.95) | 246<br>(3,026.9) | 8.13<br>(7.14,9.21) | 31<br>(1,367.1) | 2.27<br>(1.54,3.22) | 235<br>(2,995.7) | 7.84<br>(6.87,8.91) |
| *Cryptosporidium* | 1-3 | 19<br>(4,238.8) | 0.45<br>(0.27,0.70) | 0<br>(11.5) | 0.00<br>(0.00,32.08) | 19<br>(4,238.8) | 0.45<br>(0.27,0.70) | 0<br>(11.5) | 0.00<br>(0.00,32.08) |
| | 4-6 | 58<br>(4,429.7) | 1.31<br>(0.99,1.69) | 3<br>(93.4) | 3.21<br>(0.66,9.39) | 58<br>(4,429.7) | 1.31<br>(0.99,1.69) | 3<br>(93.4) | 3.21<br>(0.66,9.39) |
| | 7-9 | 118<br>(4,275.5) | 2.76<br>(2.28,3.31) | 14<br>(283.9) | 4.93<br>(2.70,8.28) | 118<br>(4,275.8) | 2.76<br>(2.28,3.30) | 15<br>(283.5) | 5.29<br>(2.96,8.73) |
| | 10-12 | 165<br>(3,836.3) | 4.30<br>(3.67,5.01) | 40<br>(641.0) | 6.24<br>(4.46,8.50) | 163<br>(3,841.6) | 4.24<br>(3.62,4.95) | 40<br>(637.5) | 6.27<br>(4.48,8.54) |
| | 13-15 | 181<br>(3,348.3) | 5.41<br>(4.65,6.25) | 61<br>(1,115.6) | 5.47<br>(4.18,7.02) | 180<br>(3,358.3) | 5.36<br>(4.61,6.20) | 58<br>(1,107.7) | 5.24<br>(3.98,6.77) |
| | 16-18 | 166<br>(2,826.0) | 5.87<br>(5.01,6.84) | 97<br>(1,583.2) | 6.13<br>(4.97,7.47) | 167<br>(2,835.0) | 5.89<br>(5.03,6.85) | 91<br>(1,577.1) | 5.77<br>(4.65,7.08) |
| | 19-21 | 128<br>(2,413.5) | 5.30<br>(4.42,6.31) | 94<br>(1,993.2) | 4.72<br>(3.81,5.77) | 127<br>(2,423.1) | 5.24<br>(4.37,6.24) | 91<br>(1,985.6) | 4.58<br>(3.69,5.63) |
| | 22-24 | 118<br>(2,099.0) | 5.62<br>(4.65,6.73) | 102<br>(2,325.0) | 4.39<br>(3.58,5.33) | 118<br>(2,106.5) | 5.60<br>(4.64,6.71) | 100<br>(2,319.1) | 4.31<br>(3.51,5.24) |
| Norovirus GII | 1-3 | 156<br>(4,008.9) | 3.89<br>(3.30,4.55) | 6<br>(92.1) | 6.52<br>(2.39,14.18) | 153<br>(4,015.2) | 3.81<br>(3.23,4.46) | 5<br>(88.8) | 5.63<br>(1.83,13.14) |
| | 4-6 | 356<br>(3,512.0) | 10.14<br>(9.11,11.25) | 58<br>(656.1) | 8.84<br>(6.71,11.43) | 351<br>(3,529.0) | 9.95<br>(8.93,11.04) | 56<br>(643.6) | 8.70<br>(6.57,11.30) |
| | 7-9 | 334<br>(2,634.9) | 12.68<br>(11.35,14.11) | 173<br>(1,589.5) | 10.88<br>(9.32,12.63) | 333<br>(2,662.8) | 12.51<br>(11.20,13.92) | 168<br>(1,566.9) | 10.72<br>(9.16,12.47) |
| | 10-12 | 188<br>(1,942.4) | 9.68<br>(8.34,11.17) | 236<br>(2,359.3) | 10.00<br>(8.77,11.36) | 190<br>(1,965.0) | 9.67<br>(8.34,11.15) | 231<br>(2,342.1) | 9.86<br>(8.63,11.22) |
| | 13-15 | 100<br>(1,592.2) | 6.28<br>(5.11,7.64) | 224<br>(2,833.4) | 7.91<br>(6.90,9.01) | 100<br>(1,612.0) | 6.20<br>(5.05,7.55) | 219<br>(2,816.9) | 7.77<br>(6.78,8.88) |
| | 16-18 | 89<br>(1,339.8) | 6.64<br>(5.33,8.17) | 181<br>(3,090.9) | 5.86<br>(5.03,6.77) | 86<br>(1,364.9) | 6.30<br>(5.04,7.78) | 174<br>(3,072.3) | 5.66<br>(4.85,6.57) |
| | 19-21 | 46<br>(1,169.7) | 3.93<br>(2.88,5.25) | 156<br>(3,276.3) | 4.76<br>(4.04,5.57) | 45<br>(1,200.8) | 3.75<br>(2.73,5.01) | 150<br>(3,249.8) | 4.62<br>(3.91,5.42) |
| | 22-24 | 23<br>(1,083.5) | 2.12<br>(1.35,3.19) | 123<br>(3,421.3) | 3.60<br>(2.99,4.29) | 24<br>(1,113.5) | 2.16<br>(1.38,3.21) | 120<br>(3,393.3) | 3.54<br>(2.93,4.23) |
| Rotavirus | 1-3 | 70<br>(2,768.5) | 2.53<br>(1.97,3.19) | 3<br>(60.2) | 4.98<br>(1.03,14.56) | 66<br>(2,780.3) | 2.37<br>(1.84,3.02) | 3<br>(54.7) | 5.48<br>(1.13,16.02) |

| | | | | | | | | | |
|---|---|---|---|---|---|---|---|---|---|
| | 4-6 | 106 (2,714.1) | 3.91 (3.20,4.72) | 8 (289.7) | 2.76 (1.19,5.44) | 105 (2,725.2) | 3.85 (3.15,4.66) | 10 (278.3) | 3.59 (1.72,6.61) |
| | 7-9 | 119 (2,417.8) | 4.92 (4.08,5.89) | 23 (626.0) | 3.67 (2.33,5.51) | 120 (2,429.5) | 4.94 (4.10,5.91) | 26 (612.7) | 4.24 (2.77,6.22) |
| | 10-12 | 102 (2,120.4) | 4.81 (3.92,5.84) | 36 (941.3) | 3.82 (2.68,5.29) | 102 (2,132.4) | 4.78 (3.90,5.81) | 38 (928.4) | 4.09 (2.90,5.62) |
| | 13-15 | 74 (1,892.3) | 3.91 (3.07,4.91) | 34 (1,199.8) | 2.83 (1.96,3.96) | 75 (1,902.7) | 3.94 (3.10,4.94) | 38 (1,187.8) | 3.20 (2.26,4.39) |
| | 16-18 | 29 (1,749.3) | 1.66 (1.11,2.38) | 29 (1,353.2) | 2.14 (1.44,3.08) | 29 (1,760.3) | 1.65 (1.10,2.37) | 32 (1,340.1) | 2.39 (1.63,3.37) |
| | 19-21 | 25 (1,663.4) | 1.50 (0.97,2.22) | 34 (1,395.5) | 2.44 (1.69,3.40) | 26 (1,671.8) | 1.56 (1.02,2.28) | 35 (1,385.9) | 2.53 (1.76,3.51) |
| | 22-24 | 21 (1,576.3) | 1.33 (0.82,2.04) | 21 (1,460.0) | 1.44 (0.89,2.20) | 23 (1,578.2) | 1.46 (0.92,2.19) | 22 (1,456.3) | 1.51 (0.95,2.29) |
| Sapovirus | 1-3 | 63 (4,163.2) | 1.51 (1.16,1.94) | 4 (38.7) | 10.34 (2.82,26.48) | 62 (4,166.3) | 1.49 (1.14,1.91) | 2 (37.6) | 5.32 (0.64,19.21) |
| | 4-6 | 229 (4,049.7) | 5.65 (4.95,6.44) | 12 (312.7) | 3.84 (1.98,6.70) | 226 (4,057.3) | 5.57 (4.87,6.35) | 9 (307.9) | 2.92 (1.34,5.55) |
| | 7-9 | 304 (3,328.8) | 9.13 (8.13,10.22) | 92 (1,003.0) | 9.17 (7.39,11.25) | 303 (3,342.6) | 9.06 (8.07,10.14) | 83 (993.9) | 8.35 (6.65,10.35) |
| | 10-12 | 271 (2,494.7) | 10.86 (9.61,12.24) | 189 (1,747.0) | 10.82 (9.33,12.48) | 272 (2,507.8) | 10.85 (9.60,12.21) | 190 (1,736.7) | 10.94 (9.44,12.61) |
| | 13-15 | 201 (1,880.8) | 10.69 (9.26,12.27) | 255 (2,400.5) | 10.62 (9.36,12.01) | 201 (1,892.4) | 10.62 (9.20,12.20) | 248 (2,394.7) | 10.36 (9.11,11.73) |
| | 16-18 | 139 (1,424.1) | 9.76 (8.21,11.53) | 284 (2,851.5) | 9.96 (8.83,11.19) | 137 (1,437.7) | 9.53 (8.00,11.26) | 283 (2,845.2) | 9.95 (8.82,11.18) |
| | 19-21 | 90 (1,137.5) | 7.91 (6.36,9.73) | 247 (3,177.6) | 7.77 (6.83,8.81) | 87 (1,157.3) | 7.52 (6.02,9.27) | 251 (3,163.9) | 7.93 (6.98,8.98) |
| | 22-24 | 62 (962.3) | 6.44 (4.94,8.26) | 241 (3,400.4) | 7.09 (6.22,8.04) | 62 (989.2) | 6.27 (4.81,8.03) | 238 (3,375.8) | 7.05 (6.18,8.01) |
| *Shigella* | 1-3 | 23 (4,234.7) | 0.54 (0.34,0.81) | 1 (14.0) | 7.14 (0.18,39.77) | 21 (4,237.5) | 0.50 (0.31,0.76) | 2 (12.5) | 15.98 (1.94,57.72) |
| | 4-6 | 51 (4,437.6) | 1.15 (0.86,1.51) | 6 (94.1) | 6.38 (2.34,13.88) | 48 (4,449.7) | 1.08 (0.80,1.43) | 6 (85.5) | 7.02 (2.58,15.28) |
| | 7-9 | 139 (4,238.0) | 3.28 (2.76,3.87) | 24 (273.0) | 8.79 (5.63,13.08) | 138 (4,254.1) | 3.24 (2.73,3.83) | 25 (259.4) | 9.64 (6.24,14.23) |
| | 10-12 | 201 (3,720.0) | 5.40 (4.68,6.20) | 64 (678.0) | 9.44 (7.27,12.05) | 199 (3,748.3) | 5.31 (4.60,6.10) | 64 (655.0) | 9.77 (7.52,12.48) |
| | 13-15 | 197 (3,194.7) | 6.17 (5.34,7.09) | 131 (1,185.6) | 11.05 (9.24,13.11) | 196 (3,220.5) | 6.09 (5.26,7.00) | 132 (1,164.6) | 11.33 (9.48,13.44) |
| | 16-18 | 221 (2,581.8) | 8.56 (7.47,9.77) | 187 (1,658.3) | 11.28 (9.72,13.01) | 216 (2,618.0) | 8.25 (7.19,9.43) | 190 (1,633.8) | 11.63 (10.03,13.41) |
| | 19-21 | 172 (2,031.8) | 8.47 (7.25,9.83) | 289 (2,087.1) | 13.85 (12.30,15.54) | 173 (2,069.6) | 8.36 (7.16,9.70) | 298 (2,056.8) | 14.49 (12.89,16.23) |
| | 22-24 | 112 (1,706.8) | 6.56 (5.40,7.90) | 321 (2,496.9) | 12.86 (11.49,14.34) | 113 (1,739.7) | 6.50 (5.35,7.81) | 328 (2,459.8) | 13.33 (11.93,14.86) |

| | | | | | | | | | |
|---|---|---|---|---|---|---|---|---|---|
| ST-ETEC | 1-3 | 99<br>(4,100.3) | 2.41<br>(1.96,2.94) | 4<br>(68.5) | 5.84<br>(1.59,14.95) | 90<br>(4,113.6) | 2.19<br>(1.76,2.69) | 4<br>(61.0) | 6.56<br>(1.79,16.79) |
| | 4-6 | 207<br>(3,967.4) | 5.22<br>(4.53,5.98) | 41<br>(392.7) | 10.44<br>(7.49,14.16) | 202<br>(4,003.5) | 5.05<br>(4.37,5.79) | 36<br>(363.1) | 9.92<br>(6.94,13.73) |
| | 7-9 | 259<br>(3,346.2) | 7.74<br>(6.83,8.74) | 112<br>(981.7) | 11.41<br>(9.39,13.73) | 259<br>(3,382.5) | 7.66<br>(6.75,8.65) | 104<br>(950.9) | 10.94<br>(8.94,13.25) |
| | 10-12 | 247<br>(2,599.5) | 9.50<br>(8.35,10.76) | 249<br>(1,579.7) | 15.76<br>(13.87,17.85) | 251<br>(2,635.6) | 9.52<br>(8.38,10.78) | 242<br>(1,549.3) | 15.62<br>(13.71,17.72) |
| | 13-15 | 167<br>(2,094.4) | 7.97<br>(6.81,9.28) | 279<br>(2,176.2) | 12.82<br>(11.36,14.42) | 169<br>(2,116.7) | 7.98<br>(6.83,9.28) | 275<br>(2,157.7) | 12.75<br>(11.28,14.34) |
| | 16-18 | 117<br>(1,697.9) | 6.89<br>(5.70,8.26) | 330<br>(2,539.2) | 13.00<br>(11.63,14.48) | 116<br>(1,722.1) | 6.74<br>(5.57,8.08) | 324<br>(2,523.5) | 12.84<br>(11.48,14.32) |
| | 19-21 | 98<br>(1,419.5) | 6.90<br>(5.60,8.41) | 336<br>(2,750.5) | 12.22<br>(10.94,13.59) | 99<br>(1,437.5) | 6.89<br>(5.60,8.38) | 334<br>(2,738.0) | 12.20<br>(10.93,13.58) |
| | 22-24 | 56<br>(1,247.3) | 4.49<br>(3.39,5.83) | 351<br>(2,988.5) | 11.74<br>(10.55,13.04) | 59<br>(1,263.6) | 4.67<br>(3.55,6.02) | 348<br>(2,974.5) | 11.70<br>(10.50,13.00) |
| tEPEC | 1-3 | 133<br>(4,013.4) | 3.31<br>(2.77,3.93) | 8<br>(90.9) | 8.80<br>(3.80,17.33) | 129<br>(4,020.9) | 3.21<br>(2.68,3.81) | 6<br>(85.1) | 7.05<br>(2.59,15.34) |
| | 4-6 | 268<br>(3,725.4) | 7.19<br>(6.36,8.11) | 47<br>(492.2) | 9.55<br>(7.02,12.70) | 268<br>(3,738.1) | 7.17<br>(6.34,8.08) | 43<br>(481.8) | 8.92<br>(6.46,12.02) |
| | 7-9 | 320<br>(2,981.7) | 10.73<br>(9.59,11.97) | 129<br>(1,232.9) | 10.46<br>(8.74,12.43) | 319<br>(2,995.5) | 10.65<br>(9.51,11.88) | 122<br>(1,221.7) | 9.99<br>(8.29,11.92) |
| | 10-12 | 219<br>(2,250.9) | 9.73<br>(8.48,11.11) | 187<br>(2,015.9) | 9.28<br>(7.99,10.71) | 218<br>(2,268.9) | 9.61<br>(8.37,10.97) | 185<br>(2,000.0) | 9.25<br>(7.97,10.68) |
| | 13-15 | 130<br>(1,828.1) | 7.11<br>(5.94,8.44) | 236<br>(2,522.6) | 9.36<br>(8.20,10.63) | 131<br>(1,843.1) | 7.11<br>(5.94,8.43) | 233<br>(2,508.9) | 9.29<br>(8.13,10.56) |
| | 16-18 | 82<br>(1,521.5) | 5.39<br>(4.29,6.69) | 235<br>(2,840.9) | 8.27<br>(7.25,9.40) | 81<br>(1,536.5) | 5.27<br>(4.19,6.55) | 231<br>(2,830.7) | 8.16<br>(7.14,9.28) |
| | 19-21 | 60<br>(1,345.9) | 4.46<br>(3.40,5.74) | 238<br>(2,996.0) | 7.94<br>(6.97,9.02) | 60<br>(1,364.1) | 4.40<br>(3.36,5.66) | 234<br>(2,981.1) | 7.85<br>(6.88,8.92) |
| | 22-24 | 44<br>(1,233.6) | 3.57<br>(2.59,4.79) | 206<br>(3,165.5) | 6.51<br>(5.65,7.46) | 45<br>(1,246.5) | 3.61<br>(2.63,4.83) | 205<br>(3,152.3) | 6.50<br>(5.64,7.46) |

**Supplemental Table 4. Median and interquartile range for age (in days) at first infection, regardless of symptomology, overall and by country.** With pathogen-attributable fraction (AFe) attribution, a pathogen was attributed to a diarrheal episode with AFe >0.5. Algorithm attribution considered both AFe and longitudinal changes in the quantity of pathogen detected. With the crude infection definition, all positives were unique infections unless positive asymptomatic and symptomatic detections were separated by <7 days. The consecutive detections definition considered both consecutive positive detections and time between consecutive positives. Italicized column headers correspond to Figure 2 data. Brazil, Peru, and South Africa had already introduced rotavirus vaccines and were not included in rotavirus analyses.

| | | **Age (days) at first infection** | | | |
|---|---|---|---|---|---|
| **Pathogen** | **Country** | ***Algorithm attribution*** | | **Afe attribution** | |
| | | **Crude** | ***Consecutive detections*** | **Crude** | **Consecutive detections** |
| Adenovirus 40/41 | All | 307.0 (183.0,458.8) | 304.0 (181.0,457.0) | 304.0 (181.0,457.0) | 304.0 (181.0,457.0) |
| | Bangladesh | 242.0 (151.0,366.5) | 217.0 (140.0,334.2) | 226.0 (143.0,335.5) | 226.0 (143.0,334.2) |
| | Brazil | 384.0 (188.2,488.0) | 384.0 (188.2,488.0) | 384.0 (188.2,488.0) | 384.0 (188.2,488.0) |
| | India | 318.0 (212.0,486.8) | 307.0 (203.0,486.0) | 307.0 (203.0,486.0) | 307.0 (203.0,486.0) |
| | Nepal | 450.0 (272.2,607.8) | 445.5 (272.8,607.2) | 445.5 (272.8,607.2) | 445.5 (272.8,607.2) |
| | Peru | 278.5 (164.0,417.0) | 243.0 (153.0,390.0) | 243.0 (153.0,390.0) | 243.0 (153.0,390.0) |
| | Pakistan | 335.5 (212.5,485.8) | 305.0 (183.5,458.5) | 305.0 (183.5,458.5) | 305.0 (183.5,458.5) |
| | South Africa | 364.0 (212.0,515.0) | 364.5 (212.5,515.0) | 364.5 (212.5,515.0) | 364.5 (212.5,515.0) |
| | Tanzania | 334.0 (161.5,449.5) | 334.0 (168.0,443.0) | 334.0 (168.0,443.0) | 334.0 (168.0,443.0) |
| Astrovirus | All | 302.0 (180.0,454.0) | 302.0 (180.0,454.0) | 285.0 (169.0,443.0) | 285.0 (169.0,443.0) |
| | Bangladesh | 271.0 (162.0,366.0) | 271.0 (162.0,366.0) | 252.0 (158.5,365.5) | 252.0 (158.5,365.5) |
| | Brazil | 517.0 (379.5,656.5) | 517.0 (379.5,656.5) | 518.5 (386.8,685.0) | 518.5 (386.8,685.0) |
| | India | 256.0 (152.0,425.0) | 256.0 (152.0,425.0) | 260.0 (151.8,413.8) | 260.0 (151.8,413.8) |
| | Nepal | 394.5 (272.2,520.5) | 394.5 (272.2,520.5) | 394.5 (272.2,520.5) | 394.5 (272.2,520.5) |
| | Peru | 308.0 (210.0,429.0) | 308.0 (210.0,429.0) | 304.0 (193.0,425.0) | 303.0 (193.0,425.0) |
| | Pakistan | 220.0 (127.5,356.0) | 220.0 (127.5,356.0) | 212.0 (106.2,334.8) | 212.0 (106.2,334.8) |
| | South Africa | 454.0 (272.2,548.0) | 454.0 (272.2,548.0) | 453.0 (267.0,548.0) | 453.0 (267.0,548.0) |
| | Tanzania | 243.0 (151.0,453.0) | 243.0 (151.0,453.0) | 243.0 (151.0,453.0) | 243.0 (151.0,453.0) |
| *Campylobacter jejuni* and *coli* | All | 244.0 (157.5,351.5) | 244.0 (157.5,351.5) | 244.0 (155.0,337.8) | 244.0 (155.0,337.8) |
| | Bangladesh | 221.0 (150.0,335.0) | 221.0 (150.0,335.0) | 214.0 (150.0,333.0) | 214.0 (150.0,333.0) |
| | Brazil | 279.0 (233.0,502.5) | 279.0 (233.0,502.5) | 278.5 (219.2,495.8) | 278.5 (219.2,495.8) |
| | India | 241.0 (152.8,336.0) | 241.0 (152.8,336.0) | 241.0 (153.0,336.0) | 241.0 (153.0,336.0) |
| | Nepal | 262.0 (184.0,339.5) | 262.0 (184.0,339.5) | 246.0 (182.5,338.0) | 246.0 (182.5,338.0) |
| | Peru | 272.0 (186.5,363.2) | 272.0 (185.8,363.2) | 272.0 (182.0,358.5) | 272.0 (182.0,358.5) |
| | Pakistan | 304.0 (210.8,455.0) | 304.0 (210.8,455.0) | 302.0 (181.5,424.0) | 302.0 (181.5,424.0) |
| | South Africa | 238.0 (127.0,327.2) | 238.0 (127.0,327.2) | 241.0 (135.0,319.5) | 241.0 (135.0,319.5) |
| | Tanzania | 188.0 (121.0,285.2) | 188.0 (121.0,285.2) | 188.0 (121.0,285.2) | 188.0 (121.0,285.2) |

| | | | | | |
|---|---|---|---|---|---|
| *Cryptosporidium* | All | 439.5 (307.0,576.0) | 439.5 (307.0,575.8) | 434.0 (307.0,576.0) | 434.0 (307.0,575.0) |
| | Bangladesh | 458.0 (305.0,559.0) | 458.0 (305.0,559.0) | 458.0 (305.0,559.0) | 458.0 (305.0,559.0) |
| | Brazil | 473.0 (405.5,551.0) | 473.0 (405.5,551.0) | 473.0 (405.5,551.0) | 473.0 (405.5,551.0) |
| | India | 413.0 (308.5,516.2) | 411.5 (308.5,516.2) | 413.0 (308.5,516.2) | 411.5 (308.5,516.2) |
| | Nepal | 549.5 (427.0,668.0) | 549.5 (427.0,668.0) | 549.5 (427.0,668.0) | 549.5 (427.0,668.0) |
| | Peru | 427.0 (334.0,547.0) | 427.0 (334.0,547.0) | 427.0 (334.0,547.0) | 427.0 (334.0,547.0) |
| | Pakistan | 424.5 (274.5,551.5) | 424.5 (274.5,551.5) | 423.0 (279.0,552.0) | 423.0 (279.0,552.0) |
| | South Africa | 487.0 (334.8,559.8) | 487.0 (334.8,559.8) | 487.0 (334.8,559.8) | 487.0 (334.8,559.8) |
| | Tanzania | 365.5 (272.8,488.5) | 365.5 (272.8,488.5) | 365.5 (272.8,488.5) | 365.5 (272.8,488.5) |
| Norovirus GII | All | 231.5 (152.0,339.0) | 231.5 (152.0,339.0) | 225.5 (151.0,340.0) | 225.5 (151.0,340.0) |
| | Bangladesh | 218.5 (165.2,303.5) | 218.5 (165.2,302.0) | 218.5 (162.5,304.0) | 218.5 (162.5,302.5) |
| | Brazil | 328.0 (194.2,424.5) | 328.0 (194.2,424.5) | 336.0 (199.5,427.5) | 336.0 (199.5,427.5) |
| | India | 250.0 (155.0,336.2) | 250.0 (155.0,336.2) | 244.0 (155.0,337.0) | 244.0 (155.0,337.0) |
| | Nepal | 275.0 (182.0,419.5) | 275.0 (182.0,419.5) | 273.0 (181.5,412.5) | 273.0 (181.5,412.5) |
| | Peru | 243.0 (181.0,317.0) | 243.0 (181.0,317.0) | 243.0 (181.0,316.5) | 243.0 (181.0,316.5) |
| | Pakistan | 183.0 (121.0,287.0) | 183.0 (120.0,287.0) | 183.0 (120.2,287.8) | 183.0 (120.2,287.8) |
| | South Africa | 275.5 (179.5,462.2) | 275.5 (179.5,462.2) | 275.5 (179.5,458.5) | 275.5 (179.5,458.5) |
| | Tanzania | 183.0 (121.0,333.0) | 183.0 (121.0,333.0) | 183.0 (121.0,306.0) | 183.0 (121.0,306.0) |
| Rotavirus | All | 273.0 (169.0,396.0) | 273.0 (169.0,396.0) | 271.0 (165.2,394.8) | 271.0 (165.2,394.8) |
| | Bangladesh | 223.0 (151.8,325.2) | 223.0 (151.8,325.2) | 223.0 (152.0,324.0) | 223.0 (152.0,324.0) |
| | India | 304.0 (200.0,399.0) | 304.0 (200.0,399.0) | 304.0 (182.2,398.2) | 304.0 (182.2,398.2) |
| | Nepal | 318.5 (212.0,472.2) | 318.5 (212.0,472.2) | 318.0 (212.0,470.0) | 318.0 (212.0,470.0) |
| | Pakistan | 303.0 (166.5,454.5) | 303.0 (166.5,454.5) | 273.0 (152.0,437.0) | 273.0 (152.0,437.0) |
| | Tanzania | 243.0 (154.0,395.0) | 243.0 (154.0,395.0) | 243.0 (154.0,395.0) | 243.0 (154.0,395.0) |
| Sapovirus | All | 307.5 (213.0,447.8) | 307.0 (213.0,447.8) | 308.0 (213.0,448.0) | 307.0 (213.0,448.0) |
| | Bangladesh | 302.0 (219.8,401.2) | 300.5 (219.8,401.2) | 302.0 (219.0,411.0) | 299.0 (219.0,411.0) |
| | Brazil | 457.0 (291.5,630.0) | 457.0 (291.5,630.0) | 457.0 (291.5,630.0) | 457.0 (291.5,630.0) |
| | India | 275.5 (187.8,393.0) | 275.5 (187.8,393.0) | 275.5 (189.2,395.2) | 275.5 (189.2,395.2) |
| | Nepal | 364.5 (249.5,484.8) | 364.5 (249.5,484.8) | 365.0 (250.0,485.5) | 365.0 (250.0,485.5) |
| | Peru | 333.0 (239.8,423.0) | 333.0 (239.8,423.0) | 333.0 (237.2,423.0) | 333.0 (237.2,423.0) |
| | Pakistan | 266.5 (181.0,365.2) | 266.5 (180.8,365.2) | 266.5 (180.2,363.8) | 266.5 (180.2,363.8) |
| | South Africa | 401.0 (242.2,545.8) | 401.0 (242.2,545.8) | 401.0 (242.5,545.5) | 401.0 (242.5,545.5) |
| | Tanzania | 303.0 (184.0,431.0) | 303.0 (184.0,431.0) | 303.0 (184.0,431.0) | 303.0 (184.0,431.0) |
| *Shigella* | All | 450.5 (314.0,573.5) | 450.5 (314.0,573.5) | 444.5 (309.8,567.2) | 444.5 (309.8,567.2) |

| | | | | | |
|---|---|---|---|---|---|
| | Bangladesh | 426.5 | 426.5 | 427.0 | 427.0 |
| | | (331.2,538.0) | (331.2,538.0) | (325.5,537.0) | (325.5,537.0) |
| | Brazil | 490.0 | 490.0 | 490.0 | 490.0 |
| | | (366.5,578.5) | (366.5,578.5) | (366.5,578.5) | (366.5,578.5) |
| | India | 425.0 | 425.0 | 425.0 | 425.0 |
| | | (305.2,549.0) | (305.0,549.0) | (304.2,549.0) | (304.2,549.0) |
| | Nepal | 556.5 | 556.5 | 550.0 | 550.0 |
| | | (427.0,638.0) | (427.0,638.0) | (425.0,638.0) | (425.0,638.0) |
| | Peru | 515.0 | 515.0 | 508.0 | 508.0 |
| | | (395.0,582.0) | (395.0,582.0) | (394.0,581.0) | (394.0,581.0) |
| | Pakistan | 426.5 | 426.5 | 427.0 | 427.0 |
| | | (305.8,563.2) | (305.8,563.2) | (303.0,564.0) | (303.0,564.0) |
| | South Africa | 455.0 | 455.0 | 455.0 | 455.0 |
| | | (310.0,606.0) | (310.0,606.0) | (305.5,605.0) | (305.5,605.0) |
| | Tanzania | 335.0 | 335.0 | 335.0 | 335.0 |
| | | (246.0,463.2) | (246.0,463.2) | (246.0,463.2) | (246.0,463.2) |
| ST-ETEC | All | 306.0 | 306.0 | 305.0 | 305.0 |
| | | (211.0,453.0) | (211.0,453.0) | (209.0,439.8) | (209.0,439.8) |
| | Bangladesh | 238.5 | 238.5 | 228.5 | 228.5 |
| | | (180.0,333.5) | (180.0,333.5) | (172.2,323.0) | (172.2,323.0) |
| | Brazil | 412.5 | 412.5 | 412.5 | 412.5 |
| | | (272.0,578.2) | (272.0,578.2) | (272.0,578.2) | (272.0,578.2) |
| | India | 306.0 | 306.0 | 306.0 | 306.0 |
| | | (243.0,437.0) | (243.0,437.0) | (242.0,429.0) | (242.0,429.0) |
| | Nepal | 333.0 | 328.0 | 328.0 | 325.0 |
| | | (212.0,430.5) | (212.0,430.5) | (212.0,430.5) | (212.0,430.5) |
| | Peru | 368.0 | 368.0 | 368.5 | 368.5 |
| | | (275.0,488.5) | (275.0,488.5) | (275.0,488.2) | (275.0,488.2) |
| | Pakistan | 335.0 | 335.0 | 334.0 | 334.0 |
| | | (183.0,499.0) | (183.0,499.0) | (182.0,486.5) | (182.0,486.5) |
| | South Africa | 457.0 | 457.0 | 457.0 | 457.0 |
| | | (326.0,580.2) | (326.0,580.2) | (334.0,580.0) | (334.0,580.0) |
| | Tanzania | 243.0 | 243.0 | 243.0 | 243.0 |
| | | (180.0,366.0) | (180.0,366.0) | (180.0,365.0) | (180.0,365.0) |
| tEPEC | All | 265.0 | 265.0 | 263.0 | 263.0 |
| | | (180.0,392.0) | (180.0,391.5) | (180.0,388.8) | (180.0,388.2) |
| | Bangladesh | 243.0 | 243.0 | 243.0 | 243.0 |
| | | (181.0,305.2) | (181.0,305.2) | (181.0,305.2) | (181.0,305.2) |
| | Brazil | 338.0 | 338.0 | 338.0 | 338.0 |
| | | (213.5,526.8) | (213.5,526.8) | (213.5,526.8) | (213.5,526.8) |
| | India | 241.0 | 241.0 | 241.0 | 241.0 |
| | | (160.2,334.0) | (160.2,334.0) | (160.2,334.0) | (160.2,334.0) |
| | Nepal | 363.0 | 363.0 | 363.0 | 363.0 |
| | | (257.2,498.5) | (257.2,498.5) | (257.2,498.5) | (257.2,498.5) |
| | Peru | 325.5 | 322.5 | 322.5 | 322.5 |
| | | (243.8,454.2) | (243.8,454.0) | (243.8,454.0) | (243.8,454.0) |
| | Pakistan | 215.5 | 215.5 | 212.0 | 212.0 |
| | | (126.0,395.5) | (126.0,395.5) | (121.0,394.0) | (121.0,394.0) |
| | South Africa | 280.0 | 280.0 | 280.0 | 280.0 |
| | | (210.8,485.5) | (210.8,485.5) | (210.8,485.5) | (210.8,485.5) |
| | Tanzania | 183.0 | 183.0 | 183.0 | 183.0 |
| | | (151.0,246.0) | (151.0,246.0) | (151.0,246.0) | (151.0,246.0) |

**Supplemental Table 5. Proportion of infections that were pathogen-attributable diarrhea and exact 95% confidence intervals (95% CIs) by 6-month age bands using different approaches to etiologic attribution (algorithm or AFe) and infection definition (crude or consecutive detections).** With pathogen-attributable fraction (AFe) attribution, a pathogen was attributed to a diarrheal episode with AFe >0.5. Algorithm attribution considered both AFe and longitudinal changes in the quantity of pathogen detected. With the crude infection definition, all positives were unique infections unless positive asymptomatic and symptomatic detections were separated by <7 days. The consecutive detections definition considered both consecutive positive detections and time between consecutive positives. Italicized column headers correspond to Figure 3 data.

| Pathogen | Age (months) | Algorithm attribution | | | | AFe attribution | | | |
|---|---|---|---|---|---|---|---|---|---|
| | | Crude | | *Consecutive detections* | | Crude | | Consecutive detections | |
| | | Diarrhea (infections) | Proportion (95% CI) | *Diarrhea (infections)* | *Proportion (95% CI)* | Diarrhea (infections) | Proportion (95% CI) | Diarrhea (infections) | Proportion (95% CI) |
| Adenovirus 40/41 | 1-6 | 142 (406) | 0.35 (0.30,0.40) | 142 (376) | 0.38 (0.33,0.43) | 83 (376) | 0.22 (0.18,0.27) | 83 (379) | 0.22 (0.18,0.26) |
| | 7-12 | 228 (702) | 0.32 (0.29,0.36) | 228 (638) | 0.36 (0.32,0.40) | 153 (652) | 0.23 (0.20,0.27) | 153 (648) | 0.24 (0.20,0.27) |
| | 13-18 | 164 (648) | 0.25 (0.22,0.29) | 164 (597) | 0.27 (0.24,0.31) | 116 (622) | 0.19 (0.16,0.22) | 116 (604) | 0.19 (0.16,0.23) |
| | 19-24 | 106 (537) | 0.20 (0.16,0.23) | 106 (503) | 0.21 (0.18,0.25) | 59 (518) | 0.11 (0.09,0.14) | 59 (506) | 0.12 (0.09,0.15) |
| Astrovirus | 1-6 | 131 (386) | 0.34 (0.29,0.39) | 131 (365) | 0.36 (0.31,0.41) | 49 (356) | 0.14 (0.10,0.18) | 49 (336) | 0.15 (0.11,0.19) |
| | 7-12 | 169 (572) | 0.30 (0.26,0.33) | 169 (533) | 0.32 (0.28,0.36) | 90 (548) | 0.16 (0.13,0.20) | 90 (515) | 0.17 (0.14,0.21) |
| | 13-18 | 155 (564) | 0.27 (0.24,0.31) | 155 (523) | 0.30 (0.26,0.34) | 103 (539) | 0.19 (0.16,0.23) | 103 (500) | 0.21 (0.17,0.24) |
| | 19-24 | 107 (409) | 0.26 (0.22,0.31) | 107 (381) | 0.28 (0.24,0.33) | 66 (388) | 0.17 (0.13,0.21) | 66 (363) | 0.18 (0.14,0.23) |
| *Campylobacter jejuni* and *coli* | 1-6 | 105 (590) | 0.18 (0.15,0.21) | 105 (488) | 0.22 (0.18,0.25) | 28 (574) | 0.05 (0.03,0.07) | 28 (468) | 0.06 (0.04,0.09) |
| | 7-12 | 204 (1,364) | 0.15 (0.13,0.17) | 204 (1,037) | 0.20 (0.17,0.22) | 74 (1,338) | 0.06 (0.04,0.07) | 74 (1,010) | 0.07 (0.06,0.09) |
| | 13-18 | 127 (1,090) | 0.12 (0.10,0.14) | 127 (831) | 0.15 (0.13,0.18) | 42 (1,066) | 0.04 (0.03,0.05) | 42 (805) | 0.05 (0.04,0.07) |
| | 19-24 | 66 (787) | 0.08 (0.07,0.11) | 66 (582) | 0.11 (0.09,0.14) | 18 (777) | 0.02 (0.01,0.04) | 18 (571) | 0.03 (0.02,0.05) |
| *Cryptosporidium* | 1-6 | 18 (89) | 0.20 (0.12,0.30) | 18 (80) | 0.22 (0.14,0.33) | 4 (89) | 0.04 (0.01,0.11) | 4 (80) | 0.05 (0.01,0.12) |
| | 7-12 | 66 (397) | 0.17 (0.13,0.21) | 66 (337) | 0.20 (0.15,0.24) | 33 (395) | 0.08 (0.06,0.12) | 33 (336) | 0.10 (0.07,0.14) |
| | 13-18 | 84 (589) | 0.14 (0.12,0.17) | 84 (505) | 0.17 (0.13,0.20) | 37 (580) | 0.06 (0.05,0.09) | 37 (496) | 0.07 (0.05,0.10) |
| | 19-24 | 77 (500) | 0.15 (0.12,0.19) | 77 (442) | 0.17 (0.14,0.21) | 45 (495) | 0.09 (0.07,0.12) | 45 (436) | 0.10 (0.08,0.14) |
| Norovirus GII | 1-6 | 106 (662) | 0.16 (0.13,0.19) | 106 (576) | 0.18 (0.15,0.22) | 31 (654) | 0.05 (0.03,0.07) | 31 (565) | 0.05 (0.04,0.08) |

| | | | | | | | | | |
|---|---|---|---|---|---|---|---|---|---|
| | 7-12 | 254 (1,060) | 0.24 (0.21,0.27) | 254 (931) | 0.27 (0.24,0.30) | 128 (1,047) | 0.12 (0.10,0.14) | 128 (922) | 0.14 (0.12,0.16) |
| | 13-18 | 152 (637) | 0.24 (0.21,0.27) | 152 (594) | 0.26 (0.22,0.29) | 90 (621) | 0.14 (0.12,0.18) | 90 (579) | 0.16 (0.13,0.19) |
| | 19-24 | 67 (369) | 0.18 (0.14,0.22) | 67 (348) | 0.19 (0.15,0.24) | 47 (360) | 0.13 (0.10,0.17) | 47 (339) | 0.14 (0.10,0.18) |
| Rotavirus | 1-6 | 109 (193) | 0.56 (0.49,0.64) | 109 (187) | 0.58 (0.51,0.65) | 103 (187) | 0.55 (0.48,0.62) | 103 (184) | 0.56 (0.48,0.63) |
| | 7-12 | 202 (287) | 0.70 (0.65,0.76) | 202 (280) | 0.72 (0.66,0.77) | 209 (291) | 0.72 (0.66,0.77) | 209 (286) | 0.73 (0.68,0.78) |
| | 13-18 | 111 (170) | 0.65 (0.58,0.72) | 111 (166) | 0.67 (0.59,0.74) | 119 (177) | 0.67 (0.60,0.74) | 119 (174) | 0.68 (0.61,0.75) |
| | 19-24 | 50 (101) | 0.50 (0.39,0.60) | 50 (101) | 0.50 (0.39,0.60) | 55 (106) | 0.52 (0.42,0.62) | 55 (106) | 0.52 (0.42,0.62) |
| Sapovirus | 1-6 | 79 (332) | 0.24 (0.19,0.29) | 79 (308) | 0.26 (0.21,0.31) | 27 (325) | 0.08 (0.06,0.12) | 27 (299) | 0.09 (0.06,0.13) |
| | 7-12 | 267 (978) | 0.27 (0.25,0.30) | 267 (856) | 0.31 (0.28,0.34) | 189 (965) | 0.20 (0.17,0.22) | 189 (848) | 0.22 (0.20,0.25) |
| | 13-18 | 266 (978) | 0.27 (0.24,0.30) | 266 (879) | 0.30 (0.27,0.33) | 208 (960) | 0.22 (0.19,0.24) | 208 (869) | 0.24 (0.21,0.27) |
| | 19-24 | 145 (705) | 0.21 (0.18,0.24) | 145 (640) | 0.23 (0.19,0.26) | 118 (691) | 0.17 (0.14,0.20) | 118 (638) | 0.18 (0.16,0.22) |
| *Shigella* | 1-6 | 27 (85) | 0.32 (0.22,0.43) | 27 (81) | 0.33 (0.23,0.45) | 19 (79) | 0.24 (0.15,0.35) | 19 (77) | 0.25 (0.16,0.36) |
| | 7-12 | 150 (529) | 0.28 (0.25,0.32) | 150 (428) | 0.35 (0.31,0.40) | 140 (520) | 0.27 (0.23,0.31) | 140 (426) | 0.33 (0.28,0.38) |
| | 13-18 | 284 (879) | 0.32 (0.29,0.36) | 284 (736) | 0.39 (0.35,0.42) | 276 (862) | 0.32 (0.29,0.35) | 276 (734) | 0.38 (0.34,0.41) |
| | 19-24 | 309 (1,163) | 0.27 (0.24,0.29) | 309 (894) | 0.35 (0.31,0.38) | 320 (1,148) | 0.28 (0.25,0.31) | 320 (912) | 0.35 (0.32,0.38) |
| ST-ETEC | 1-6 | 87 (375) | 0.23 (0.19,0.28) | 87 (351) | 0.25 (0.20,0.30) | 29 (356) | 0.08 (0.06,0.11) | 29 (332) | 0.09 (0.06,0.12) |
| | 7-12 | 215 (977) | 0.22 (0.19,0.25) | 215 (867) | 0.25 (0.22,0.28) | 143 (963) | 0.15 (0.13,0.17) | 143 (856) | 0.17 (0.14,0.19) |
| | 13-18 | 211 (1,035) | 0.20 (0.18,0.23) | 211 (893) | 0.24 (0.21,0.27) | 153 (1,015) | 0.15 (0.13,0.17) | 153 (884) | 0.17 (0.15,0.20) |
| | 19-24 | 158 (965) | 0.16 (0.14,0.19) | 158 (841) | 0.19 (0.16,0.22) | 127 (951) | 0.13 (0.11,0.16) | 127 (840) | 0.15 (0.13,0.18) |
| tEPEC | 1-6 | 98 (541) | 0.18 (0.15,0.22) | 98 (456) | 0.21 (0.18,0.26) | 18 (535) | 0.03 (0.02,0.05) | 18 (446) | 0.04 (0.02,0.06) |
| | 7-12 | 99 (1,056) | 0.09 (0.08,0.11) | 99 (855) | 0.12 (0.10,0.14) | 8 (1,048) | 0.01 (0.00,0.01) | 8 (844) | 0.01 (0.00,0.02) |
| | 13-18 | 58 (784) | 0.07 (0.06,0.09) | 58 (683) | 0.08 (0.07,0.11) | 4 (777) | 0.01 (0.00,0.01) | 4 (676) | 0.01 (0.00,0.02) |
| | 19-24 | 42 (603) | 0.07 (0.05,0.09) | 42 (548) | 0.08 (0.06,0.10) | 1 (599) | 0.00 (0.00,0.01) | 1 (544) | 0.00 (0.00,0.01) |

**Supplemental Table 6. Proportion of infections that were pathogen-attributable diarrhea and exact 95% confidence intervals (95% CIs) by 6-month age bands and infection history using different approaches to etiologic attribution (algorithm or AFe) and the consecutive detections definition for unique infections.** With pathogen-attributable fraction (AFe) attribution, a pathogen was attributed to a diarrheal episode with AFe >0.5. Algorithm attribution considered both AFe and longitudinal changes in the quantity of pathogen detected. The consecutive detections definition considered both consecutive positive detections and time between consecutive positives.

| Pathogen | Age (months) | Algorithm attribution | | | | AFe attribution | | | |
|---|---|---|---|---|---|---|---|---|---|
| | | No prior infections | | ≥1 prior infection | | No prior infections | | ≥1 prior infection | |
| | | Diarrhea (infections) | Proportion (95% CI) | Diarrhea (infections) | Proportion (95% CI) | Diarrhea (infections) | Proportion (95% CI) | Diarrhea (infections) | Proportion (95% CI) |
| Adenovirus 40/41 | 1-6 | 119 (334) | 0.36 (0.30,0.41) | 23 (42) | 0.55 (0.39,0.70) | 63 (336) | 0.19 (0.15,0.23) | 20 (43) | 0.47 (0.31,0.62) |
| | 7-12 | 116 (380) | 0.31 (0.26,0.35) | 112 (258) | 0.43 (0.37,0.50) | 57 (378) | 0.15 (0.12,0.19) | 96 (270) | 0.36 (0.30,0.42) |
| | 13-18 | 48 (280) | 0.17 (0.13,0.22) | 116 (317) | 0.37 (0.31,0.42) | 31 (280) | 0.11 (0.08,0.15) | 85 (324) | 0.26 (0.22,0.31) |
| | 19-24 | 14 (150) | 0.09 (0.05,0.15) | 92 (353) | 0.26 (0.22,0.31) | 2 (150) | 0.01 (0.00,0.05) | 57 (356) | 0.16 (0.12,0.20) |
| Astrovirus | 1-6 | 113 (332) | 0.34 (0.29,0.39) | 18 (33) | 0.55 (0.36,0.72) | 44 (316) | 0.14 (0.10,0.18) | 5 (20) | 0.25 (0.09,0.49) |
| | 7-12 | 106 (369) | 0.29 (0.24,0.34) | 63 (164) | 0.38 (0.31,0.46) | 54 (363) | 0.15 (0.11,0.19) | 36 (152) | 0.24 (0.17,0.31) |
| | 13-18 | 61 (243) | 0.25 (0.20,0.31) | 94 (280) | 0.34 (0.28,0.39) | 47 (246) | 0.19 (0.14,0.25) | 56 (254) | 0.22 (0.17,0.28) |
| | 19-24 | 24 (120) | 0.20 (0.13,0.28) | 83 (261) | 0.32 (0.26,0.38) | 21 (123) | 0.17 (0.11,0.25) | 45 (240) | 0.19 (0.14,0.24) |
| *Campylobacter jejuni* and *coli* | 1-6 | 86 (426) | 0.20 (0.16,0.24) | 19 (62) | 0.31 (0.20,0.44) | 20 (412) | 0.05 (0.03,0.07) | 8 (56) | 0.14 (0.06,0.26) |
| | 7-12 | 109 (534) | 0.20 (0.17,0.24) | 95 (503) | 0.19 (0.16,0.23) | 37 (528) | 0.07 (0.05,0.10) | 37 (482) | 0.08 (0.05,0.10) |
| | 13-18 | 31 (178) | 0.17 (0.12,0.24) | 96 (653) | 0.15 (0.12,0.18) | 10 (182) | 0.05 (0.03,0.10) | 32 (623) | 0.05 (0.04,0.07) |
| | 19-24 | 8 (64) | 0.12 (0.06,0.23) | 58 (518) | 0.11 (0.09,0.14) | 1 (69) | 0.01 (0.00,0.08) | 17 (502) | 0.03 (0.02,0.05) |
| *Cryptosporidium* | 1-6 | 18 (77) | 0.23 (0.14,0.34) | 0 (3) | 0.00 (0.00,0.71) | 4 (77) | 0.05 (0.01,0.13) | 0 (3) | 0.00 (0.00,0.71) |
| | 7-12 | 48 (283) | 0.17 (0.13,0.22) | 18 (54) | 0.33 (0.21,0.47) | 23 (281) | 0.08 (0.05,0.12) | 10 (55) | 0.18 (0.09,0.31) |
| | 13-18 | 56 (347) | 0.16 (0.12,0.20) | 28 (158) | 0.18 (0.12,0.25) | 23 (347) | 0.07 (0.04,0.10) | 14 (149) | 0.09 (0.05,0.15) |
| | 19-24 | 40 (246) | 0.16 (0.12,0.21) | 37 (196) | 0.19 (0.14,0.25) | 24 (245) | 0.10 (0.06,0.14) | 21 (191) | 0.11 (0.07,0.16) |
| Norovirus GII | 1-6 | 84 (512) | 0.16 (0.13,0.20) | 22 (64) | 0.34 (0.23,0.47) | 24 (504) | 0.05 (0.03,0.07) | 7 (61) | 0.11 (0.05,0.22) |

| | | | | | | | | | |
|---|---|---|---|---|---|---|---|---|---|
| | 7-12 | 140<br>(522) | 0.27<br>(0.23,0.31) | 114<br>(409) | 0.28<br>(0.24,0.32) | 75<br>(523) | 0.14<br>(0.11,0.18) | 53<br>(399) | 0.13<br>(0.10,0.17) |
| | 13-18 | 49<br>(189) | 0.26<br>(0.20,0.33) | 103<br>(405) | 0.25<br>(0.21,0.30) | 34<br>(186) | 0.18<br>(0.13,0.25) | 56<br>(393) | 0.14<br>(0.11,0.18) |
| | 19-24 | 9<br>(69) | 0.13<br>(0.06,0.23) | 58<br>(279) | 0.21<br>(0.16,0.26) | 7<br>(69) | 0.10<br>(0.04,0.20) | 40<br>(270) | 0.15<br>(0.11,0.20) |
| Rotavirus | 1-6 | 101<br>(176) | 0.57<br>(0.50,0.65) | 8<br>(11) | 0.73<br>(0.39,0.94) | 93<br>(171) | 0.54<br>(0.47,0.62) | 10<br>(13) | 0.77<br>(0.46,0.95) |
| | 7-12 | 155<br>(221) | 0.70<br>(0.64,0.76) | 47<br>(59) | 0.80<br>(0.67,0.89) | 157<br>(222) | 0.71<br>(0.64,0.77) | 52<br>(64) | 0.81<br>(0.70,0.90) |
| | 13-18 | 70<br>(103) | 0.68<br>(0.58,0.77) | 41<br>(63) | 0.65<br>(0.52,0.77) | 71<br>(104) | 0.68<br>(0.58,0.77) | 48<br>(70) | 0.69<br>(0.56,0.79) |
| | 19-24 | 20<br>(46) | 0.43<br>(0.29,0.59) | 30<br>(55) | 0.55<br>(0.41,0.68) | 23<br>(49) | 0.47<br>(0.33,0.62) | 32<br>(57) | 0.56<br>(0.42,0.69) |
| Sapovirus | 1-6 | 68<br>(292) | 0.23<br>(0.19,0.29) | 11<br>(16) | 0.69<br>(0.41,0.89) | 26<br>(288) | 0.09<br>(0.06,0.13) | 1<br>(11) | 0.09<br>(0.00,0.41) |
| | 7-12 | 172<br>(575) | 0.30<br>(0.26,0.34) | 95<br>(281) | 0.34<br>(0.28,0.40) | 123<br>(575) | 0.21<br>(0.18,0.25) | 66<br>(273) | 0.24<br>(0.19,0.30) |
| | 13-18 | 81<br>(340) | 0.24<br>(0.19,0.29) | 185<br>(539) | 0.34<br>(0.30,0.39) | 57<br>(338) | 0.17<br>(0.13,0.21) | 151<br>(531) | 0.28<br>(0.25,0.32) |
| | 19-24 | 24<br>(152) | 0.16<br>(0.10,0.23) | 121<br>(488) | 0.25<br>(0.21,0.29) | 17<br>(149) | 0.11<br>(0.07,0.18) | 101<br>(489) | 0.21<br>(0.17,0.25) |
| *Shigella* | 1-6 | 25<br>(74) | 0.34<br>(0.23,0.46) | 2<br>(7) | 0.29<br>(0.04,0.71) | 16<br>(69) | 0.23<br>(0.14,0.35) | 3<br>(8) | 0.38<br>(0.09,0.76) |
| | 7-12 | 120<br>(340) | 0.35<br>(0.30,0.41) | 30<br>(88) | 0.34<br>(0.24,0.45) | 107<br>(337) | 0.32<br>(0.27,0.37) | 33<br>(89) | 0.37<br>(0.27,0.48) |
| | 13-18 | 173<br>(418) | 0.41<br>(0.37,0.46) | 111<br>(318) | 0.35<br>(0.30,0.40) | 165<br>(412) | 0.40<br>(0.35,0.45) | 111<br>(322) | 0.34<br>(0.29,0.40) |
| | 19-24 | 102<br>(284) | 0.36<br>(0.30,0.42) | 207<br>(610) | 0.34<br>(0.30,0.38) | 96<br>(286) | 0.34<br>(0.28,0.39) | 224<br>(626) | 0.36<br>(0.32,0.40) |
| ST-ETEC | 1-6 | 70<br>(306) | 0.23<br>(0.18,0.28) | 17<br>(45) | 0.38<br>(0.24,0.53) | 24<br>(292) | 0.08<br>(0.05,0.12) | 5<br>(40) | 0.12<br>(0.04,0.27) |
| | 7-12 | 106<br>(506) | 0.21<br>(0.17,0.25) | 109<br>(361) | 0.30<br>(0.25,0.35) | 69<br>(510) | 0.14<br>(0.11,0.17) | 74<br>(346) | 0.21<br>(0.17,0.26) |
| | 13-18 | 41<br>(284) | 0.14<br>(0.11,0.19) | 170<br>(609) | 0.28<br>(0.24,0.32) | 23<br>(285) | 0.08<br>(0.05,0.12) | 130<br>(599) | 0.22<br>(0.18,0.25) |
| | 19-24 | 24<br>(154) | 0.16<br>(0.10,0.22) | 134<br>(687) | 0.20<br>(0.17,0.23) | 17<br>(158) | 0.11<br>(0.06,0.17) | 110<br>(682) | 0.16<br>(0.13,0.19) |
| tEPEC | 1-6 | 78<br>(401) | 0.19<br>(0.16,0.24) | 20<br>(55) | 0.36<br>(0.24,0.50) | 12<br>(397) | 0.03<br>(0.02,0.05) | 6<br>(49) | 0.12<br>(0.05,0.25) |
| | 7-12 | 59<br>(539) | 0.11<br>(0.08,0.14) | 40<br>(316) | 0.13<br>(0.09,0.17) | 5<br>(537) | 0.01<br>(0.00,0.02) | 3<br>(307) | 0.01<br>(0.00,0.03) |
| | 13-18 | 17<br>(212) | 0.08<br>(0.05,0.13) | 41<br>(471) | 0.09<br>(0.06,0.12) | 2<br>(212) | 0.01<br>(0.00,0.03) | 2<br>(464) | 0.00<br>(0.00,0.02) |
| | 19-24 | 10<br>(104) | 0.10<br>(0.05,0.17) | 32<br>(444) | 0.07<br>(0.05,0.10) | 1<br>(105) | 0.01<br>(0.00,0.05) | 0<br>(439) | 0.00<br>(0.00,0.01) |

**Supplemental Table 7. Proportion of pathogen-attributable diarrheal episodes that were severe (score >6) and exact 95% confidence intervals (95% CIs) by 6-month age bands using different approaches to etiologic attribution (algorithm or AFe).** With pathogen-attributable fraction (AFe) attribution, a pathogen was attributed to a diarrheal episode with AFe >0.5. Algorithm attribution considered both AFe and longitudinal changes in the quantity of pathogen detected. The two infection definitions used defined pathogen-attributable diarrheal episodes in the same way, so they are not separately reported. Italicized column headers correspond to Figure 3 data.

| Pathogen | Age (months) | *Algorithm attribution* | | AFe attribution | |
|---|---|---|---|---|---|
| | | *Severe diarrhea (diarrhea)* | *Proportion (95% CI)* | Severe diarrhea (diarrhea) | Proportion (95% CI) |
| Adenovirus 40/41 | 1-6 | 18 (142) | 0.127 (0.077,0.193) | 11 (83) | 0.133 (0.068,0.225) |
| | 7-12 | 38 (228) | 0.167 (0.121,0.222) | 31 (153) | 0.203 (0.142,0.275) |
| | 13-18 | 21 (164) | 0.128 (0.081,0.189) | 15 (116) | 0.129 (0.074,0.204) |
| | 19-24 | 5 (106) | 0.047 (0.015,0.107) | 4 (59) | 0.068 (0.019,0.165) |
| Astrovirus | 1-6 | 29 (131) | 0.221 (0.154,0.302) | 10 (49) | 0.204 (0.102,0.343) |
| | 7-12 | 40 (169) | 0.237 (0.175,0.308) | 20 (90) | 0.222 (0.141,0.322) |
| | 13-18 | 18 (155) | 0.116 (0.070,0.177) | 14 (103) | 0.136 (0.076,0.218) |
| | 19-24 | 6 (107) | 0.056 (0.021,0.118) | 2 (66) | 0.030 (0.004,0.105) |
| *Campylobacter jejuni* and *coli* | 1-6 | 9 (105) | 0.086 (0.040,0.156) | 0 (28) | 0.000 (0.000,0.123) |
| | 7-12 | 40 (204) | 0.196 (0.144,0.257) | 9 (74) | 0.122 (0.057,0.218) |
| | 13-18 | 10 (127) | 0.079 (0.038,0.140) | 3 (42) | 0.071 (0.015,0.195) |
| | 19-24 | 2 (66) | 0.030 (0.004,0.105) | 0 (18) | 0.000 (0.000,0.185) |
| *Cryptosporidium* | 1-6 | 5 (18) | 0.278 (0.097,0.535) | 1 (4) | 0.250 (0.006,0.806) |
| | 7-12 | 10 (66) | 0.152 (0.075,0.261) | 7 (33) | 0.212 (0.090,0.389) |
| | 13-18 | 13 (84) | 0.155 (0.085,0.250) | 5 (37) | 0.135 (0.045,0.288) |
| | 19-24 | 5 (77) | 0.065 (0.021,0.145) | 3 (45) | 0.067 (0.014,0.183) |
| Norovirus GII | 1-6 | 26 (106) | 0.245 (0.167,0.338) | 4 (31) | 0.129 (0.036,0.298) |
| | 7-12 | 35 (254) | 0.138 (0.098,0.186) | 12 (128) | 0.094 (0.049,0.158) |
| | 13-18 | 29 (152) | 0.191 (0.132,0.262) | 19 (90) | 0.211 (0.132,0.310) |
| | 19-24 | 5 (67) | 0.075 (0.025,0.166) | 5 (47) | 0.106 (0.035,0.231) |
| Rotavirus | 1-6 | 41 (109) | 0.376 (0.285,0.474) | 38 (103) | 0.369 (0.276,0.470) |
| | 7-12 | 65 (202) | 0.322 (0.258,0.391) | 65 (209) | 0.311 (0.249,0.379) |
| | 13-18 | 31 (111) | 0.279 (0.198,0.372) | 34 (119) | 0.286 (0.207,0.376) |
| | 19-24 | 5 (50) | 0.100 (0.033,0.218) | 5 (55) | 0.091 (0.030,0.200) |
| Sapovirus | 1-6 | 23 (79) | 0.291 (0.194,0.404) | 3 (27) | 0.111 (0.024,0.292) |
| | 7-12 | 50 (267) | 0.187 (0.142,0.239) | 25 (189) | 0.132 (0.087,0.189) |

| | | | | | |
|---|---|---|---|---|---|
| | 13-18 | 39 (266) | 0.147 (0.106,0.195) | 25 (208) | 0.120 (0.079,0.172) |
| | 19-24 | 13 (145) | 0.090 (0.049,0.148) | 14 (118) | 0.119 (0.066,0.191) |
| *Shigella* | 1-6 | 8 (27) | 0.296 (0.138,0.502) | 5 (19) | 0.263 (0.091,0.512) |
| | 7-12 | 24 (150) | 0.160 (0.105,0.229) | 21 (140) | 0.150 (0.095,0.220) |
| | 13-18 | 44 (284) | 0.155 (0.115,0.202) | 38 (276) | 0.138 (0.099,0.184) |
| | 19-24 | 26 (309) | 0.084 (0.056,0.121) | 26 (320) | 0.081 (0.054,0.117) |
| ST-ETEC | 1-6 | 23 (87) | 0.264 (0.176,0.370) | 8 (29) | 0.276 (0.127,0.472) |
| | 7-12 | 32 (215) | 0.149 (0.104,0.204) | 20 (143) | 0.140 (0.088,0.208) |
| | 13-18 | 32 (211) | 0.152 (0.106,0.207) | 23 (153) | 0.150 (0.098,0.217) |
| | 19-24 | 13 (158) | 0.082 (0.045,0.137) | 11 (127) | 0.087 (0.044,0.150) |
| tEPEC | 1-6 | 28 (98) | 0.286 (0.199,0.386) | 5 (18) | 0.278 (0.097,0.535) |
| | 7-12 | 17 (99) | 0.172 (0.103,0.261) | 2 (8) | 0.250 (0.032,0.651) |
| | 13-18 | 8 (58) | 0.138 (0.061,0.254) | 1 (4) | 0.250 (0.006,0.806) |
| | 19-24 | 2 (42) | 0.048 (0.006,0.162) | 0 (1) | 0.000 (0.000,0.975) |

**Supplemental Table 8. Proportion of pathogen-attributable diarrheal episodes that were severe (score >6) and exact 95% confidence intervals (95% CIs) by 6-month age bands and infection history using different approaches to etiologic attribution (algorithm or AFe) and the consecutive detections definition for unique infections.** With pathogen-attributable fraction (AFe) attribution, a pathogen was attributed to a diarrheal episode with AFe >0.5. Algorithm attribution considered both AFe and longitudinal changes in the quantity of pathogen detected. The consecutive detections definition considered both consecutive positive detections and time between consecutive positives.

| Pathogen | Age (months) | Algorithm attribution | | | | AFe attribution | | | |
|---|---|---|---|---|---|---|---|---|---|
| | | No prior infections | | ≥1 prior infection | | No prior infections | | ≥1 prior infection | |
| | | Severe diarrhea (diarrhea) | Proportion (95% CI) | Severe diarrhea (diarrhea) | Proportion (95% CI) | Severe diarrhea (diarrhea) | Proportion (95% CI) | Severe diarrhea (diarrhea) | Proportion (95% CI) |
| Adenovirus 40/41 | 1-6 | 14 (119) | 0.118 (0.066,0.190) | 4 (23) | 0.174 (0.050,0.388) | 9 (63) | 0.143 (0.067,0.254) | 2 (20) | 0.100 (0.012,0.317) |
| | 7-12 | 19 (116) | 0.164 (0.102,0.244) | 19 (112) | 0.170 (0.105,0.252) | 12 (57) | 0.211 (0.114,0.339) | 19 (96) | 0.198 (0.124,0.292) |
| | 13-18 | 6 (48) | 0.125 (0.047,0.252) | 15 (116) | 0.129 (0.074,0.204) | 4 (31) | 0.129 (0.036,0.298) | 11 (85) | 0.129 (0.066,0.220) |
| | 19-24 | 1 (14) | 0.071 (0.002,0.339) | 4 (92) | 0.043 (0.012,0.108) | 0 (2) | 0.000 (0.000,0.842) | 4 (57) | 0.070 (0.019,0.170) |
| Astrovirus | 1-6 | 27 (113) | 0.239 (0.164,0.328) | 2 (18) | 0.111 (0.014,0.347) | 10 (44) | 0.227 (0.115,0.378) | 0 (5) | 0.000 (0.000,0.522) |
| | 7-12 | 26 (106) | 0.245 (0.167,0.338) | 14 (63) | 0.222 (0.127,0.345) | 15 (54) | 0.278 (0.165,0.416) | 5 (36) | 0.139 (0.047,0.295) |
| | 13-18 | 8 (61) | 0.131 (0.058,0.242) | 10 (94) | 0.106 (0.052,0.187) | 9 (47) | 0.191 (0.091,0.333) | 5 (56) | 0.089 (0.030,0.196) |
| | 19-24 | 2 (24) | 0.083 (0.010,0.270) | 4 (83) | 0.048 (0.013,0.119) | 1 (21) | 0.048 (0.001,0.238) | 1 (45) | 0.022 (0.001,0.118) |
| *Campylobacter jejuni* and *coli* | 1-6 | 9 (86) | 0.105 (0.049,0.189) | 0 (19) | 0.000 (0.000,0.176) | 0 (20) | 0.000 (0.000,0.168) | 0 (8) | 0.000 (0.000,0.369) |
| | 7-12 | 19 (109) | 0.174 (0.108,0.259) | 21 (95) | 0.221 (0.142,0.318) | 2 (37) | 0.054 (0.007,0.182) | 7 (37) | 0.189 (0.080,0.352) |
| | 13-18 | 3 (31) | 0.097 (0.020,0.258) | 7 (96) | 0.073 (0.030,0.144) | 0 (10) | 0.000 (0.000,0.308) | 3 (32) | 0.094 (0.020,0.250) |
| | 19-24 | 1 (8) | 0.125 (0.003,0.527) | 1 (58) | 0.017 (0.000,0.092) | 0 (1) | 0.000 (0.000,0.975) | 0 (17) | 0.000 (0.000,0.195) |
| Cryptosporidium | 1-6 | 5 (18) | 0.278 (0.097,0.535) | -- | -- | 1 (4) | 0.250 (0.006,0.806) | -- | -- |
| | 7-12 | 9 (48) | 0.188 (0.089,0.326) | 1 (18) | 0.056 (0.001,0.273) | 5 (23) | 0.217 (0.075,0.437) | 2 (10) | 0.200 (0.025,0.556) |
| | 13-18 | 10 (56) | 0.179 (0.089,0.304) | 3 (28) | 0.107 (0.023,0.282) | 3 (23) | 0.130 (0.028,0.336) | 2 (14) | 0.143 (0.018,0.428) |
| | 19-24 | 2 (40) | 0.050 (0.006,0.169) | 3 (37) | 0.081 (0.017,0.219) | 2 (24) | 0.083 (0.010,0.270) | 1 (21) | 0.048 (0.001,0.238) |
| Norovirus GII | 1-6 | 20 (84) | 0.238 (0.152,0.343) | 6 (22) | 0.273 (0.107,0.502) | 3 (24) | 0.125 (0.027,0.324) | 1 (7) | 0.143 (0.004,0.579) |

| | | | | | | | | | |
|---|---|---|---|---|---|---|---|---|---|
| | 7-12 | 21<br>(140) | 0.150<br>(0.095,0.220) | 14<br>(114) | 0.123<br>(0.069,0.197) | 7<br>(75) | 0.093<br>(0.038,0.183) | 5<br>(53) | 0.094<br>(0.031,0.207) |
| | 13-18 | 9<br>(49) | 0.184<br>(0.088,0.320) | 20<br>(103) | 0.194<br>(0.123,0.284) | 5<br>(34) | 0.147<br>(0.050,0.311) | 14<br>(56) | 0.250<br>(0.144,0.384) |
| | 19-24 | 2<br>(9) | 0.222<br>(0.028,0.600) | 3<br>(58) | 0.052<br>(0.011,0.144) | 2<br>(7) | 0.286<br>(0.037,0.710) | 3<br>(40) | 0.075<br>(0.016,0.204) |
| Rotavirus | 1-6 | 40<br>(101) | 0.396<br>(0.300,0.498) | 1<br>(8) | 0.125<br>(0.003,0.527) | 36<br>(93) | 0.387<br>(0.288,0.494) | 2<br>(10) | 0.200<br>(0.025,0.556) |
| | 7-12 | 54<br>(155) | 0.348<br>(0.274,0.429) | 11<br>(47) | 0.234<br>(0.123,0.380) | 53<br>(157) | 0.338<br>(0.264,0.417) | 12<br>(52) | 0.231<br>(0.125,0.368) |
| | 13-18 | 24<br>(70) | 0.343<br>(0.233,0.466) | 7<br>(41) | 0.171<br>(0.072,0.321) | 26<br>(71) | 0.366<br>(0.255,0.489) | 8<br>(48) | 0.167<br>(0.075,0.302) |
| | 19-24 | 2<br>(20) | 0.100<br>(0.012,0.317) | 3<br>(30) | 0.100<br>(0.021,0.265) | 3<br>(23) | 0.130<br>(0.028,0.336) | 2<br>(32) | 0.062<br>(0.008,0.208) |
| Sapovirus | 1-6 | 18<br>(68) | 0.265<br>(0.165,0.386) | 5<br>(11) | 0.455<br>(0.167,0.766) | 3<br>(26) | 0.115<br>(0.024,0.302) | 0<br>(1) | 0.000<br>(0.000,0.975) |
| | 7-12 | 32<br>(172) | 0.186<br>(0.131,0.252) | 18<br>(95) | 0.189<br>(0.116,0.283) | 15<br>(123) | 0.122<br>(0.070,0.193) | 10<br>(66) | 0.152<br>(0.075,0.261) |
| | 13-18 | 10<br>(81) | 0.123<br>(0.061,0.215) | 29<br>(185) | 0.157<br>(0.108,0.217) | 7<br>(57) | 0.123<br>(0.051,0.237) | 18<br>(151) | 0.119<br>(0.072,0.182) |
| | 19-24 | 1<br>(24) | 0.042<br>(0.001,0.211) | 12<br>(121) | 0.099<br>(0.052,0.167) | 1<br>(17) | 0.059<br>(0.001,0.287) | 13<br>(101) | 0.129<br>(0.070,0.210) |
| Shigella | 1-6 | 8<br>(25) | 0.320<br>(0.149,0.535) | 0<br>(2) | 0.000<br>(0.000,0.842) | 5<br>(16) | 0.312<br>(0.110,0.587) | 0<br>(3) | 0.000<br>(0.000,0.708) |
| | 7-12 | 21<br>(120) | 0.175<br>(0.112,0.255) | 3<br>(30) | 0.100<br>(0.021,0.265) | 19<br>(107) | 0.178<br>(0.110,0.263) | 2<br>(33) | 0.061<br>(0.007,0.202) |
| | 13-18 | 30<br>(173) | 0.173<br>(0.120,0.238) | 14<br>(111) | 0.126<br>(0.071,0.203) | 27<br>(165) | 0.164<br>(0.111,0.229) | 11<br>(111) | 0.099<br>(0.051,0.170) |
| | 19-24 | 11<br>(102) | 0.108<br>(0.055,0.185) | 15<br>(207) | 0.072<br>(0.041,0.117) | 11<br>(96) | 0.115<br>(0.059,0.196) | 15<br>(224) | 0.067<br>(0.038,0.108) |
| ST-ETEC | 1-6 | 18<br>(70) | 0.257<br>(0.160,0.376) | 5<br>(17) | 0.294<br>(0.103,0.560) | 6<br>(24) | 0.250<br>(0.098,0.467) | 2<br>(5) | 0.400<br>(0.053,0.853) |
| | 7-12 | 20<br>(106) | 0.189<br>(0.119,0.276) | 12<br>(109) | 0.110<br>(0.058,0.184) | 15<br>(69) | 0.217<br>(0.127,0.333) | 5<br>(74) | 0.068<br>(0.022,0.151) |
| | 13-18 | 9<br>(41) | 0.220<br>(0.106,0.376) | 23<br>(170) | 0.135<br>(0.088,0.196) | 5<br>(23) | 0.217<br>(0.075,0.437) | 18<br>(130) | 0.138<br>(0.084,0.210) |
| | 19-24 | 5<br>(24) | 0.208<br>(0.071,0.422) | 8<br>(134) | 0.060<br>(0.026,0.114) | 2<br>(17) | 0.118<br>(0.015,0.364) | 9<br>(110) | 0.082<br>(0.038,0.150) |
| tEPEC | 1-6 | 23<br>(78) | 0.295<br>(0.197,0.409) | 5<br>(20) | 0.250<br>(0.087,0.491) | 5<br>(12) | 0.417<br>(0.152,0.723) | 0<br>(6) | 0.000<br>(0.000,0.459) |
| | 7-12 | 11<br>(59) | 0.186<br>(0.097,0.309) | 6<br>(40) | 0.150<br>(0.057,0.298) | 2<br>(5) | 0.400<br>(0.053,0.853) | 0<br>(3) | 0.000<br>(0.000,0.708) |
| | 13-18 | 5<br>(17) | 0.294<br>(0.103,0.560) | 3<br>(41) | 0.073<br>(0.015,0.199) | 1<br>(2) | 0.500<br>(0.013,0.987) | 0<br>(2) | 0.000<br>(0.000,0.842) |
| | 19-24 | 1<br>(10) | 0.100<br>(0.003,0.445) | 1<br>(32) | 0.031<br>(0.001,0.162) | 0<br>(1) | 0.000<br>(0.000,0.975) | -- | -- |

**Supplemental Table 9. Median and interquartile range (in months) for duration of infection by 6-month age bands using different approaches to etiologic attribution (algorithm or AFe) and the consecutive detections definition for unique infections.** With pathogen-attributable fraction (AFe) attribution, a pathogen was attributed to a diarrheal episode with AFe >0.5. Algorithm attribution considered both AFe and longitudinal changes in the quantity of pathogen detected. The consecutive detections definition considered both consecutive positive detections and time between consecutive positives.

| Pathogen | Age (months) | Median duration in days (interquartile range) | |
|---|---|---|---|
| | | Algorithm attribution | AFe attribution |
| Adenovirus 40/41 | 1-6 | 1 (1,1) | 1 (1,1) |
| | 7-12 | 1 (1,1) | 1 (1,1) |
| | 13-18 | 1 (1,1) | 1 (1,1) |
| | 19-24 | 1 (1,1) | 1 (1,1) |
| Astrovirus | 1-6 | 1 (1,1) | 1 (1,1) |
| | 7-12 | 1 (1,1) | 1 (1,1) |
| | 13-18 | 1 (1,1) | 1 (1,1) |
| | 19-24 | 1 (1,1) | 1 (1,1) |
| *Campylobacter jejuni* and *coli* | 1-6 | 1 (1,1) | 1 (1,1) |
| | 7-12 | 1 (1,1) | 1 (1,1) |
| | 13-18 | 1 (1,1) | 1 (1,1) |
| | 19-24 | 1 (1,1) | 1 (1,1) |
| *Cryptosporidium* | 1-6 | 1 (1,1) | 1 (1,1) |
| | 7-12 | 1 (1,1) | 1 (1,1) |
| | 13-18 | 1 (1,1) | 1 (1,1) |
| | 19-24 | 1 (1,1) | 1 (1,1) |
| Norovirus GII | 1-6 | 1 (1,1) | 1 (1,1) |
| | 7-12 | 1 (1,1) | 1 (1,1) |
| | 13-18 | 1 (1,1) | 1 (1,1) |
| | 19-24 | 1 (1,1) | 1 (1,1) |
| Rotavirus | 1-6 | 1 (1,1) | 1 (1,1) |
| | 7-12 | 1 (0,1) | 1 (0,1) |
| | 13-18 | 1 (1,1) | 1 (1,1) |
| | 19-24 | 1 (1,1) | 1 (1,1) |
| Sapovirus | 1-6 | 1 (1,1) | 1 (1,1) |
| | 7-12 | 1 (1,1) | 1 (1,1) |

| | | | |
|---|---|---|---|
| | 13-18 | 1<br>(1,1) | 1<br>(1,1) |
| | 19-24 | 1<br>(1,1) | 1<br>(1,1) |
| *Shigella* | 1-6 | 1<br>(1,1) | 1<br>(1,1) |
| | 7-12 | 1<br>(1,1) | 1<br>(1,1) |
| | 13-18 | 1<br>(1,1) | 1<br>(1,1) |
| | 19-24 | 1<br>(1,1) | 1<br>(1,1) |
| ST-ETEC | 1-6 | 1<br>(1,1) | 1<br>(1,1) |
| | 7-12 | 1<br>(1,1) | 1<br>(1,1) |
| | 13-18 | 1<br>(1,1) | 1<br>(1,1) |
| | 19-24 | 1<br>(1,1) | 1<br>(1,1) |
| tEPEC | 1-6 | 1<br>(1,1) | 1<br>(1,1) |
| | 7-12 | 1<br>(1,1) | 1<br>(1,1) |
| | 13-18 | 1<br>(1,1) | 1<br>(1,1) |
| | 19-24 | 1<br>(1,1) | 1<br>(1,1) |

**Supplemental Table 10. Median and interquartile range (in months) for duration of infection by 6-month age bands and infection history using different approaches to etiologic attribution (algorithm or AFe) and the consecutive detections definition for unique infections.** With pathogen-attributable fraction (AFe) attribution, a pathogen was attributed to a diarrheal episode with AFe >0.5. Algorithm attribution considered both AFe and longitudinal changes in the quantity of pathogen detected. The consecutive detections definition considered both consecutive positive detections and time between consecutive positives.

| Pathogen | Age (months) | Median duration in days (interquartile range) | | | |
|---|---|---|---|---|---|
| | | Algorithm attribution | | AFe attribution | |
| | | No prior infections | ≥1 prior infection | No prior infections | ≥1 prior infection |
| Adenovirus 40/41 | 1-6 | 1 (1,1) | 1 (0,1) | 1 (1,1) | 1 (0,1) |
| | 7-12 | 1 (1,1) | 1 (1,1) | 1 (1,1) | 1 (1,1) |
| | 13-18 | 1 (1,1) | 1 (1,1) | 1 (1,1) | 1 (1,1) |
| | 19-24 | 1 (1,1) | 1 (1,1) | 1 (1,1) | 1 (1,1) |
| Astrovirus | 1-6 | 1 (1,1) | 1 (1,1) | 1 (1,1) | 1 (1,1) |
| | 7-12 | 1 (1,1) | 1 (1,1) | 1 (1,1) | 1 (1,1) |
| | 13-18 | 1 (1,1) | 1 (1,1) | 1 (1,1) | 1 (1,1) |
| | 19-24 | 1 (1,1) | 1 (1,1) | 1 (1,1) | 1 (1,1) |
| *Campylobacter jejuni* and *coli* | 1-6 | 1 (1,1) | 1 (1,1) | 1 (1,1) | 1 (1,1) |
| | 7-12 | 1 (1,1) | 1 (1,1) | 1 (1,1) | 1 (1,1) |
| | 13-18 | 1 (1,1) | 1 (1,1) | 1 (1,1) | 1 (1,1) |
| | 19-24 | 1 (1,1) | 1 (1,1) | 1 (1,1) | 1 (1,1) |
| Cryptosporidium | 1-6 | 1 (1,1) | 1 (1,1) | 1 (1,1) | 1 (1,1) |
| | 7-12 | 1 (1,1) | 1 (1,1) | 1 (1,1) | 1 (1,1) |
| | 13-18 | 1 (1,1) | 1 (1,1) | 1 (1,1) | 1 (1,1) |
| | 19-24 | 1 (1,1) | 1 (1,1) | 1 (1,1) | 1 (1,1) |
| Norovirus GII | 1-6 | 1 (1,1) | 1 (1,1) | 1 (1,1) | 1 (1,1) |
| | 7-12 | 1 (1,1) | 1 (1,1) | 1 (1,1) | 1 (1,1) |
| | 13-18 | 1 (1,1) | 1 (1,1) | 1 (1,1) | 1 (1,1) |
| | 19-24 | 1 (1,1) | 1 (1,1) | 1 (1,1) | 1 (1,1) |
| Rotavirus | 1-6 | 1 (1,1) | 1 (1,1) | 1 (1,1) | 1 (1,1) |
| | 7-12 | 1 (1,1) | 1 (0,1) | 1 (1,1) | 1 (0,1) |
| | 13-18 | 1 (1,1) | 1 (1,1) | 1 (1,1) | 1 (0,1) |
| | 19-24 | 1 (1,1) | 1 (1,1) | 1 (1,1) | 1 (1,1) |
| Sapovirus | 1-6 | 1 (1,1) | 1 (1,1) | 1 (1,1) | 1 (1,1) |
| | 7-12 | 1 (1,1) | 1 (1,1) | 1 (1,1) | 1 (1,1) |

| | | | | | |
|---|---|---|---|---|---|
| | 13-18 | 1<br>(1,1) | 1<br>(1,1) | 1<br>(1,1) | 1<br>(1,1) |
| | 19-24 | 1<br>(1,1) | 1<br>(1,1) | 1<br>(1,1) | 1<br>(1,1) |
| Shigella | 1-6 | 1<br>(1,1) | 1<br>(1,1) | 1<br>(1,1) | 1<br>(0,1) |
| | 7-12 | 1<br>(1,1) | 1<br>(1,1) | 1<br>(1,1) | 1<br>(1,1) |
| | 13-18 | 1<br>(1,1) | 1<br>(1,1) | 1<br>(1,1) | 1<br>(1,1) |
| | 19-24 | 1<br>(1,1) | 1<br>(1,1) | 1<br>(1,1) | 1<br>(1,1) |
| ST-ETEC | 1-6 | 1<br>(1,1) | 1<br>(0,1) | 1<br>(1,1) | 1<br>(1,1) |
| | 7-12 | 1<br>(1,1) | 1<br>(1,1) | 1<br>(1,1) | 1<br>(1,1) |
| | 13-18 | 1<br>(1,1) | 1<br>(1,1) | 1<br>(1,1) | 1<br>(1,1) |
| | 19-24 | 1<br>(1,1) | 1<br>(1,1) | 1<br>(1,1) | 1<br>(1,1) |
| tEPEC | 1-6 | 1<br>(1,1) | 1<br>(1,1) | 1<br>(1,1) | 1<br>(1,1) |
| | 7-12 | 1<br>(1,1) | 1<br>(1,1) | 1<br>(1,1) | 1<br>(1,1) |
| | 13-18 | 1<br>(1,1) | 1<br>(1,1) | 1<br>(1,1) | 1<br>(1,1) |
| | 19-24 | 1<br>(1,1) | 1<br>(1,1) | 1<br>(1,1) | 1<br>(1,1) |

**Supplemental Table 11. Median and interquartile range (in days) for duration of pathogen-attributable diarrhea by 6-month age bands using different approaches to etiologic attribution (algorithm or AFe).** With pathogen-attributable fraction (AFe) attribution, a pathogen was attributed to a diarrheal episode with AFe >0.5. Algorithm attribution considered both AFe and longitudinal changes in the quantity of pathogen detected. Because the recorded start and end of symptoms are consistent across infection definitions, they are not separately reported. Italicized column headers correspond to Figure 4 data.

| Pathogen | Age (months) | Median duration in days (interquartile range) | |
|---|---|---|---|
| | | ***Algorithm attribution*** | **AFe attribution** |
| Adenovirus 40/41 | 1-6 | 3.0 (1.0,4.8) | 2.0 (1.0,4.0) |
| | 7-12 | 2.0 (1.0,4.0) | 2.0 (1.0,4.0) |
| | 13-18 | 2.0 (1.0,3.2) | 2.0 (1.0,3.0) |
| | 19-24 | 2.0 (1.0,3.0) | 2.0 (1.0,2.0) |
| Astrovirus | 1-6 | 3.0 (1.0,6.0) | 3.0 (1.0,5.0) |
| | 7-12 | 3.0 (2.0,5.0) | 3.0 (2.0,4.0) |
| | 13-18 | 2.0 (1.0,4.0) | 2.0 (1.0,4.0) |
| | 19-24 | 2.0 (1.0,3.0) | 2.0 (1.0,3.0) |
| *Campylobacter jejuni* and *coli* | 1-6 | 3.0 (1.0,5.0) | 2.5 (2.0,7.0) |
| | 7-12 | 3.0 (2.0,5.0) | 2.5 (1.0,4.8) |
| | 13-18 | 2.0 (1.0,4.0) | 2.0 (1.0,3.8) |
| | 19-24 | 2.0 (1.0,3.8) | 2.0 (1.0,2.8) |
| *Cryptosporidium* | 1-6 | 3.0 (2.0,7.0) | 5.5 (3.0,10.5) |
| | 7-12 | 3.0 (1.2,5.0) | 3.0 (1.0,4.0) |
| | 13-18 | 3.0 (1.0,6.0) | 2.0 (1.0,6.0) |
| | 19-24 | 2.0 (1.0,3.0) | 2.0 (2.0,4.0) |
| Norovirus GII | 1-6 | 3.0 (1.0,5.8) | 3.0 (1.0,4.5) |
| | 7-12 | 2.0 (1.0,4.0) | 2.0 (1.0,4.0) |
| | 13-18 | 2.0 (1.0,4.0) | 2.0 (1.0,3.8) |
| | 19-24 | 2.0 (1.0,4.0) | 2.0 (1.0,4.0) |
| Rotavirus | 1-6 | 3.0 (2.0,6.0) | 3.0 (2.0,5.0) |
| | 7-12 | 3.0 (2.0,4.0) | 3.0 (2.0,4.0) |
| | 13-18 | 3.0 (1.0,4.0) | 3.0 (1.0,4.0) |
| | 19-24 | 2.0 (1.0,3.0) | 2.0 (1.0,3.0) |
| Sapovirus | 1-6 | 3.0 (1.0,7.0) | 2.0 (1.0,4.0) |
| | 7-12 | 3.0 (1.0,5.0) | 2.0 (1.0,4.0) |

| | | | |
|---|---|---|---|
| | 13-18 | 2.0 (1.0,4.0) | 2.0 (1.0,4.0) |
| | 19-24 | 2.0 (1.0,4.0) | 2.0 (1.0,4.0) |
| *Shigella* | 1-6 | 3.0 (1.0,7.0) | 3.0 (1.5,7.0) |
| | 7-12 | 2.5 (1.0,5.0) | 2.0 (1.0,5.0) |
| | 13-18 | 3.0 (1.0,6.0) | 2.0 (1.0,5.0) |
| | 19-24 | 2.0 (1.0,4.0) | 2.0 (1.0,4.0) |
| ST-ETEC | 1-6 | 3.0 (1.0,6.0) | 4.0 (2.0,5.0) |
| | 7-12 | 2.0 (1.0,4.0) | 2.0 (1.0,4.0) |
| | 13-18 | 2.0 (1.0,4.0) | 2.0 (1.0,4.0) |
| | 19-24 | 2.0 (1.0,4.0) | 2.0 (1.0,4.0) |
| tEPEC | 1-6 | 3.0 (1.0,6.0) | 3.0 (2.0,5.8) |
| | 7-12 | 2.0 (1.0,5.0) | 3.5 (2.0,5.2) |
| | 13-18 | 3.0 (1.0,5.0) | 3.0 (3.0,3.0) |
| | 19-24 | 3.0 (1.0,4.0) | 11.0 (11.0,11.0) |

**Supplemental Table 12. Median and interquartile range (in days) for duration of pathogen-attributable diarrhea by 6-month age bands and infection history using different approaches to etiologic attribution (algorithm or AFe).** With pathogen-attributable fraction (AFe) attribution, a pathogen was attributed to a diarrheal episode with AFe >0.5. Algorithm attribution considered both AFe and longitudinal changes in the quantity of pathogen detected. Because the recorded start and end of symptoms are consistent across infection definitions, they are not separately reported.

| Pathogen | Age (months) | Median duration in days (interquartile range) | | | |
|---|---|---|---|---|---|
| | | Algorithm attribution | | AFe attribution | |
| | | No prior infections | ≥1 prior infection | No prior infections | ≥1 prior infection |
| Adenovirus 40/41 | 1-6 | 2.0 (1.0,4.0) | 3.0 (1.0,6.5) | 2.0 (1.0,4.0) | 3.0 (1.0,4.0) |
| | 7-12 | 3.0 (1.0,4.2) | 2.0 (1.0,4.0) | 3.0 (2.0,4.0) | 2.0 (1.0,4.0) |
| | 13-18 | 2.0 (1.0,4.0) | 2.0 (1.0,3.0) | 2.0 (1.0,3.5) | 2.0 (1.0,3.0) |
| | 19-24 | 3.0 (2.0,5.8) | 2.0 (1.0,3.0) | 4.5 (3.2,5.8) | 2.0 (1.0,2.0) |
| Astrovirus | 1-6 | 2.0 (1.0,6.0) | 3.0 (1.2,4.8) | 3.0 (1.0,5.0) | 5.0 (2.0,6.0) |
| | 7-12 | 3.0 (2.0,5.0) | 3.0 (2.0,4.0) | 3.0 (2.0,4.0) | 2.5 (1.0,3.2) |
| | 13-18 | 3.0 (1.0,4.0) | 2.0 (1.0,4.0) | 2.0 (2.0,4.0) | 2.0 (1.0,4.0) |
| | 19-24 | 2.0 (1.0,3.0) | 2.0 (1.0,3.0) | 2.0 (1.0,3.0) | 2.0 (1.0,3.0) |
| *Campylobacter jejuni* and *coli* | 1-6 | 3.0 (1.0,5.0) | 2.0 (1.5,5.0) | 3.0 (1.8,6.2) | 2.0 (2.0,7.2) |
| | 7-12 | 3.0 (2.0,5.0) | 3.0 (2.0,5.0) | 3.0 (1.0,4.0) | 2.0 (2.0,6.0) |
| | 13-18 | 3.0 (1.5,5.0) | 2.0 (1.0,4.0) | 2.5 (1.2,3.8) | 2.0 (1.0,3.2) |
| | 19-24 | 1.5 (0.8,3.2) | 2.0 (1.0,3.8) | 1.0 (1.0,1.0) | 2.0 (1.0,3.0) |
| Cryptosporidium | 1-6 | 3.0 (2.0,7.0) | NA | 5.5 (3.0,10.5) | NA |
| | 7-12 | 3.0 (2.0,6.0) | 2.0 (1.0,3.8) | 3.0 (1.5,4.5) | 2.5 (1.2,3.8) |
| | 13-18 | 3.0 (1.0,6.0) | 3.0 (1.8,5.2) | 2.0 (1.0,5.5) | 2.5 (2.0,5.2) |
| | 19-24 | 2.0 (1.0,3.0) | 2.0 (1.0,3.0) | 3.0 (2.0,4.2) | 2.0 (2.0,3.0) |
| Norovirus GII | 1-6 | 3.0 (1.0,6.0) | 3.0 (1.0,4.8) | 3.0 (1.0,4.2) | 3.0 (2.0,4.5) |
| | 7-12 | 2.0 (1.0,5.0) | 2.5 (1.0,4.0) | 2.0 (1.0,4.0) | 2.0 (1.0,4.0) |
| | 13-18 | 3.0 (2.0,4.0) | 2.0 (1.0,4.0) | 3.0 (2.0,4.0) | 2.0 (1.0,3.0) |
| | 19-24 | 2.0 (1.0,4.0) | 2.0 (1.0,4.0) | 2.0 (1.5,5.0) | 2.5 (1.0,4.0) |
| Rotavirus | 1-6 | 3.0 (2.0,6.0) | 2.5 (1.0,4.2) | 3.0 (2.0,5.0) | 2.5 (1.0,4.8) |
| | 7-12 | 3.0 (2.0,4.5) | 2.0 (1.0,4.0) | 3.0 (2.0,4.0) | 2.0 (1.0,4.0) |
| | 13-18 | 3.0 (1.0,4.8) | 2.0 (1.0,4.0) | 3.0 (1.0,5.0) | 2.0 (1.0,3.0) |
| | 19-24 | 2.5 (1.0,3.0) | 2.0 (1.0,3.0) | 2.0 (1.0,3.0) | 2.0 (1.0,3.0) |
| Sapovirus | 1-6 | 3.0 (1.0,7.0) | 5.0 (2.5,6.0) | 2.0 (1.0,4.0) | 15.0 (15.0,15.0) |
| | 7-12 | 3.0 (1.0,4.2) | 3.0 (2.0,5.0) | 3.0 (1.0,4.0) | 2.0 (1.0,4.0) |

| | | | | | |
|---|---|---|---|---|---|
| | 13-18 | 2.0 (1.0,4.0) | 2.0 (1.0,4.0) | 2.0 (1.0,4.0) | 2.0 (1.0,4.0) |
| | 19-24 | 2.0 (1.0,4.0) | 2.0 (1.0,4.0) | 2.0 (1.0,4.0) | 2.0 (1.0,4.0) |
| Shigella | 1-6 | 3.0 (1.0,7.0) | 0.5 (0.2,0.8) | 3.0 (1.8,7.0) | 2.0 (1.5,4.5) |
| | 7-12 | 3.0 (1.0,6.0) | 2.0 (1.0,3.0) | 3.0 (1.0,5.5) | 2.0 (1.0,3.0) |
| | 13-18 | 3.0 (2.0,6.0) | 2.0 (1.0,5.0) | 3.0 (2.0,6.0) | 2.0 (1.0,5.0) |
| | 19-24 | 3.0 (1.0,4.8) | 2.0 (1.0,3.0) | 3.0 (1.0,5.0) | 2.0 (1.0,3.0) |
| ST-ETEC | 1-6 | 3.0 (1.0,6.0) | 3.0 (1.0,4.0) | 3.5 (1.8,4.2) | 4.0 (3.0,5.0) |
| | 7-12 | 3.0 (1.0,5.0) | 2.0 (1.0,3.0) | 3.0 (1.0,6.0) | 2.0 (1.0,3.0) |
| | 13-18 | 3.0 (1.0,5.0) | 2.0 (1.0,4.0) | 2.0 (1.0,4.5) | 2.0 (1.0,4.0) |
| | 19-24 | 2.5 (1.0,4.2) | 2.0 (1.0,4.0) | 2.0 (1.0,3.0) | 2.0 (1.0,4.0) |
| tEPEC | 1-6 | 3.0 (1.0,6.0) | 3.0 (2.0,7.2) | 3.0 (2.0,8.8) | 2.5 (1.2,4.5) |
| | 7-12 | 2.0 (1.0,6.0) | 2.5 (1.0,4.2) | 5.0 (2.0,6.0) | 3.0 (1.5,3.5) |
| | 13-18 | 3.0 (2.0,5.0) | 3.0 (1.0,5.0) | 3.0 (3.0,3.0) | 3.0 (3.0,3.0) |
| | 19-24 | 3.0 (1.5,3.8) | 2.5 (1.0,5.0) | 11.0 (11.0,11.0) | NA |

**Supplemental Table 13. Infection incidence rates per 100 infant-months and exact 95% confidence intervals (95% CIs) by country using different approaches to etiologic attribution (algorithm or AFe) and infection definition (crude or consecutive detections).** With pathogen-attributable fraction (AFe) attribution, a pathogen was attributed to a diarrheal episode with AFe >0.5. Algorithm attribution considered both AFe and longitudinal changes in the quantity of pathogen detected. With the crude infection definition, all positives were unique infections unless positive asymptomatic and symptomatic detections were separated by <7 days. The consecutive detections definition considered both consecutive positive detections and time between consecutive positives. Italicized column headers correspond to Figure 5 data.

| Pathogen | Country | Algorithm attribution | | | | AFe attribution | | | |
|---|---|---|---|---|---|---|---|---|---|
| | | Crude | | *Consecutive detections* | | Crude | | Consecutive detections | |
| | | Infections (Infant-months) | Incidence (95% CI) | *Infections (Infant-months)* | *Incidence (95% CI)* | Infections (Infant-months) | Incidence (95% CI) | Infections (Infant-months) | Incidence (95% CI) |
| Adenovirus 40/41 | Bangladesh | 584 (4,617.7) | 12.65 (11.64,13.72) | 540 (4,296.2) | 12.57 (11.53,13.68) | 543 (4,625.9) | 11.74 (10.77,12.77) | 570 (4,275.2) | 13.33 (12.26,14.47) |
| | Brazil | 63 (2,901.8) | 2.17 (1.67,2.78) | 63 (2,852.0) | 2.21 (1.70,2.83) | 63 (2,901.8) | 2.17 (1.67,2.78) | 63 (2,852.0) | 2.21 (1.70,2.83) |
| | India | 316 (4,961.6) | 6.37 (5.69,7.11) | 282 (4,739.6) | 5.95 (5.28,6.69) | 308 (4,962.1) | 6.21 (5.53,6.94) | 282 (4,739.6) | 5.95 (5.28,6.69) |
| | Nepal | 159 (5,211.3) | 3.05 (2.60,3.56) | 150 (5,095.2) | 2.94 (2.49,3.45) | 157 (5,211.6) | 3.01 (2.56,3.52) | 150 (5,095.2) | 2.94 (2.49,3.45) |
| | Peru | 520 (4,396.4) | 11.83 (10.83,12.89) | 466 (4,099.2) | 11.37 (10.36,12.45) | 476 (4,399.7) | 10.82 (9.87,11.84) | 464 (4,099.2) | 11.32 (10.31,12.40) |
| | Pakistan | 330 (5,427.9) | 6.08 (5.44,6.77) | 309 (5,203.7) | 5.94 (5.29,6.64) | 302 (5,433.6) | 5.56 (4.95,6.22) | 304 (5,203.7) | 5.84 (5.20,6.54) |
| | South Africa | 124 (4,629.3) | 2.68 (2.23,3.19) | 123 (4,530.6) | 2.71 (2.26,3.24) | 123 (4,629.3) | 2.66 (2.21,3.17) | 123 (4,530.6) | 2.71 (2.26,3.24) |
| | Tanzania | 197 (4,317.0) | 4.56 (3.95,5.25) | 181 (4,159.1) | 4.35 (3.74,5.03) | 196 (4,317.0) | 4.54 (3.93,5.22) | 181 (4,159.1) | 4.35 (3.74,5.03) |
| Astrovirus | Bangladesh | 413 (4,641.3) | 8.90 (8.06,9.80) | 370 (4,394.6) | 8.42 (7.58,9.32) | 396 (4,642.0) | 8.53 (7.71,9.41) | 357 (4,405.1) | 8.10 (7.29,8.99) |
| | Brazil | 24 (2,898.5) | 0.83 (0.53,1.23) | 24 (2,879.7) | 0.83 (0.53,1.24) | 23 (2,898.7) | 0.79 (0.50,1.19) | 23 (2,880.3) | 0.80 (0.51,1.20) |
| | India | 235 (4,951.6) | 4.75 (4.16,5.39) | 217 (4,789.6) | 4.53 (3.95,5.18) | 225 (4,953.0) | 4.54 (3.97,5.18) | 209 (4,795.6) | 4.36 (3.79,4.99) |
| | Nepal | 186 (5,215.2) | 3.57 (3.07,4.12) | 180 (5,091.8) | 3.54 (3.04,4.09) | 186 (5,215.5) | 3.57 (3.07,4.12) | 179 (5,092.4) | 3.52 (3.02,4.07) |
| | Peru | 407 (4,444.7) | 9.16 (8.29,10.09) | 371 (4,224.2) | 8.78 (7.91,9.72) | 374 (4,448.4) | 8.41 (7.58,9.30) | 345 (4,241.9) | 8.13 (7.30,9.04) |
| | Pakistan | 413 (5,398.5) | 7.65 (6.93,8.42) | 391 (5,146.0) | 7.60 (6.86,8.39) | 375 (5,414.4) | 6.93 (6.24,7.66) | 353 (5,178.4) | 6.82 (6.12,7.57) |
| | South Africa | 111 (4,618.6) | 2.40 (1.98,2.89) | 110 (4,531.5) | 2.43 (2.00,2.93) | 110 (4,618.6) | 2.38 (1.96,2.87) | 109 (4,532.5) | 2.40 (1.97,2.90) |
| | Tanzania | 142 (4,332.6) | 3.28 (2.76,3.86) | 139 (4,218.7) | 3.29 (2.77,3.89) | 142 (4,332.6) | 3.28 (2.76,3.86) | 139 (4,218.7) | 3.29 (2.77,3.89) |
| *Campylobacter jejuni and coli* | Bangladesh | 880 (4,551.7) | 19.33 (18.08,20.65) | 647 (3,994.4) | 16.20 (14.97,17.50) | 868 (4,550.2) | 19.08 (17.83,20.39) | 627 (4,001.2) | 15.67 (14.47,16.95) |

| | | | | | | | | | |
|---|---|---|---|---|---|---|---|---|---|
| | Brazil | 33<br>(2,901.9) | 1.14<br>(0.78,1.60) | 32<br>(2,875.4) | 1.11<br>(0.76,1.57) | 32<br>(2,902.0) | 1.10<br>(0.75,1.56) | 31<br>(2,876.5) | 1.08<br>(0.73,1.53) |
| | India | 293<br>(4,948.6) | 5.92<br>(5.26,6.64) | 242<br>(4,735.0) | 5.11<br>(4.49,5.80) | 291<br>(4,948.3) | 5.88<br>(5.22,6.60) | 242<br>(4,736.0) | 5.11<br>(4.49,5.80) |
| | Nepal | 709<br>(5,113.4) | 13.87<br>(12.86,14.92) | 545<br>(4,621.5) | 11.79<br>(10.82,12.83) | 703<br>(5,115.8) | 13.74<br>(12.74,14.80) | 538<br>(4,626.2) | 11.63<br>(10.67,12.65) |
| | Peru | 610<br>(4,411.2) | 13.83<br>(12.75,14.97) | 505<br>(4,053.4) | 12.46<br>(11.40,13.59) | 599<br>(4,412.0) | 13.58<br>(12.51,14.71) | 495<br>(4,061.0) | 12.19<br>(11.14,13.31) |
| | Pakistan | 311<br>(5,449.6) | 5.71<br>(5.09,6.38) | 288<br>(5,257.0) | 5.48<br>(4.86,6.15) | 274<br>(5,460.8) | 5.02<br>(4.44,5.65) | 250<br>(5,282.4) | 4.73<br>(4.16,5.36) |
| | South Africa | 87<br>(4,613.6) | 1.89<br>(1.51,2.33) | 84<br>(4,545.1) | 1.85<br>(1.47,2.29) | 84<br>(4,614.0) | 1.82<br>(1.45,2.25) | 81<br>(4,547.1) | 1.78<br>(1.41,2.21) |
| | Tanzania | 908<br>(4,168.2) | 21.78<br>(20.39,23.25) | 595<br>(3,442.7) | 17.28<br>(15.92,18.73) | 904<br>(4,168.2) | 21.69<br>(20.30,23.15) | 590<br>(3,445.7) | 17.12<br>(15.77,18.56) |
| *Cryptosporidium* | Bangladesh | 244<br>(4,669.8) | 5.23<br>(4.59,5.92) | 197<br>(4,521.9) | 4.36<br>(3.77,5.01) | 243<br>(4,669.4) | 5.20<br>(4.57,5.90) | 197<br>(4,522.4) | 4.36<br>(3.77,5.01) |
| | Brazil | 15<br>(2,901.0) | 0.52<br>(0.29,0.85) | 15<br>(2,889.6) | 0.52<br>(0.29,0.86) | 14<br>(2,901.2) | 0.48<br>(0.26,0.81) | 14<br>(2,890.0) | 0.48<br>(0.26,0.81) |
| | India | 146<br>(4,967.0) | 2.94<br>(2.48,3.46) | 142<br>(4,858.9) | 2.92<br>(2.46,3.44) | 145<br>(4,967.0) | 2.92<br>(2.46,3.43) | 141<br>(4,859.8) | 2.90<br>(2.44,3.42) |
| | Nepal | 183<br>(5,216.8) | 3.51<br>(3.02,4.05) | 153<br>(5,088.0) | 3.01<br>(2.55,3.52) | 183<br>(5,216.5) | 3.51<br>(3.02,4.05) | 153<br>(5,088.0) | 3.01<br>(2.55,3.52) |
| | Peru | 396<br>(4,452.0) | 8.89<br>(8.04,9.82) | 310<br>(4,213.1) | 7.36<br>(6.56,8.22) | 392<br>(4,451.2) | 8.81<br>(7.96,9.72) | 308<br>(4,215.0) | 7.31<br>(6.51,8.17) |
| | Pakistan | 200<br>(5,467.8) | 3.66<br>(3.17,4.20) | 186<br>(5,341.6) | 3.48<br>(3.00,4.02) | 191<br>(5,470.7) | 3.49<br>(3.01,4.02) | 174<br>(5,348.1) | 3.25<br>(2.79,3.77) |
| | South Africa | 91<br>(4,614.4) | 1.97<br>(1.59,2.42) | 90<br>(4,542.5) | 1.98<br>(1.59,2.44) | 91<br>(4,614.1) | 1.97<br>(1.59,2.42) | 90<br>(4,542.5) | 1.98<br>(1.59,2.44) |
| | Tanzania | 300<br>(4,297.8) | 6.98<br>(6.21,7.82) | 271<br>(4,058.3) | 6.68<br>(5.91,7.52) | 300<br>(4,297.9) | 6.98<br>(6.21,7.82) | 271<br>(4,058.3) | 6.68<br>(5.91,7.52) |
| Norovirus GII | Bangladesh | 394<br>(4,645.5) | 8.48<br>(7.66,9.36) | 359<br>(4,393.4) | 8.17<br>(7.35,9.06) | 389<br>(4,644.1) | 8.38<br>(7.56,9.25) | 353<br>(4,396.8) | 8.03<br>(7.21,8.91) |
| | Brazil | 59<br>(2,896.9) | 2.04<br>(1.55,2.63) | 57<br>(2,850.7) | 2.00<br>(1.51,2.59) | 58<br>(2,897.0) | 2.00<br>(1.52,2.59) | 56<br>(2,851.7) | 1.96<br>(1.48,2.55) |
| | India | 333<br>(4,941.4) | 6.74<br>(6.03,7.50) | 308<br>(4,701.7) | 6.55<br>(5.84,7.32) | 328<br>(4,941.5) | 6.64<br>(5.94,7.40) | 302<br>(4,705.3) | 6.42<br>(5.71,7.18) |
| | Nepal | 323<br>(5,187.3) | 6.23<br>(5.57,6.94) | 299<br>(4,974.7) | 6.01<br>(5.35,6.73) | 312<br>(5,189.4) | 6.01<br>(5.36,6.72) | 288<br>(4,983.7) | 5.78<br>(5.13,6.49) |
| | Peru | 537<br>(4,433.7) | 12.11<br>(11.11,13.18) | 461<br>(4,125.3) | 11.17<br>(10.18,12.24) | 523<br>(4,433.6) | 11.80<br>(10.81,12.85) | 450<br>(4,134.3) | 10.88<br>(9.90,11.94) |
| | Pakistan | 654<br>(5,377.6) | 12.16<br>(11.25,13.13) | 561<br>(4,953.5) | 11.33<br>(10.41,12.30) | 646<br>(5,385.7) | 11.99<br>(11.09,12.96) | 553<br>(4,959.5) | 11.15<br>(10.24,12.12) |
| | South Africa | 158<br>(4,624.9) | 3.42<br>(2.90,3.99) | 151<br>(4,498.9) | 3.36<br>(2.84,3.94) | 158<br>(4,624.7) | 3.42<br>(2.90,3.99) | 151<br>(4,499.4) | 3.36<br>(2.84,3.94) |
| | Tanzania | 270<br>(4,317.2) | 6.25<br>(5.53,7.05) | 253<br>(4,103.9) | 6.16<br>(5.43,6.97) | 268<br>(4,317.6) | 6.21<br>(5.49,7.00) | 252<br>(4,105.9) | 6.14<br>(5.40,6.94) |

| | | | | | | | | | |
|---|---|---|---|---|---|---|---|---|---|
| Rotavirus | Bangladesh | 275 (4,666.2) | 5.89 (5.22,6.63) | 269 (4,527.4) | 5.94 (5.25,6.70) | 289 (4,664.9) | 6.20 (5.50,6.95) | 285 (4,518.6) | 6.31 (5.60,7.08) |
| | India | 117 (4,963.0) | 2.36 (1.95,2.83) | 116 (4,886.5) | 2.37 (1.96,2.85) | 117 (4,963.0) | 2.36 (1.95,2.83) | 117 (4,887.8) | 2.39 (1.98,2.87) |
| | Nepal | 155 (5,220.2) | 2.97 (2.52,3.48) | 146 (5,136.2) | 2.84 (2.40,3.34) | 155 (5,219.8) | 2.97 (2.52,3.48) | 147 (5,136.7) | 2.86 (2.42,3.36) |
| | Pakistan | 124 (5,480.0) | 2.26 (1.88,2.70) | 123 (5,409.7) | 2.27 (1.89,2.71) | 120 (5,481.8) | 2.19 (1.81,2.62) | 121 (5,413.3) | 2.24 (1.85,2.67) |
| | Tanzania | 80 (4,332.5) | 1.85 (1.46,2.30) | 80 (4,268.1) | 1.87 (1.49,2.33) | 80 (4,332.5) | 1.85 (1.46,2.30) | 80 (4,268.1) | 1.87 (1.49,2.33) |
| Sapovirus | Bangladesh | 550 (4,616.5) | 11.91 (10.94,12.95) | 479 (4,272.2) | 11.21 (10.23,12.26) | 543 (4,615.9) | 11.76 (10.79,12.80) | 477 (4,276.2) | 11.15 (10.18,12.20) |
| | Brazil | 70 (2,895.5) | 2.42 (1.88,3.05) | 69 (2,840.8) | 2.43 (1.89,3.07) | 69 (2,895.7) | 2.38 (1.85,3.02) | 68 (2,841.4) | 2.39 (1.86,3.03) |
| | India | 382 (4,943.1) | 7.73 (6.97,8.54) | 351 (4,672.7) | 7.51 (6.75,8.34) | 377 (4,942.9) | 7.63 (6.88,8.44) | 348 (4,676.7) | 7.44 (6.68,8.27) |
| | Nepal | 371 (5,177.6) | 7.17 (6.45,7.93) | 324 (4,936.0) | 6.56 (5.87,7.32) | 369 (5,178.2) | 7.13 (6.42,7.89) | 324 (4,938.0) | 6.56 (5.87,7.32) |
| | Peru | 509 (4,441.1) | 11.46 (10.49,12.50) | 444 (4,152.4) | 10.69 (9.72,11.73) | 496 (4,443.0) | 11.16 (10.20,12.19) | 443 (4,158.7) | 10.65 (9.68,11.69) |
| | Pakistan | 590 (5,379.5) | 10.97 (10.10,11.89) | 524 (5,002.5) | 10.47 (9.60,11.41) | 567 (5,392.1) | 10.52 (9.67,11.42) | 502 (5,018.7) | 10.00 (9.15,10.92) |
| | South Africa | 238 (4,607.0) | 5.17 (4.53,5.87) | 226 (4,417.4) | 5.12 (4.47,5.83) | 237 (4,607.2) | 5.14 (4.51,5.84) | 225 (4,418.5) | 5.09 (4.45,5.80) |
| | Tanzania | 283 (4,306.1) | 6.57 (5.83,7.38) | 266 (4,078.2) | 6.52 (5.76,7.36) | 283 (4,306.1) | 6.57 (5.83,7.38) | 267 (4,078.2) | 6.55 (5.79,7.38) |
| *Shigella* | Bangladesh | 590 (4,611.3) | 12.79 (11.78,13.87) | 492 (4,251.1) | 11.57 (10.57,12.64) | 571 (4,615.6) | 12.37 (11.38,13.43) | 496 (4,262.9) | 11.64 (10.63,12.71) |
| | Brazil | 67 (2,891.9) | 2.32 (1.80,2.94) | 62 (2,840.2) | 2.18 (1.67,2.80) | 67 (2,891.9) | 2.32 (1.80,2.94) | 62 (2,840.2) | 2.18 (1.67,2.80) |
| | India | 403 (4,931.1) | 8.17 (7.39,9.01) | 334 (4,642.7) | 7.19 (6.44,8.01) | 400 (4,931.9) | 8.11 (7.34,8.95) | 336 (4,644.3) | 7.23 (6.48,8.05) |
| | Nepal | 247 (5,198.1) | 4.75 (4.18,5.38) | 179 (5,038.1) | 3.55 (3.05,4.11) | 244 (5,199.1) | 4.69 (4.12,5.32) | 182 (5,041.3) | 3.61 (3.10,4.17) |
| | Peru | 519 (4,434.5) | 11.70 (10.72,12.76) | 390 (4,116.5) | 9.47 (8.56,10.46) | 513 (4,435.6) | 11.57 (10.59,12.61) | 405 (4,120.9) | 9.83 (8.89,10.83) |
| | Pakistan | 192 (5,459.9) | 3.52 (3.04,4.05) | 187 (5,355.8) | 3.49 (3.01,4.03) | 177 (5,464.1) | 3.24 (2.78,3.75) | 173 (5,366.4) | 3.22 (2.76,3.74) |
| | South Africa | 128 (4,620.8) | 2.77 (2.31,3.29) | 123 (4,522.1) | 2.72 (2.26,3.25) | 127 (4,620.8) | 2.75 (2.29,3.27) | 122 (4,523.1) | 2.70 (2.24,3.22) |
| | Tanzania | 510 (4,272.6) | 11.94 (10.92,13.02) | 372 (3,865.8) | 9.62 (8.67,10.65) | 510 (4,272.2) | 11.94 (10.92,13.02) | 373 (3,865.8) | 9.65 (8.69,10.68) |
| ST-ETEC | Bangladesh | 966 (4,540.0) | 21.28 (19.96,22.66) | 805 (3,939.2) | 20.44 (19.05,21.90) | 942 (4,541.2) | 20.74 (19.44,22.11) | 795 (3,953.4) | 20.11 (18.74,21.56) |
| | Brazil | 43 (2,895.9) | 1.48 (1.07,2.00) | 43 (2,860.5) | 1.50 (1.09,2.02) | 43 (2,895.9) | 1.48 (1.07,2.00) | 43 (2,860.5) | 1.50 (1.09,2.02) |

| | | | | | | | | | |
|---|---|---|---|---|---|---|---|---|---|
| | India | 394 (4,918.1) | 8.01 (7.24,8.84) | 362 (4,636.8) | 7.81 (7.02,8.65) | 387 (4,918.4) | 7.87 (7.10,8.69) | 357 (4,641.5) | 7.69 (6.91,8.53) |
| | Nepal | 477 (5,156.4) | 9.25 (8.44,10.12) | 414 (4,824.5) | 8.58 (7.77,9.45) | 474 (5,156.4) | 9.19 (8.38,10.06) | 419 (4,826.0) | 8.68 (7.87,9.55) |
| | Peru | 353 (4,462.1) | 7.91 (7.11,8.78) | 335 (4,252.3) | 7.88 (7.06,8.77) | 343 (4,463.0) | 7.69 (6.89,8.54) | 327 (4,257.8) | 7.68 (6.87,8.56) |
| | Pakistan | 317 (5,440.1) | 5.83 (5.20,6.51) | 302 (5,252.7) | 5.75 (5.12,6.44) | 296 (5,449.2) | 5.43 (4.83,6.09) | 281 (5,268.2) | 5.33 (4.73,6.00) |
| | South Africa | 108 (4,597.1) | 2.35 (1.93,2.84) | 107 (4,512.2) | 2.37 (1.94,2.87) | 107 (4,597.2) | 2.33 (1.91,2.81) | 106 (4,513.2) | 2.35 (1.92,2.84) |
| | Tanzania | 694 (4,224.0) | 16.43 (15.23,17.70) | 584 (3,671.5) | 15.91 (14.64,17.25) | 693 (4,224.0) | 16.41 (15.21,17.67) | 584 (3,672.6) | 15.90 (14.64,17.25) |
| tEPEC | Bangladesh | 659 (4,585.9) | 14.37 (13.29,15.51) | 548 (4,160.1) | 13.17 (12.09,14.32) | 659 (4,584.3) | 14.38 (13.30,15.52) | 545 (4,160.1) | 13.10 (12.02,14.25) |
| | Brazil | 51 (2,902.1) | 1.76 (1.31,2.31) | 49 (2,863.3) | 1.71 (1.27,2.26) | 51 (2,902.0) | 1.76 (1.31,2.31) | 49 (2,863.3) | 1.71 (1.27,2.26) |
| | India | 526 (4,907.5) | 10.72 (9.82,11.67) | 448 (4,511.5) | 9.93 (9.03,10.89) | 526 (4,904.9) | 10.72 (9.83,11.68) | 446 (4,511.5) | 9.89 (8.99,10.85) |
| | Nepal | 314 (5,189.6) | 6.05 (5.40,6.76) | 265 (4,963.8) | 5.34 (4.72,6.02) | 314 (5,189.2) | 6.05 (5.40,6.76) | 263 (4,963.8) | 5.30 (4.68,5.98) |
| | Peru | 461 (4,435.4) | 10.39 (9.47,11.39) | 389 (4,145.6) | 9.38 (8.47,10.36) | 460 (4,434.6) | 10.37 (9.45,11.37) | 387 (4,145.6) | 9.34 (8.43,10.31) |
| | Pakistan | 344 (5,434.0) | 6.33 (5.68,7.04) | 319 (5,229.9) | 6.10 (5.45,6.81) | 320 (5,450.3) | 5.87 (5.25,6.55) | 298 (5,247.7) | 5.68 (5.05,6.36) |
| | South Africa | 112 (4,612.3) | 2.43 (2.00,2.92) | 105 (4,524.6) | 2.32 (1.90,2.81) | 112 (4,612.0) | 2.43 (2.00,2.92) | 105 (4,524.6) | 2.32 (1.90,2.81) |
| | Tanzania | 517 (4,273.0) | 12.10 (11.08,13.19) | 419 (3,858.7) | 10.86 (9.84,11.95) | 517 (4,272.4) | 12.10 (11.08,13.19) | 417 (3,858.7) | 10.81 (9.79,11.90) |

**Supplemental Table 14. Pathogen-attributable diarrhea incidence rates per 100 infant-months and exact 95% confidence intervals (95% CIs) by country using different approaches to etiologic attribution (algorithm or AFe).** With pathogen-attributable fraction (AFe) attribution, a pathogen was attributed to a diarrheal episode with AFe >0.5. Algorithm attribution considered both AFe and longitudinal changes in the quantity of pathogen detected. Since the different infection definitions only affect sub-clinical infection durations, diarrhea rates are identical between the two definitions and are not separately reported. Italicized column headers correspond to Figure 5 data.

| Pathogen | Country | ***Algorithm attribution*** | | **AFe attribution** | |
|---|---|---|---|---|---|
| | | ***Episodes (Infant-months)*** | ***Incidence (95% CI)*** | **Episodes (Infant-months)** | **Incidence (95% CI)** |
| Adenovirus 40/41 | Bangladesh | 312 (4,671.2) | 6.68 (5.96,7.46) | 304 (4,672.9) | 6.51 (5.79,7.28) |
| | Brazil | 5 (2,913.2) | 0.17 (0.06,0.40) | 5 (2,913.2) | 0.17 (0.06,0.40) |
| | India | 41 (5,015.7) | 0.82 (0.59,1.11) | 26 (5,017.6) | 0.52 (0.34,0.76) |
| | Nepal | 24 (5,237.9) | 0.46 (0.29,0.68) | 12 (5,240.1) | 0.23 (0.12,0.40) |
| | Peru | 159 (4,467.4) | 3.56 (3.03,4.16) | 51 (4,483.3) | 1.14 (0.85,1.50) |
| | Pakistan | 82 (5,476.7) | 1.50 (1.19,1.86) | 0 (5,493.0) | 0.00 (0.00,0.07) |
| | South Africa | 6 (4,652.5) | 0.13 (0.05,0.28) | 4 (4,652.7) | 0.09 (0.02,0.22) |
| | Tanzania | 11 (4,353.6) | 0.25 (0.13,0.45) | 9 (4,353.8) | 0.21 (0.09,0.39) |
| Astrovirus | Bangladesh | 92 (4,704.4) | 1.96 (1.58,2.40) | 46 (4,710.8) | 0.98 (0.71,1.30) |
| | Brazil | 4 (2,902.5) | 0.14 (0.04,0.35) | 3 (2,902.6) | 0.10 (0.02,0.30) |
| | India | 55 (4,987.0) | 1.10 (0.83,1.44) | 37 (4,990.0) | 0.74 (0.52,1.02) |
| | Nepal | 47 (5,242.6) | 0.90 (0.66,1.19) | 39 (5,244.4) | 0.74 (0.53,1.02) |
| | Peru | 179 (4,489.6) | 3.99 (3.42,4.62) | 114 (4,499.6) | 2.53 (2.09,3.04) |
| | Pakistan | 172 (5,445.9) | 3.16 (2.70,3.67) | 58 (5,476.8) | 1.06 (0.80,1.37) |
| | South Africa | 6 (4,639.3) | 0.13 (0.05,0.28) | 4 (4,639.4) | 0.09 (0.02,0.22) |
| | Tanzania | 7 (4,359.2) | 0.16 (0.06,0.33) | 7 (4,359.2) | 0.16 (0.06,0.33) |
| *Campylobacter jejuni and coli* | Bangladesh | 83 (4,708.5) | 1.76 (1.40,2.19) | 1 (4,720.8) | 0.02 (0.00,0.12) |
| | Brazil | 7 (2,907.0) | 0.24 (0.10,0.50) | 5 (2,907.3) | 0.17 (0.06,0.40) |
| | India | 39 (4,998.6) | 0.78 (0.55,1.07) | 29 (4,999.9) | 0.58 (0.39,0.83) |
| | Nepal | 69 (5,239.3) | 1.32 (1.02,1.67) | 2 (5,253.7) | 0.04 (0.00,0.14) |
| | Peru | 181 (4,495.6) | 4.03 (3.46,4.66) | 114 (4,507.4) | 2.53 (2.09,3.04) |
| | Pakistan | 97 (5,491.7) | 1.77 (1.43,2.15) | 4 (5,513.9) | 0.07 (0.02,0.19) |
| | South Africa | 11 (4,628.6) | 0.24 (0.12,0.43) | 6 (4,629.3) | 0.13 (0.05,0.28) |
| | Tanzania | 15 (4,343.9) | 0.35 (0.19,0.57) | 1 (4,345.8) | 0.02 (0.00,0.13) |
| *Cryptosporidium* | Bangladesh | 38 (4,710.3) | 0.81 (0.57,1.11) | 18 (4,713.7) | 0.38 (0.23,0.60) |
| | Brazil | 1 (2,903.8) | 0.03 (0.00,0.19) | 0 (2,904.0) | 0.00 (0.00,0.13) |
| | India | 25 (4,990.8) | 0.50 (0.32,0.74) | 15 (4,992.5) | 0.30 (0.17,0.50) |
| | Nepal | 21 (5,248.6) | 0.40 (0.25,0.61) | 14 (5,249.8) | 0.27 (0.15,0.45) |

| | | | | | |
|---|---|---|---|---|---|
| | Peru | 89 (4,512.4) | 1.97 (1.58,2.43) | 56 (4,517.3) | 1.24 (0.94,1.61) |
| | Pakistan | 63 (5,494.7) | 1.15 (0.88,1.47) | 14 (5,505.5) | 0.25 (0.14,0.43) |
| | South Africa | 3 (4,631.7) | 0.06 (0.01,0.19) | 0 (4,632.0) | 0.00 (0.00,0.08) |
| | Tanzania | 5 (4,355.9) | 0.11 (0.04,0.27) | 2 (4,356.5) | 0.05 (0.01,0.17) |
| Norovirus GII | Bangladesh | 82 (4,706.9) | 1.74 (1.39,2.16) | 43 (4,712.1) | 0.91 (0.66,1.23) |
| | Brazil | 12 (2,906.1) | 0.41 (0.21,0.72) | 11 (2,906.3) | 0.38 (0.19,0.68) |
| | India | 42 (4,998.7) | 0.84 (0.61,1.14) | 13 (5,003.5) | 0.26 (0.14,0.44) |
| | Nepal | 95 (5,232.2) | 1.82 (1.47,2.22) | 68 (5,237.4) | 1.30 (1.01,1.65) |
| | Peru | 194 (4,501.1) | 4.31 (3.72,4.96) | 121 (4,512.6) | 2.68 (2.22,3.20) |
| | Pakistan | 125 (5,481.6) | 2.28 (1.90,2.72) | 14 (5,510.0) | 0.25 (0.14,0.43) |
| | South Africa | 11 (4,653.8) | 0.24 (0.12,0.42) | 9 (4,654.0) | 0.19 (0.09,0.37) |
| | Tanzania | 18 (4,366.8) | 0.41 (0.24,0.65) | 17 (4,367.0) | 0.39 (0.23,0.62) |
| Rotavirus | Bangladesh | 206 (4,679.8) | 4.40 (3.82,5.05) | 221 (4,678.3) | 4.72 (4.12,5.39) |
| | India | 66 (4,973.1) | 1.33 (1.03,1.69) | 67 (4,972.8) | 1.35 (1.04,1.71) |
| | Nepal | 97 (5,231.6) | 1.85 (1.50,2.26) | 98 (5,231.0) | 1.87 (1.52,2.28) |
| | Pakistan | 84 (5,487.9) | 1.53 (1.22,1.90) | 81 (5,489.5) | 1.48 (1.17,1.83) |
| | Tanzania | 19 (4,344.5) | 0.44 (0.26,0.68) | 19 (4,344.5) | 0.44 (0.26,0.68) |
| Sapovirus | Bangladesh | 124 (4,700.3) | 2.64 (2.19,3.15) | 91 (4,704.8) | 1.93 (1.56,2.37) |
| | Brazil | 9 (2,907.5) | 0.31 (0.14,0.59) | 8 (2,907.7) | 0.28 (0.12,0.54) |
| | India | 78 (5,002.9) | 1.56 (1.23,1.95) | 64 (5,004.4) | 1.28 (0.98,1.63) |
| | Nepal | 105 (5,230.0) | 2.01 (1.64,2.43) | 97 (5,231.7) | 1.85 (1.50,2.26) |
| | Peru | 221 (4,497.7) | 4.91 (4.29,5.61) | 199 (4,501.4) | 4.42 (3.83,5.08) |
| | Pakistan | 194 (5,457.4) | 3.55 (3.07,4.09) | 60 (5,491.8) | 1.09 (0.83,1.41) |
| | South Africa | 10 (4,651.8) | 0.21 (0.10,0.40) | 8 (4,652.3) | 0.17 (0.07,0.34) |
| | Tanzania | 16 (4,358.6) | 0.37 (0.21,0.60) | 15 (4,358.8) | 0.34 (0.19,0.57) |
| *Shigella* | Bangladesh | 270 (4,674.3) | 5.78 (5.11,6.51) | 275 (4,673.8) | 5.88 (5.21,6.62) |
| | Brazil | 12 (2,902.7) | 0.41 (0.21,0.72) | 12 (2,902.7) | 0.41 (0.21,0.72) |
| | India | 100 (4,990.7) | 2.00 (1.63,2.44) | 101 (4,990.7) | 2.02 (1.65,2.46) |
| | Nepal | 83 (5,230.4) | 1.59 (1.26,1.97) | 80 (5,231.4) | 1.53 (1.21,1.90) |
| | Peru | 161 (4,505.0) | 3.57 (3.04,4.17) | 162 (4,504.7) | 3.60 (3.06,4.19) |
| | Pakistan | 116 (5,474.8) | 2.12 (1.75,2.54) | 102 (5,478.9) | 1.86 (1.52,2.26) |
| | South Africa | 12 (4,643.6) | 0.26 (0.13,0.45) | 10 (4,643.8) | 0.22 (0.10,0.40) |
| | Tanzania | 16 (4,369.8) | 0.37 (0.21,0.59) | 13 (4,369.9) | 0.30 (0.16,0.51) |
| ST-ETEC | Bangladesh | 262 (4,678.4) | 5.60 (4.94,6.32) | 208 (4,685.6) | 4.44 (3.86,5.09) |

| | | | | | |
|---|---|---|---|---|---|
| | Brazil | 5<br>(2,903.3) | 0.17<br>(0.06,0.40) | 4<br>(2,903.5) | 0.14<br>(0.04,0.35) |
| | India | 73<br>(4,981.3) | 1.47<br>(1.15,1.84) | 56<br>(4,983.5) | 1.12<br>(0.85,1.46) |
| | Nepal | 87<br>(5,233.2) | 1.66<br>(1.33,2.05) | 67<br>(5,236.5) | 1.28<br>(0.99,1.62) |
| | Peru | 93<br>(4,513.2) | 2.06<br>(1.66,2.52) | 57<br>(4,519.3) | 1.26<br>(0.96,1.63) |
| | Pakistan | 117<br>(5,479.4) | 2.14<br>(1.77,2.56) | 31<br>(5,501.3) | 0.56<br>(0.38,0.80) |
| | South Africa | 5<br>(4,617.4) | 0.11<br>(0.04,0.25) | 4<br>(4,617.5) | 0.09<br>(0.02,0.22) |
| | Tanzania | 29<br>(4,354.8) | 0.67<br>(0.45,0.96) | 25<br>(4,355.5) | 0.57<br>(0.37,0.85) |
| tEPEC | Bangladesh | 39<br>(4,707.9) | 0.83<br>(0.59,1.13) | 2<br>(4,713.6) | 0.04<br>(0.01,0.15) |
| | Brazil | 1<br>(2,911.9) | 0.03<br>(0.00,0.19) | 0<br>(2,912.0) | 0.00<br>(0.00,0.13) |
| | India | 42<br>(5,002.7) | 0.84<br>(0.61,1.13) | 8<br>(5,006.8) | 0.16<br>(0.07,0.31) |
| | Nepal | 28<br>(5,245.9) | 0.53<br>(0.35,0.77) | 5<br>(5,250.0) | 0.10<br>(0.03,0.22) |
| | Peru | 58<br>(4,514.6) | 1.28<br>(0.98,1.66) | 2<br>(4,524.7) | 0.04<br>(0.01,0.16) |
| | Pakistan | 116<br>(5,478.8) | 2.12<br>(1.75,2.54) | 11<br>(5,511.1) | 0.20<br>(0.10,0.36) |
| | South Africa | 3<br>(4,633.7) | 0.06<br>(0.01,0.19) | 1<br>(4,633.9) | 0.02<br>(0.00,0.12) |
| | Tanzania | 10<br>(4,372.7) | 0.23<br>(0.11,0.42) | 2<br>(4,373.7) | 0.05<br>(0.01,0.17) |

**Supplemental Table 15. Severe (score >6) pathogen-attributable diarrhea incidence rates per 100 infant-months and exact 95% confidence intervals (95% CIs) by 3-month age bands and country using different approaches to etiologic attribution (algorithm or AFe).** With pathogen-attributable fraction (AFe) attribution, a pathogen was attributed to a diarrheal episode with AFe >0.5. Algorithm attribution considered both AFe and longitudinal changes in the quantity of pathogen detected. Since the different infection definitions only affect sub-clinical infection durations, diarrhea rates are identical between the two definitions and are not separately reported. Italicized column headers correspond to Figure 5 data.

| Pathogen | Country | *Algorithm attribution* | | AFe attribution | |
|---|---|---|---|---|---|
| | | *Episodes (Infant-months)* | *Incidence (95% CI)* | Episodes (Infant-months) | Incidence (95% CI) |
| Adenovirus 40/41 | Bangladesh | 34 (4,712.8) | 0.72 (0.50,1.01) | 39 (4,712.2) | 0.83 (0.59,1.13) |
| | Brazil | 0 (2,914.0) | 0.00 (0.00,0.13) | 0 (2,914.0) | 0.00 (0.00,0.13) |
| | India | 7 (5,020.4) | 0.14 (0.06,0.29) | 7 (5,020.4) | 0.14 (0.06,0.29) |
| | Nepal | 5 (5,241.4) | 0.10 (0.03,0.22) | 3 (5,241.9) | 0.06 (0.01,0.17) |
| | Peru | 16 (4,488.1) | 0.36 (0.20,0.58) | 12 (4,489.2) | 0.27 (0.14,0.47) |
| | Pakistan | 20 (5,486.8) | 0.36 (0.22,0.56) | 0 (5,493.0) | 0.00 (0.00,0.07) |
| | South Africa | 0 (4,653.0) | 0.00 (0.00,0.08) | 0 (4,653.0) | 0.00 (0.00,0.08) |
| | Tanzania | 0 (4,355.0) | 0.00 (0.00,0.08) | 0 (4,355.0) | 0.00 (0.00,0.08) |
| Astrovirus | Bangladesh | 11 (4,716.1) | 0.23 (0.12,0.42) | 5 (4,717.7) | 0.11 (0.03,0.25) |
| | Brazil | 0 (2,903.0) | 0.00 (0.00,0.13) | 0 (2,903.0) | 0.00 (0.00,0.13) |
| | India | 9 (4,993.0) | 0.18 (0.08,0.34) | 7 (4,993.5) | 0.14 (0.06,0.29) |
| | Nepal | 0 (5,251.0) | 0.00 (0.00,0.07) | 0 (5,251.0) | 0.00 (0.00,0.07) |
| | Peru | 16 (4,512.5) | 0.35 (0.20,0.58) | 12 (4,513.6) | 0.27 (0.14,0.46) |
| | Pakistan | 54 (5,467.3) | 0.99 (0.74,1.29) | 19 (5,484.0) | 0.35 (0.21,0.54) |
| | South Africa | 1 (4,639.8) | 0.02 (0.00,0.12) | 1 (4,639.8) | 0.02 (0.00,0.12) |
| | Tanzania | 2 (4,359.6) | 0.05 (0.01,0.17) | 2 (4,359.6) | 0.05 (0.01,0.17) |
| *Campylobacter jejuni and coli* | Bangladesh | 5 (4,719.5) | 0.11 (0.03,0.25) | 0 (4,721.0) | 0.00 (0.00,0.08) |
| | Brazil | 0 (2,908.0) | 0.00 (0.00,0.13) | 0 (2,908.0) | 0.00 (0.00,0.13) |
| | India | 1 (5,003.8) | 0.02 (0.00,0.11) | 0 (5,004.0) | 0.00 (0.00,0.07) |
| | Nepal | 10 (5,250.4) | 0.19 (0.09,0.35) | 0 (5,254.0) | 0.00 (0.00,0.07) |
| | Peru | 18 (4,524.1) | 0.40 (0.24,0.63) | 11 (4,525.8) | 0.24 (0.12,0.43) |
| | Pakistan | 26 (5,504.2) | 0.47 (0.31,0.69) | 1 (5,514.4) | 0.02 (0.00,0.10) |
| | South Africa | 0 (4,630.0) | 0.00 (0.00,0.08) | 0 (4,630.0) | 0.00 (0.00,0.08) |
| | Tanzania | 1 (4,345.9) | 0.02 (0.00,0.13) | 0 (4,346.0) | 0.00 (0.00,0.08) |
| *Cryptosporidium* | Bangladesh | 5 (4,715.5) | 0.11 (0.03,0.25) | 3 (4,715.9) | 0.06 (0.01,0.19) |
| | Brazil | 0 (2,904.0) | 0.00 (0.00,0.13) | 0 (2,904.0) | 0.00 (0.00,0.13) |
| | India | 3 (4,994.4) | 0.06 (0.01,0.18) | 2 (4,994.6) | 0.04 (0.00,0.14) |
| | Nepal | 3 (5,252.3) | 0.06 (0.01,0.17) | 3 (5,252.3) | 0.06 (0.01,0.17) |

| | | | | | |
|---|---|---|---|---|---|
| | Peru | 4 (4,524.9) | 0.09 (0.02,0.23) | 5 (4,524.8) | 0.11 (0.04,0.26) |
| | Pakistan | 17 (5,502.6) | 0.31 (0.18,0.49) | 3 (5,507.8) | 0.05 (0.01,0.16) |
| | South Africa | 0 (4,632.0) | 0.00 (0.00,0.08) | 0 (4,632.0) | 0.00 (0.00,0.08) |
| | Tanzania | 1 (4,356.6) | 0.02 (0.00,0.13) | 0 (4,357.0) | 0.00 (0.00,0.08) |
| Norovirus GII | Bangladesh | 9 (4,717.0) | 0.19 (0.09,0.36) | 7 (4,717.4) | 0.15 (0.06,0.31) |
| | Brazil | 0 (2,908.0) | 0.00 (0.00,0.13) | 0 (2,908.0) | 0.00 (0.00,0.13) |
| | India | 4 (5,003.7) | 0.08 (0.02,0.20) | 1 (5,004.9) | 0.02 (0.00,0.11) |
| | Nepal | 11 (5,247.3) | 0.21 (0.10,0.38) | 6 (5,248.4) | 0.11 (0.04,0.25) |
| | Peru | 32 (4,524.6) | 0.71 (0.48,1.00) | 20 (4,527.1) | 0.44 (0.27,0.68) |
| | Pakistan | 39 (5,499.0) | 0.71 (0.50,0.97) | 6 (5,511.1) | 0.11 (0.04,0.24) |
| | South Africa | 0 (4,655.0) | 0.00 (0.00,0.08) | 0 (4,655.0) | 0.00 (0.00,0.08) |
| | Tanzania | 0 (4,369.0) | 0.00 (0.00,0.08) | 0 (4,369.0) | 0.00 (0.00,0.08) |
| Rotavirus | Bangladesh | 55 (4,700.7) | 1.17 (0.88,1.52) | 56 (4,700.7) | 1.19 (0.90,1.55) |
| | India | 12 (4,980.3) | 0.24 (0.12,0.42) | 13 (4,980.1) | 0.26 (0.14,0.45) |
| | Nepal | 26 (5,244.1) | 0.50 (0.32,0.73) | 27 (5,243.6) | 0.51 (0.34,0.75) |
| | Pakistan | 46 (5,493.3) | 0.84 (0.61,1.12) | 43 (5,494.9) | 0.78 (0.57,1.05) |
| | Tanzania | 3 (4,346.6) | 0.07 (0.01,0.20) | 3 (4,346.6) | 0.07 (0.01,0.20) |
| Sapovirus | Bangladesh | 14 (4,715.2) | 0.30 (0.16,0.50) | 12 (4,715.4) | 0.25 (0.13,0.44) |
| | Brazil | 1 (2,908.9) | 0.03 (0.00,0.19) | 0 (2,909.0) | 0.00 (0.00,0.13) |
| | India | 6 (5,011.0) | 0.12 (0.04,0.26) | 5 (5,011.2) | 0.10 (0.03,0.23) |
| | Nepal | 17 (5,246.6) | 0.32 (0.19,0.52) | 14 (5,247.6) | 0.27 (0.15,0.45) |
| | Peru | 18 (4,527.6) | 0.40 (0.24,0.63) | 15 (4,528.1) | 0.33 (0.19,0.55) |
| | Pakistan | 65 (5,481.5) | 1.19 (0.92,1.51) | 18 (5,500.7) | 0.33 (0.19,0.52) |
| | South Africa | 1 (4,652.6) | 0.02 (0.00,0.12) | 0 (4,653.0) | 0.00 (0.00,0.08) |
| | Tanzania | 3 (4,360.3) | 0.07 (0.01,0.20) | 3 (4,360.3) | 0.07 (0.01,0.20) |
| *Shigella* | Bangladesh | 17 (4,711.2) | 0.36 (0.21,0.58) | 17 (4,711.0) | 0.36 (0.21,0.58) |
| | Brazil | 0 (2,905.0) | 0.00 (0.00,0.13) | 0 (2,905.0) | 0.00 (0.00,0.13) |
| | India | 14 (5,001.5) | 0.28 (0.15,0.47) | 13 (5,001.7) | 0.26 (0.14,0.44) |
| | Nepal | 13 (5,244.0) | 0.25 (0.13,0.42) | 10 (5,245.0) | 0.19 (0.09,0.35) |
| | Peru | 15 (4,527.0) | 0.33 (0.19,0.55) | 14 (4,527.2) | 0.31 (0.17,0.52) |
| | Pakistan | 41 (5,492.6) | 0.75 (0.54,1.01) | 34 (5,495.0) | 0.62 (0.43,0.86) |
| | South Africa | 1 (4,644.7) | 0.02 (0.00,0.12) | 1 (4,644.7) | 0.02 (0.00,0.12) |
| | Tanzania | 1 (4,371.6) | 0.02 (0.00,0.13) | 1 (4,371.6) | 0.02 (0.00,0.13) |
| ST-ETEC | Bangladesh | 28 (4,710.3) | 0.59 (0.40,0.86) | 22 (4,711.8) | 0.47 (0.29,0.71) |

| | | | | | |
|---|---|---|---|---|---|
| | Brazil | 0<br>(2,904.0) | 0.00<br>(0.00,0.13) | 0<br>(2,904.0) | 0.00<br>(0.00,0.13) |
| | India | 5<br>(4,989.1) | 0.10<br>(0.03,0.23) | 4<br>(4,989.3) | 0.08<br>(0.02,0.21) |
| | Nepal | 18<br>(5,245.9) | 0.34<br>(0.20,0.54) | 15<br>(5,246.5) | 0.29<br>(0.16,0.47) |
| | Peru | 12<br>(4,526.4) | 0.27<br>(0.14,0.46) | 8<br>(4,527.1) | 0.18<br>(0.08,0.35) |
| | Pakistan | 34<br>(5,495.0) | 0.62<br>(0.43,0.86) | 10<br>(5,505.6) | 0.18<br>(0.09,0.33) |
| | South Africa | 0<br>(4,618.0) | 0.00<br>(0.00,0.08) | 0<br>(4,618.0) | 0.00<br>(0.00,0.08) |
| | Tanzania | 3<br>(4,358.3) | 0.07<br>(0.01,0.20) | 3<br>(4,358.3) | 0.07<br>(0.01,0.20) |
| tEPEC | Bangladesh | 4<br>(4,712.8) | 0.08<br>(0.02,0.22) | 1<br>(4,713.8) | 0.02<br>(0.00,0.12) |
| | Brazil | 0<br>(2,912.0) | 0.00<br>(0.00,0.13) | 0<br>(2,912.0) | 0.00<br>(0.00,0.13) |
| | India | 1<br>(5,007.6) | 0.02<br>(0.00,0.11) | 1<br>(5,007.6) | 0.02<br>(0.00,0.11) |
| | Nepal | 3<br>(5,249.8) | 0.06<br>(0.01,0.17) | 1<br>(5,250.7) | 0.02<br>(0.00,0.11) |
| | Peru | 4<br>(4,524.1) | 0.09<br>(0.02,0.23) | 0<br>(4,525.0) | 0.00<br>(0.00,0.08) |
| | Pakistan | 43<br>(5,492.5) | 0.78<br>(0.57,1.05) | 5<br>(5,512.3) | 0.09<br>(0.03,0.21) |
| | South Africa | 0<br>(4,634.0) | 0.00<br>(0.00,0.08) | 0<br>(4,634.0) | 0.00<br>(0.00,0.08) |
| | Tanzania | 0<br>(4,374.0) | 0.00<br>(0.00,0.08) | 0<br>(4,374.0) | 0.00<br>(0.00,0.08) |

**Supplemental Table 16. Proportion of infections that were pathogen-attributable diarrhea and proportion of pathogen-attributable diarrheal episodes that were severe (score >6) and exact 95% confidence intervals (95% CIs) by 3-month age bands using algorithm attribution and different approaches to infection definition (crude or consecutive detections).** Algorithm attribution considered both pathogen-attributable fraction and longitudinal changes in the quantity of pathogen detected. With the crude infection definition, all positives were unique infections unless positive asymptomatic and symptomatic detections were separated by <7 days. The consecutive detections definition considered both consecutive positive detections and time between consecutive positives. The two infection definitions used defined pathogen-attributable diarrheal episodes in the same way, so they are not separately reported for the proportion of attributable diarrheal episodes that were severe.

| Pathogen | Age (months) | Proportion of infections that were pathogen-attributable diarrhea | | | | Proportion of pathogen-attributable diarrheal episodes that were severe | |
|---|---|---|---|---|---|---|---|
| | | Crude | | Consecutive detections | | | |
| | | Diarrhea (infections) | Proportion (95% CI) | Diarrhea (infections) | Proportion (95% CI) | Severe diarrhea (diarrhea) | Proportion (95% CI) |
| Adenovirus 40/41 | 1-3 | 43 (127) | 0.34 (0.26,0.43) | 43 (124) | 0.35 (0.26,0.44) | 5 (43) | 0.116 (0.039,0.251) |
| | 4-6 | 99 (279) | 0.35 (0.30,0.41) | 99 (252) | 0.39 (0.33,0.46) | 13 (99) | 0.131 (0.072,0.214) |
| | 7-9 | 125 (355) | 0.35 (0.30,0.40) | 125 (320) | 0.39 (0.34,0.45) | 23 (125) | 0.184 (0.120,0.263) |
| | 10-12 | 103 (347) | 0.30 (0.25,0.35) | 103 (318) | 0.32 (0.27,0.38) | 15 (103) | 0.146 (0.084,0.229) |
| | 13-15 | 100 (359) | 0.28 (0.23,0.33) | 100 (327) | 0.31 (0.26,0.36) | 18 (100) | 0.180 (0.110,0.269) |
| | 16-18 | 64 (289) | 0.22 (0.17,0.27) | 64 (270) | 0.24 (0.19,0.29) | 3 (64) | 0.047 (0.010,0.131) |
| | 19-21 | 58 (281) | 0.21 (0.16,0.26) | 58 (261) | 0.22 (0.17,0.28) | 4 (58) | 0.069 (0.019,0.167) |
| | 22-24 | 48 (256) | 0.19 (0.14,0.24) | 48 (242) | 0.20 (0.15,0.25) | 1 (48) | 0.021 (0.001,0.111) |
| Astrovirus | 1-3 | 54 (152) | 0.36 (0.28,0.44) | 54 (148) | 0.36 (0.29,0.45) | 15 (54) | 0.278 (0.165,0.416) |
| | 4-6 | 77 (234) | 0.33 (0.27,0.39) | 77 (217) | 0.35 (0.29,0.42) | 14 (77) | 0.182 (0.103,0.286) |
| | 7-9 | 99 (284) | 0.35 (0.29,0.41) | 99 (269) | 0.37 (0.31,0.43) | 29 (99) | 0.293 (0.206,0.393) |
| | 10-12 | 70 (288) | 0.24 (0.19,0.30) | 70 (264) | 0.27 (0.21,0.32) | 11 (70) | 0.157 (0.081,0.264) |
| | 13-15 | 90 (297) | 0.30 (0.25,0.36) | 90 (273) | 0.33 (0.27,0.39) | 9 (90) | 0.100 (0.047,0.181) |
| | 16-18 | 65 (267) | 0.24 (0.19,0.30) | 65 (250) | 0.26 (0.21,0.32) | 9 (65) | 0.138 (0.065,0.247) |
| | 19-21 | 50 (200) | 0.25 (0.19,0.32) | 50 (184) | 0.27 (0.21,0.34) | 3 (50) | 0.060 (0.013,0.165) |
| | 22-24 | 57 (209) | 0.27 (0.21,0.34) | 57 (197) | 0.29 (0.23,0.36) | 3 (57) | 0.053 (0.011,0.146) |
| *Campylobacter jejuni* and *coli* | 1-3 | 29 (171) | 0.17 (0.12,0.23) | 29 (155) | 0.19 (0.13,0.26) | 1 (29) | 0.034 (0.001,0.178) |

| | | | | | | | |
|---|---|---|---|---|---|---|---|
| | 4-6 | 76<br>(419) | 0.18<br>(0.15,0.22) | 76<br>(333) | 0.23<br>(0.18,0.28) | 8<br>(76) | 0.105<br>(0.047,0.197) |
| | 7-9 | 113<br>(658) | 0.17<br>(0.14,0.20) | 113<br>(517) | 0.22<br>(0.18,0.26) | 26<br>(113) | 0.230<br>(0.156,0.319) |
| | 10-12 | 91<br>(706) | 0.13<br>(0.11,0.16) | 91<br>(520) | 0.17<br>(0.14,0.21) | 14<br>(91) | 0.154<br>(0.087,0.245) |
| | 13-15 | 70<br>(586) | 0.12<br>(0.09,0.15) | 70<br>(446) | 0.16<br>(0.12,0.19) | 7<br>(70) | 0.100<br>(0.041,0.195) |
| | 16-18 | 57<br>(504) | 0.11<br>(0.09,0.14) | 57<br>(385) | 0.15<br>(0.11,0.19) | 3<br>(57) | 0.053<br>(0.011,0.146) |
| | 19-21 | 33<br>(421) | 0.08<br>(0.05,0.11) | 33<br>(309) | 0.11<br>(0.07,0.15) | 2<br>(33) | 0.061<br>(0.007,0.202) |
| | 22-24 | 33<br>(366) | 0.09<br>(0.06,0.12) | 33<br>(273) | 0.12<br>(0.08,0.17) | 0<br>(33) | 0.000<br>(0.000,0.106) |
| *Cryptosporidium* | 1-3 | 3<br>(20) | 0.15<br>(0.03,0.38) | 3<br>(19) | 0.16<br>(0.03,0.40) | 1<br>(3) | 0.333<br>(0.008,0.906) |
| | 4-6 | 15<br>(69) | 0.22<br>(0.13,0.33) | 15<br>(61) | 0.25<br>(0.14,0.37) | 4<br>(15) | 0.267<br>(0.078,0.551) |
| | 7-9 | 29<br>(156) | 0.19<br>(0.13,0.26) | 29<br>(132) | 0.22<br>(0.15,0.30) | 6<br>(29) | 0.207<br>(0.080,0.397) |
| | 10-12 | 37<br>(241) | 0.15<br>(0.11,0.21) | 37<br>(205) | 0.18<br>(0.13,0.24) | 4<br>(37) | 0.108<br>(0.030,0.254) |
| | 13-15 | 44<br>(293) | 0.15<br>(0.11,0.20) | 44<br>(242) | 0.18<br>(0.14,0.24) | 7<br>(44) | 0.159<br>(0.066,0.301) |
| | 16-18 | 40<br>(296) | 0.14<br>(0.10,0.18) | 40<br>(263) | 0.15<br>(0.11,0.20) | 6<br>(40) | 0.150<br>(0.057,0.298) |
| | 19-21 | 34<br>(248) | 0.14<br>(0.10,0.19) | 34<br>(222) | 0.15<br>(0.11,0.21) | 4<br>(34) | 0.118<br>(0.033,0.275) |
| | 22-24 | 43<br>(252) | 0.17<br>(0.13,0.22) | 43<br>(220) | 0.20<br>(0.15,0.25) | 1<br>(43) | 0.023<br>(0.001,0.123) |
| Norovirus GII | 1-3 | 31<br>(179) | 0.17<br>(0.12,0.24) | 31<br>(162) | 0.19<br>(0.13,0.26) | 7<br>(31) | 0.226<br>(0.096,0.411) |
| | 4-6 | 75<br>(483) | 0.16<br>(0.12,0.19) | 75<br>(414) | 0.18<br>(0.15,0.22) | 19<br>(75) | 0.253<br>(0.160,0.367) |
| | 7-9 | 142<br>(588) | 0.24<br>(0.21,0.28) | 142<br>(507) | 0.28<br>(0.24,0.32) | 23<br>(142) | 0.162<br>(0.106,0.233) |
| | 10-12 | 112<br>(472) | 0.24<br>(0.20,0.28) | 112<br>(424) | 0.26<br>(0.22,0.31) | 12<br>(112) | 0.107<br>(0.057,0.180) |
| | 13-15 | 92<br>(348) | 0.26<br>(0.22,0.31) | 92<br>(324) | 0.28<br>(0.24,0.34) | 18<br>(92) | 0.196<br>(0.120,0.291) |
| | 16-18 | 60<br>(289) | 0.21<br>(0.16,0.26) | 60<br>(270) | 0.22<br>(0.17,0.28) | 11<br>(60) | 0.183<br>(0.095,0.304) |
| | 19-21 | 36<br>(214) | 0.17<br>(0.12,0.23) | 36<br>(202) | 0.18<br>(0.13,0.24) | 3<br>(36) | 0.083<br>(0.018,0.225) |
| | 22-24 | 31<br>(155) | 0.20<br>(0.14,0.27) | 31<br>(146) | 0.21<br>(0.15,0.29) | 2<br>(31) | 0.065<br>(0.008,0.214) |

| | | | | | | | |
|---|---|---|---|---|---|---|---|
| Rotavirus | 1-3 | 37<br>(74) | 0.50<br>(0.38,0.62) | 37<br>(73) | 0.51<br>(0.39,0.63) | 14<br>(37) | 0.378<br>(0.225,0.552) |
| | 4-6 | 72<br>(119) | 0.61<br>(0.51,0.69) | 72<br>(114) | 0.63<br>(0.54,0.72) | 27<br>(72) | 0.375<br>(0.264,0.497) |
| | 7-9 | 104<br>(144) | 0.72<br>(0.64,0.79) | 104<br>(142) | 0.73<br>(0.65,0.80) | 39<br>(104) | 0.375<br>(0.282,0.475) |
| | 10-12 | 98<br>(143) | 0.69<br>(0.60,0.76) | 98<br>(138) | 0.71<br>(0.63,0.78) | 26<br>(98) | 0.265<br>(0.181,0.364) |
| | 13-15 | 71<br>(109) | 0.65<br>(0.55,0.74) | 71<br>(108) | 0.66<br>(0.56,0.75) | 22<br>(71) | 0.310<br>(0.205,0.431) |
| | 16-18 | 40<br>(61) | 0.66<br>(0.52,0.77) | 40<br>(58) | 0.69<br>(0.55,0.80) | 9<br>(40) | 0.225<br>(0.108,0.385) |
| | 19-21 | 29<br>(59) | 0.49<br>(0.36,0.63) | 29<br>(59) | 0.49<br>(0.36,0.63) | 3<br>(29) | 0.103<br>(0.022,0.274) |
| | 22-24 | 21<br>(42) | 0.50<br>(0.34,0.66) | 21<br>(42) | 0.50<br>(0.34,0.66) | 2<br>(21) | 0.095<br>(0.012,0.304) |
| Sapovirus | 1-3 | 13<br>(71) | 0.18<br>(0.10,0.29) | 13<br>(67) | 0.19<br>(0.11,0.31) | 6<br>(13) | 0.462<br>(0.192,0.749) |
| | 4-6 | 66<br>(261) | 0.25<br>(0.20,0.31) | 66<br>(241) | 0.27<br>(0.22,0.33) | 17<br>(66) | 0.258<br>(0.158,0.380) |
| | 7-9 | 128<br>(446) | 0.29<br>(0.25,0.33) | 128<br>(396) | 0.32<br>(0.28,0.37) | 28<br>(128) | 0.219<br>(0.151,0.300) |
| | 10-12 | 139<br>(532) | 0.26<br>(0.22,0.30) | 139<br>(460) | 0.30<br>(0.26,0.35) | 22<br>(139) | 0.158<br>(0.102,0.230) |
| | 13-15 | 157<br>(519) | 0.30<br>(0.26,0.34) | 157<br>(456) | 0.34<br>(0.30,0.39) | 22<br>(157) | 0.140<br>(0.090,0.204) |
| | 16-18 | 109<br>(459) | 0.24<br>(0.20,0.28) | 109<br>(423) | 0.26<br>(0.22,0.30) | 17<br>(109) | 0.156<br>(0.094,0.238) |
| | 19-21 | 86<br>(381) | 0.23<br>(0.18,0.27) | 86<br>(337) | 0.26<br>(0.21,0.31) | 10<br>(86) | 0.116<br>(0.057,0.203) |
| | 22-24 | 59<br>(324) | 0.18<br>(0.14,0.23) | 59<br>(303) | 0.19<br>(0.15,0.24) | 3<br>(59) | 0.051<br>(0.011,0.141) |
| *Shigella* | 1-3 | 6<br>(25) | 0.24<br>(0.09,0.45) | 6<br>(24) | 0.25<br>(0.10,0.47) | 2<br>(6) | 0.333<br>(0.043,0.777) |
| | 4-6 | 21<br>(60) | 0.35<br>(0.23,0.48) | 21<br>(57) | 0.37<br>(0.24,0.51) | 6<br>(21) | 0.286<br>(0.113,0.522) |
| | 7-9 | 54<br>(193) | 0.28<br>(0.22,0.35) | 54<br>(163) | 0.33<br>(0.26,0.41) | 12<br>(54) | 0.222<br>(0.120,0.356) |
| | 10-12 | 96<br>(336) | 0.29<br>(0.24,0.34) | 96<br>(265) | 0.36<br>(0.30,0.42) | 12<br>(96) | 0.125<br>(0.066,0.208) |
| | 13-15 | 133<br>(392) | 0.34<br>(0.29,0.39) | 133<br>(328) | 0.41<br>(0.35,0.46) | 18<br>(133) | 0.135<br>(0.082,0.205) |
| | 16-18 | 151<br>(487) | 0.31<br>(0.27,0.35) | 151<br>(408) | 0.37<br>(0.32,0.42) | 26<br>(151) | 0.172<br>(0.116,0.242) |
| | 19-21 | 166<br>(599) | 0.28<br>(0.24,0.31) | 166<br>(461) | 0.36<br>(0.32,0.41) | 13<br>(166) | 0.078<br>(0.042,0.130) |

| | | | | | | | |
|---|---|---|---|---|---|---|---|
| | 22-24 | 143<br>(564) | 0.25<br>(0.22,0.29) | 143<br>(433) | 0.33<br>(0.29,0.38) | 13<br>(143) | 0.091<br>(0.049,0.150) |
| ST-ETEC | 1-3 | 29<br>(109) | 0.27<br>(0.19,0.36) | 29<br>(103) | 0.28<br>(0.20,0.38) | 8<br>(29) | 0.276<br>(0.127,0.472) |
| | 4-6 | 58<br>(266) | 0.22<br>(0.17,0.27) | 58<br>(248) | 0.23<br>(0.18,0.29) | 15<br>(58) | 0.259<br>(0.153,0.390) |
| | 7-9 | 97<br>(422) | 0.23<br>(0.19,0.27) | 97<br>(371) | 0.26<br>(0.22,0.31) | 20<br>(97) | 0.206<br>(0.131,0.300) |
| | 10-12 | 118<br>(555) | 0.21<br>(0.18,0.25) | 118<br>(496) | 0.24<br>(0.20,0.28) | 12<br>(118) | 0.102<br>(0.054,0.171) |
| | 13-15 | 103<br>(527) | 0.20<br>(0.16,0.23) | 103<br>(446) | 0.23<br>(0.19,0.27) | 16<br>(103) | 0.155<br>(0.091,0.240) |
| | 16-18 | 108<br>(508) | 0.21<br>(0.18,0.25) | 108<br>(447) | 0.24<br>(0.20,0.28) | 16<br>(108) | 0.148<br>(0.087,0.229) |
| | 19-21 | 85<br>(509) | 0.17<br>(0.14,0.20) | 85<br>(434) | 0.20<br>(0.16,0.24) | 10<br>(85) | 0.118<br>(0.058,0.206) |
| | 22-24 | 73<br>(456) | 0.16<br>(0.13,0.20) | 73<br>(407) | 0.18<br>(0.14,0.22) | 3<br>(73) | 0.041<br>(0.009,0.115) |
| tEPEC | 1-3 | 37<br>(160) | 0.23<br>(0.17,0.30) | 37<br>(141) | 0.26<br>(0.19,0.34) | 11<br>(37) | 0.297<br>(0.159,0.470) |
| | 4-6 | 61<br>(381) | 0.16<br>(0.12,0.20) | 61<br>(315) | 0.19<br>(0.15,0.24) | 17<br>(61) | 0.279<br>(0.171,0.408) |
| | 7-9 | 49<br>(560) | 0.09<br>(0.07,0.11) | 49<br>(449) | 0.11<br>(0.08,0.14) | 10<br>(49) | 0.204<br>(0.102,0.343) |
| | 10-12 | 50<br>(496) | 0.10<br>(0.08,0.13) | 50<br>(406) | 0.12<br>(0.09,0.16) | 7<br>(50) | 0.140<br>(0.058,0.267) |
| | 13-15 | 29<br>(428) | 0.07<br>(0.05,0.10) | 29<br>(366) | 0.08<br>(0.05,0.11) | 3<br>(29) | 0.103<br>(0.022,0.274) |
| | 16-18 | 29<br>(356) | 0.08<br>(0.06,0.11) | 29<br>(317) | 0.09<br>(0.06,0.13) | 5<br>(29) | 0.172<br>(0.058,0.358) |
| | 19-21 | 24<br>(324) | 0.07<br>(0.05,0.11) | 24<br>(298) | 0.08<br>(0.05,0.12) | 1<br>(24) | 0.042<br>(0.001,0.211) |
| | 22-24 | 18<br>(279) | 0.06<br>(0.04,0.10) | 18<br>(250) | 0.07<br>(0.04,0.11) | 1<br>(18) | 0.056<br>(0.001,0.273) |

**Supplemental Table 17. Proportion of infections that were pathogen-attributable diarrhea and proportion of pathogen-attributable diarrheal episodes that were severe (score >6) and exact 95% confidence intervals (95% CIs) by 3-month age bands and infection history using algorithm attribution and the consecutive detections definition for unique infections.** Algorithm attribution considered both pathogen-attributable fraction and longitudinal changes in the quantity of pathogen detected. The consecutive detections definition considered both consecutive positive detections and time between consecutive positives.

| Pathogen | Age (months) | Proportion of infections that were pathogen-attributable diarrhea | | | | Proportion of pathogen-attributable diarrheal episodes that were severe | | | |
|---|---|---|---|---|---|---|---|---|---|
| | | No prior infections | | ≥1 prior infection | | No prior infections | | ≥1 prior infection | |
| | | Diarrhea (infections) | Proportion (95% CI) | *Diarrhea (infections)* | *Proportion (95% CI)* | Severe diarrhea (diarrhea) | Proportion (95% CI) | Severe diarrhea (diarrhea) | Proportion (95% CI) |
| Adenovirus 40/41 | 1-3 | 40 (120) | 0.33 (0.25,0.43) | 3 (4) | 0.75 (0.19,0.99) | 4 (40) | 0.100 (0.028,0.237) | 1 (3) | 0.333 (0.008,0.906) |
| | 4-6 | 79 (214) | 0.37 (0.30,0.44) | 20 (38) | 0.53 (0.36,0.69) | 10 (79) | 0.127 (0.062,0.220) | 3 (20) | 0.150 (0.032,0.379) |
| | 7-9 | 67 (198) | 0.34 (0.27,0.41) | 58 (122) | 0.48 (0.38,0.57) | 11 (67) | 0.164 (0.085,0.275) | 12 (58) | 0.207 (0.112,0.334) |
| | 10-12 | 49 (182) | 0.27 (0.21,0.34) | 54 (136) | 0.40 (0.31,0.48) | 8 (49) | 0.163 (0.073,0.297) | 7 (54) | 0.130 (0.054,0.249) |
| | 13-15 | 34 (172) | 0.20 (0.14,0.27) | 66 (155) | 0.43 (0.35,0.51) | 4 (34) | 0.118 (0.033,0.275) | 14 (66) | 0.212 (0.121,0.330) |
| | 16-18 | 14 (108) | 0.13 (0.07,0.21) | 50 (162) | 0.31 (0.24,0.39) | 2 (14) | 0.143 (0.018,0.428) | 1 (50) | 0.020 (0.001,0.106) |
| | 19-21 | 8 (87) | 0.09 (0.04,0.17) | 50 (174) | 0.29 (0.22,0.36) | 1 (8) | 0.125 (0.003,0.527) | 3 (50) | 0.060 (0.013,0.165) |
| | 22-24 | 6 (63) | 0.10 (0.04,0.20) | 42 (179) | 0.23 (0.17,0.30) | 0 (6) | 0.000 (0.000,0.459) | 1 (42) | 0.024 (0.001,0.126) |
| Astrovirus | 1-3 | 50 (138) | 0.36 (0.28,0.45) | 4 (10) | 0.40 (0.12,0.74) | 15 (50) | 0.300 (0.179,0.446) | 0 (4) | 0.000 (0.000,0.602) |
| | 4-6 | 63 (194) | 0.32 (0.26,0.40) | 14 (23) | 0.61 (0.39,0.80) | 12 (63) | 0.190 (0.102,0.309) | 2 (14) | 0.143 (0.018,0.428) |
| | 7-9 | 70 (204) | 0.34 (0.28,0.41) | 29 (65) | 0.45 (0.32,0.57) | 22 (70) | 0.314 (0.209,0.436) | 7 (29) | 0.241 (0.103,0.435) |
| | 10-12 | 36 (165) | 0.22 (0.16,0.29) | 34 (99) | 0.34 (0.25,0.45) | 4 (36) | 0.111 (0.031,0.261) | 7 (34) | 0.206 (0.087,0.379) |
| | 13-15 | 39 (147) | 0.27 (0.20,0.34) | 51 (126) | 0.40 (0.32,0.50) | 6 (39) | 0.154 (0.059,0.305) | 3 (51) | 0.059 (0.012,0.162) |
| | 16-18 | 22 (96) | 0.23 (0.15,0.33) | 43 (154) | 0.28 (0.21,0.36) | 2 (22) | 0.091 (0.011,0.292) | 7 (43) | 0.163 (0.068,0.307) |
| | 19-21 | 11 (67) | 0.16 (0.08,0.27) | 39 (117) | 0.33 (0.25,0.43) | 1 (11) | 0.091 (0.002,0.413) | 2 (39) | 0.051 (0.006,0.173) |
| | 22-24 | 13 (53) | 0.25 (0.14,0.38) | 44 (144) | 0.31 (0.23,0.39) | 1 (13) | 0.077 (0.002,0.360) | 2 (44) | 0.045 (0.006,0.155) |
| *Campylobacter jejuni* and *coli* | 1-3 | 28 (153) | 0.18 (0.13,0.25) | 1 (2) | 0.50 (0.01,0.99) | 1 (28) | 0.036 (0.001,0.183) | 0 (1) | 0.000 (0.000,0.975) |
| | 4-6 | 58 (273) | 0.21 (0.17,0.27) | 18 (60) | 0.30 (0.19,0.43) | 8 (58) | 0.138 (0.061,0.254) | 0 (18) | 0.000 (0.000,0.185) |

| | | | | | | | | | |
|---|---|---|---|---|---|---|---|---|---|
| | 7-9 | 76 (331) | 0.23 (0.19,0.28) | 37 (186) | 0.20 (0.14,0.26) | 15 (76) | 0.197 (0.115,0.305) | 11 (37) | 0.297 (0.159,0.470) |
| | 10-12 | 33 (203) | 0.16 (0.11,0.22) | 58 (317) | 0.18 (0.14,0.23) | 4 (33) | 0.121 (0.034,0.282) | 10 (58) | 0.172 (0.086,0.294) |
| | 13-15 | 17 (116) | 0.15 (0.09,0.22) | 53 (330) | 0.16 (0.12,0.20) | 3 (17) | 0.176 (0.038,0.434) | 4 (53) | 0.075 (0.021,0.182) |
| | 16-18 | 14 (62) | 0.23 (0.13,0.35) | 43 (323) | 0.13 (0.10,0.18) | 0 (14) | 0.000 (0.000,0.232) | 3 (43) | 0.070 (0.015,0.191) |
| | 19-21 | 6 (37) | 0.16 (0.06,0.32) | 27 (272) | 0.10 (0.07,0.14) | 1 (6) | 0.167 (0.004,0.641) | 1 (27) | 0.037 (0.001,0.190) |
| | 22-24 | 2 (27) | 0.07 (0.01,0.24) | 31 (246) | 0.13 (0.09,0.17) | 0 (2) | 0.000 (0.000,0.842) | 0 (31) | 0.000 (0.000,0.112) |
| *Cryptosporidium* | 1-3 | 3 (19) | 0.16 (0.03,0.40) | NA | NA | 1 (3) | 0.333 (0.008,0.906) | NA | NA |
| | 4-6 | 15 (58) | 0.26 (0.15,0.39) | 0 (3) | 0.00 (0.00,0.71) | 4 (15) | 0.267 (0.078,0.551) | NA | NA |
| | 7-9 | 24 (118) | 0.20 (0.13,0.29) | 5 (14) | 0.36 (0.13,0.65) | 6 (24) | 0.250 (0.098,0.467) | 0 (5) | 0.000 (0.000,0.522) |
| | 10-12 | 24 (165) | 0.15 (0.10,0.21) | 13 (40) | 0.32 (0.19,0.49) | 3 (24) | 0.125 (0.027,0.324) | 1 (13) | 0.077 (0.002,0.360) |
| | 13-15 | 30 (181) | 0.17 (0.11,0.23) | 14 (61) | 0.23 (0.13,0.35) | 6 (30) | 0.200 (0.077,0.386) | 1 (14) | 0.071 (0.002,0.339) |
| | 16-18 | 26 (166) | 0.16 (0.10,0.22) | 14 (97) | 0.14 (0.08,0.23) | 4 (26) | 0.154 (0.044,0.349) | 2 (14) | 0.143 (0.018,0.428) |
| | 19-21 | 19 (128) | 0.15 (0.09,0.22) | 15 (94) | 0.16 (0.09,0.25) | 1 (19) | 0.053 (0.001,0.260) | 3 (15) | 0.200 (0.043,0.481) |
| | 22-24 | 21 (118) | 0.18 (0.11,0.26) | 22 (102) | 0.22 (0.14,0.31) | 1 (21) | 0.048 (0.001,0.238) | 0 (22) | 0.000 (0.000,0.154) |
| Norovirus GII | 1-3 | 29 (156) | 0.19 (0.13,0.26) | 2 (6) | 0.33 (0.04,0.78) | 6 (29) | 0.207 (0.080,0.397) | 1 (2) | 0.500 (0.013,0.987) |
| | 4-6 | 55 (356) | 0.15 (0.12,0.20) | 20 (58) | 0.34 (0.22,0.48) | 14 (55) | 0.255 (0.147,0.390) | 5 (20) | 0.250 (0.087,0.491) |
| | 7-9 | 91 (334) | 0.27 (0.23,0.32) | 51 (173) | 0.29 (0.23,0.37) | 16 (91) | 0.176 (0.104,0.270) | 7 (51) | 0.137 (0.057,0.263) |
| | 10-12 | 49 (188) | 0.26 (0.20,0.33) | 63 (236) | 0.27 (0.21,0.33) | 5 (49) | 0.102 (0.034,0.222) | 7 (63) | 0.111 (0.046,0.216) |
| | 13-15 | 27 (100) | 0.27 (0.19,0.37) | 65 (224) | 0.29 (0.23,0.35) | 4 (27) | 0.148 (0.042,0.337) | 14 (65) | 0.215 (0.123,0.335) |
| | 16-18 | 22 (89) | 0.25 (0.16,0.35) | 38 (181) | 0.21 (0.15,0.28) | 5 (22) | 0.227 (0.078,0.454) | 6 (38) | 0.158 (0.060,0.313) |
| | 19-21 | 4 (46) | 0.09 (0.02,0.21) | 32 (156) | 0.21 (0.14,0.28) | 2 (4) | 0.500 (0.068,0.932) | 1 (32) | 0.031 (0.001,0.162) |
| | 22-24 | 5 (23) | 0.22 (0.07,0.44) | 26 (123) | 0.21 (0.14,0.29) | 0 (5) | 0.000 (0.000,0.522) | 2 (26) | 0.077 (0.009,0.251) |
| Rotavirus | 1-3 | 35 (70) | 0.50 (0.38,0.62) | 2 (3) | 0.67 (0.09,0.99) | 14 (35) | 0.400 (0.239,0.579) | 0 (2) | 0.000 (0.000,0.842) |

| | | | | | | | | | |
|---|---|---|---|---|---|---|---|---|---|
| | 4-6 | 66 (106) | 0.62 (0.52,0.71) | 6 (8) | 0.75 (0.35,0.97) | 26 (66) | 0.394 (0.276,0.522) | 1 (6) | 0.167 (0.004,0.641) |
| | 7-9 | 84 (119) | 0.71 (0.62,0.79) | 20 (23) | 0.87 (0.66,0.97) | 31 (84) | 0.369 (0.266,0.481) | 8 (20) | 0.400 (0.191,0.639) |
| | 10-12 | 71 (102) | 0.70 (0.60,0.78) | 27 (36) | 0.75 (0.58,0.88) | 23 (71) | 0.324 (0.218,0.445) | 3 (27) | 0.111 (0.024,0.292) |
| | 13-15 | 50 (74) | 0.68 (0.56,0.78) | 21 (34) | 0.62 (0.44,0.78) | 18 (50) | 0.360 (0.229,0.508) | 4 (21) | 0.190 (0.054,0.419) |
| | 16-18 | 20 (29) | 0.69 (0.49,0.85) | 20 (29) | 0.69 (0.49,0.85) | 6 (20) | 0.300 (0.119,0.543) | 3 (20) | 0.150 (0.032,0.379) |
| | 19-21 | 9 (25) | 0.36 (0.18,0.57) | 20 (34) | 0.59 (0.41,0.75) | 1 (9) | 0.111 (0.003,0.482) | 2 (20) | 0.100 (0.012,0.317) |
| | 22-24 | 11 (21) | 0.52 (0.30,0.74) | 10 (21) | 0.48 (0.26,0.70) | 1 (11) | 0.091 (0.002,0.413) | 1 (10) | 0.100 (0.003,0.445) |
| Sapovirus | 1-3 | 10 (63) | 0.16 (0.08,0.27) | 3 (4) | 0.75 (0.19,0.99) | 5 (10) | 0.500 (0.187,0.813) | 1 (3) | 0.333 (0.008,0.906) |
| | 4-6 | 58 (229) | 0.25 (0.20,0.31) | 8 (12) | 0.67 (0.35,0.90) | 13 (58) | 0.224 (0.125,0.353) | 4 (8) | 0.500 (0.157,0.843) |
| | 7-9 | 91 (304) | 0.30 (0.25,0.35) | 37 (92) | 0.40 (0.30,0.51) | 18 (91) | 0.198 (0.122,0.294) | 10 (37) | 0.270 (0.138,0.441) |
| | 10-12 | 81 (271) | 0.30 (0.25,0.36) | 58 (189) | 0.31 (0.24,0.38) | 14 (81) | 0.173 (0.098,0.273) | 8 (58) | 0.138 (0.061,0.254) |
| | 13-15 | 60 (201) | 0.30 (0.24,0.37) | 97 (255) | 0.38 (0.32,0.44) | 7 (60) | 0.117 (0.048,0.226) | 15 (97) | 0.155 (0.089,0.242) |
| | 16-18 | 21 (139) | 0.15 (0.10,0.22) | 88 (284) | 0.31 (0.26,0.37) | 3 (21) | 0.143 (0.030,0.363) | 14 (88) | 0.159 (0.090,0.252) |
| | 19-21 | 13 (90) | 0.14 (0.08,0.23) | 73 (247) | 0.30 (0.24,0.36) | 1 (13) | 0.077 (0.002,0.360) | 9 (73) | 0.123 (0.058,0.221) |
| | 22-24 | 11 (62) | 0.18 (0.09,0.30) | 48 (241) | 0.20 (0.15,0.26) | 0 (11) | 0.000 (0.000,0.285) | 3 (48) | 0.062 (0.013,0.172) |
| *Shigella* | 1-3 | 5 (23) | 0.22 (0.07,0.44) | 1 (1) | 1.00 (0.03,1.00) | 2 (5) | 0.400 (0.053,0.853) | 0 (1) | 0.000 (0.000,0.975) |
| | 4-6 | 20 (51) | 0.39 (0.26,0.54) | 1 (6) | 0.17 (0.00,0.64) | 6 (20) | 0.300 (0.119,0.543) | 0 (1) | 0.000 (0.000,0.975) |
| | 7-9 | 49 (139) | 0.35 (0.27,0.44) | 5 (24) | 0.21 (0.07,0.42) | 11 (49) | 0.224 (0.118,0.366) | 1 (5) | 0.200 (0.005,0.716) |
| | 10-12 | 71 (201) | 0.35 (0.29,0.42) | 25 (64) | 0.39 (0.27,0.52) | 10 (71) | 0.141 (0.070,0.244) | 2 (25) | 0.080 (0.010,0.260) |
| | 13-15 | 81 (197) | 0.41 (0.34,0.48) | 52 (131) | 0.40 (0.31,0.49) | 10 (81) | 0.123 (0.061,0.215) | 8 (52) | 0.154 (0.069,0.281) |
| | 16-18 | 92 (221) | 0.42 (0.35,0.48) | 59 (187) | 0.32 (0.25,0.39) | 20 (92) | 0.217 (0.138,0.316) | 6 (59) | 0.102 (0.038,0.208) |
| | 19-21 | 68 (172) | 0.40 (0.32,0.47) | 98 (289) | 0.34 (0.28,0.40) | 7 (68) | 0.103 (0.042,0.201) | 6 (98) | 0.061 (0.023,0.129) |
| | 22-24 | 34 (112) | 0.30 (0.22,0.40) | 109 (321) | 0.34 (0.29,0.39) | 4 (34) | 0.118 (0.033,0.275) | 9 (109) | 0.083 (0.038,0.151) |

| | | | | | | | | | |
|---|---|---|---|---|---|---|---|---|---|
| ST-ETEC | 1-3 | 27<br>(99) | 0.27<br>(0.19,0.37) | 2<br>(4) | 0.50<br>(0.07,0.93) | 7<br>(27) | 0.259<br>(0.111,0.463) | 1<br>(2) | 0.500<br>(0.013,0.987) |
| | 4-6 | 43<br>(207) | 0.21<br>(0.15,0.27) | 15<br>(41) | 0.37<br>(0.22,0.53) | 11<br>(43) | 0.256<br>(0.135,0.412) | 4<br>(15) | 0.267<br>(0.078,0.551) |
| | 7-9 | 57<br>(259) | 0.22<br>(0.17,0.28) | 40<br>(112) | 0.36<br>(0.27,0.45) | 14<br>(57) | 0.246<br>(0.141,0.378) | 6<br>(40) | 0.150<br>(0.057,0.298) |
| | 10-12 | 49<br>(247) | 0.20<br>(0.15,0.25) | 69<br>(249) | 0.28<br>(0.22,0.34) | 6<br>(49) | 0.122<br>(0.046,0.248) | 6<br>(69) | 0.087<br>(0.033,0.180) |
| | 13-15 | 25<br>(167) | 0.15<br>(0.10,0.21) | 78<br>(279) | 0.28<br>(0.23,0.34) | 4<br>(25) | 0.160<br>(0.045,0.361) | 12<br>(78) | 0.154<br>(0.082,0.253) |
| | 16-18 | 16<br>(117) | 0.14<br>(0.08,0.21) | 92<br>(330) | 0.28<br>(0.23,0.33) | 5<br>(16) | 0.312<br>(0.110,0.587) | 11<br>(92) | 0.120<br>(0.061,0.204) |
| | 19-21 | 15<br>(98) | 0.15<br>(0.09,0.24) | 70<br>(336) | 0.21<br>(0.17,0.26) | 4<br>(15) | 0.267<br>(0.078,0.551) | 6<br>(70) | 0.086<br>(0.032,0.177) |
| | 22-24 | 9<br>(56) | 0.16<br>(0.08,0.28) | 64<br>(351) | 0.18<br>(0.14,0.23) | 1<br>(9) | 0.111<br>(0.003,0.482) | 2<br>(64) | 0.031<br>(0.004,0.108) |
| tEPEC | 1-3 | 31<br>(133) | 0.23<br>(0.16,0.31) | 6<br>(8) | 0.75<br>(0.35,0.97) | 10<br>(31) | 0.323<br>(0.167,0.514) | 1<br>(6) | 0.167<br>(0.004,0.641) |
| | 4-6 | 47<br>(268) | 0.18<br>(0.13,0.23) | 14<br>(47) | 0.30<br>(0.17,0.45) | 13<br>(47) | 0.277<br>(0.156,0.426) | 4<br>(14) | 0.286<br>(0.084,0.581) |
| | 7-9 | 34<br>(320) | 0.11<br>(0.07,0.15) | 15<br>(129) | 0.12<br>(0.07,0.18) | 7<br>(34) | 0.206<br>(0.087,0.379) | 3<br>(15) | 0.200<br>(0.043,0.481) |
| | 10-12 | 25<br>(219) | 0.11<br>(0.08,0.16) | 25<br>(187) | 0.13<br>(0.09,0.19) | 4<br>(25) | 0.160<br>(0.045,0.361) | 3<br>(25) | 0.120<br>(0.025,0.312) |
| | 13-15 | 8<br>(130) | 0.06<br>(0.03,0.12) | 21<br>(236) | 0.09<br>(0.06,0.13) | 3<br>(8) | 0.375<br>(0.085,0.755) | 0<br>(21) | 0.000<br>(0.000,0.161) |
| | 16-18 | 9<br>(82) | 0.11<br>(0.05,0.20) | 20<br>(235) | 0.09<br>(0.05,0.13) | 2<br>(9) | 0.222<br>(0.028,0.600) | 3<br>(20) | 0.150<br>(0.032,0.379) |
| | 19-21 | 7<br>(60) | 0.12<br>(0.05,0.23) | 17<br>(238) | 0.07<br>(0.04,0.11) | 1<br>(7) | 0.143<br>(0.004,0.579) | 0<br>(17) | 0.000<br>(0.000,0.195) |
| | 22-24 | 3<br>(44) | 0.07<br>(0.01,0.19) | 15<br>(206) | 0.07<br>(0.04,0.12) | 0<br>(3) | 0.000<br>(0.000,0.708) | 1<br>(15) | 0.067<br>(0.002,0.319) |

**Supplemental Table 18. Median and interquartile range for duration of infection by 3-month age bands (unstratified or stratified by infection history) using algorithm attribution and the consecutive detections definition for unique infections.** Algorithm attribution considered both pathogen-attributable fraction and longitudinal changes in the quantity of pathogen detected. The consecutive detections definition considered both consecutive positive detections and time between consecutive positives.

| Pathogen | Age (months) | Median duration in months (interquartile range) | | |
|---|---|---|---|---|
| | | Unstratified | Stratified by infection history | |
| | | | No prior infections | ≥1 prior infection |
| Adenovirus 40/41 | 1-3 | 1 (1,1) | 1 (1,1) | 1 (1,1) |
| | 4-6 | 1 (1,1) | 1 (1,1) | 1 (0,1) |
| | 7-9 | 1 (1,1) | 1 (1,1) | 1 (1,1) |
| | 10-12 | 1 (1,1) | 1 (1,1) | 1 (1,1) |
| | 13-15 | 1 (1,1) | 1 (1,1) | 1 (1,1) |
| | 16-18 | 1 (1,1) | 1 (1,1) | 1 (1,1) |
| | 19-21 | 1 (1,1) | 1 (1,1) | 1 (1,1) |
| | 22-24 | 1 (1,1) | 1 (1,1) | 1 (1,1) |
| Astrovirus | 1-3 | 1 (1,1) | 1 (1,1) | 1 (1,1) |
| | 4-6 | 1 (1,1) | 1 (1,1) | 1 (1,1) |
| | 7-9 | 1 (1,1) | 1 (1,1) | 1 (1,1) |
| | 10-12 | 1 (1,1) | 1 (1,1) | 1 (1,1) |
| | 13-15 | 1 (1,1) | 1 (1,1) | 1 (1,1) |
| | 16-18 | 1 (1,1) | 1 (1,1) | 1 (1,1) |
| | 19-21 | 1 (1,1) | 1 (1,1) | 1 (1,1) |
| | 22-24 | 1 (1,1) | 1 (1,1) | 1 (1,1) |
| *Campylobacter jejuni* and *coli* | 1-3 | 1 (1,1) | 1 (1,1) | 1 (1,1) |
| | 4-6 | 1 (1,1) | 1 (1,1) | 1 (1,1) |
| | 7-9 | 1 (1,1) | 1 (1,1) | 1 (1,1) |
| | 10-12 | 1 (1,1) | 1 (1,1) | 1 (1,1) |
| | 13-15 | 1 (1,1) | 1 (1,1) | 1 (1,1) |
| | 16-18 | 1 (1,1) | 1 (1,1) | 1 (1,1) |
| | 19-21 | 1 (1,1) | 1 (1,1) | 1 (1,1) |
| | 22-24 | 1 (1,1) | 1 (1,1) | 1 (1,1) |
| Cryptosporidium | 1-3 | 1 (1,1) | 1 (1,1) | -- |
| | 4-6 | 1 (1,1) | 1 (1,1) | 1 (1,1) |

| | | | | |
|---|---|---|---|---|
| | 7-9 | 1 (1,1) | 1 (1,1) | 1 (1,1) |
| | 10-12 | 1 (1,1) | 1 (1,1) | 1 (1,1) |
| | 13-15 | 1 (1,1) | 1 (1,1) | 1 (1,1) |
| | 16-18 | 1 (1,1) | 1 (1,1) | 1 (1,1) |
| | 19-21 | 1 (1,1) | 1 (1,1) | 1 (1,1) |
| | 22-24 | 1 (1,1) | 1 (1,1) | 1 (1,1) |
| Norovirus GII | 1-3 | 1 (1,1) | 1 (1,1) | 1 (1,1) |
| | 4-6 | 1 (1,1) | 1 (1,1) | 1 (1,1) |
| | 7-9 | 1 (1,1) | 1 (1,1) | 1 (1,1) |
| | 10-12 | 1 (1,1) | 1 (1,1) | 1 (1,1) |
| | 13-15 | 1 (1,1) | 1 (1,1) | 1 (1,1) |
| | 16-18 | 1 (1,1) | 1 (1,1) | 1 (1,1) |
| | 19-21 | 1 (1,1) | 1 (1,1) | 1 (1,1) |
| | 22-24 | 1 (1,1) | 1 (1,1) | 1 (1,1) |
| Rotavirus | 1-3 | 1 (1,1) | 1 (1,1) | 1 (1,1) |
| | 4-6 | 1 (1,1) | 1 (1,1) | 1 (1,1) |
| | 7-9 | 1 (1,1) | 1 (0,1) | 1 (1,1) |
| | 10-12 | 1 (0,1) | 1 (1,1) | 1 (0,1) |
| | 13-15 | 1 (0,1) | 1 (0,1) | 1 (1,1) |
| | 16-18 | 1 (1,1) | 1 (1,1) | 1 (1,1) |
| | 19-21 | 1 (1,1) | 1 (1,1) | 1 (1,1) |
| | 22-24 | 1 (1,1) | 1 (1,1) | 1 (1,1) |
| Sapovirus | 1-3 | 1 (1,1) | 1 (1,1) | 1 (1,1) |
| | 4-6 | 1 (1,1) | 1 (1,1) | 1 (1,1) |
| | 7-9 | 1 (1,1) | 1 (1,1) | 1 (1,1) |
| | 10-12 | 1 (1,1) | 1 (1,1) | 1 (1,1) |
| | 13-15 | 1 (1,1) | 1 (1,1) | 1 (1,1) |
| | 16-18 | 1 (1,1) | 1 (1,1) | 1 (1,1) |
| | 19-21 | 1 (1,1) | 1 (1,1) | 1 (1,1) |
| | 22-24 | 1 (1,1) | 1 (1,1) | 1 (1,1) |
| Shigella | 1-3 | 1 (1,1) | 1 (1,1) | 0 (0,0) |
| | 4-6 | 1 (1,1) | 1 (1,1) | 1 (1,1) |

| | | | | |
|---|---|---|---|---|
| | 7-9 | 1<br>(1,1) | 1<br>(1,1) | 1<br>(1,1) |
| | 10-12 | 1<br>(1,1) | 1<br>(1,1) | 1<br>(1,1) |
| | 13-15 | 1<br>(1,1) | 1<br>(1,1) | 1<br>(1,1) |
| | 16-18 | 1<br>(1,1) | 1<br>(1,1) | 1<br>(1,1) |
| | 19-21 | 1<br>(1,1) | 1<br>(1,1) | 1<br>(1,1) |
| | 22-24 | 1<br>(1,1) | 1<br>(1,1) | 1<br>(1,1) |
| ST-ETEC | 1-3 | 1<br>(1,1) | 1<br>(1,1) | 1<br>(0,1) |
| | 4-6 | 1<br>(1,1) | 1<br>(1,1) | 1<br>(1,1) |
| | 7-9 | 1<br>(1,1) | 1<br>(1,1) | 1<br>(1,1) |
| | 10-12 | 1<br>(1,1) | 1<br>(1,1) | 1<br>(1,1) |
| | 13-15 | 1<br>(1,1) | 1<br>(1,1) | 1<br>(1,1) |
| | 16-18 | 1<br>(1,1) | 1<br>(1,1) | 1<br>(1,1) |
| | 19-21 | 1<br>(1,1) | 1<br>(1,1) | 1<br>(1,1) |
| | 22-24 | 1<br>(1,1) | 1<br>(1,1) | 1<br>(1,1) |
| tEPEC | 1-3 | 1<br>(1,1) | 1<br>(1,1) | 1<br>(0,1) |
| | 4-6 | 1<br>(1,1) | 1<br>(1,1) | 1<br>(1,1) |
| | 7-9 | 1<br>(1,1) | 1<br>(1,1) | 1<br>(1,1) |
| | 10-12 | 1<br>(1,1) | 1<br>(1,1) | 1<br>(1,1) |
| | 13-15 | 1<br>(1,1) | 1<br>(1,1) | 1<br>(1,1) |
| | 16-18 | 1<br>(1,1) | 1<br>(1,1) | 1<br>(1,1) |
| | 19-21 | 1<br>(1,1) | 1<br>(1,1) | 1<br>(1,1) |
| | 22-24 | 1<br>(1,1) | 1<br>(1,1) | 1<br>(1,1) |

**Supplemental Table 19. Median and interquartile range for duration of pathogen-attributable diarrhea by 3-month age bands (unstratified or stratified by infection history) using algorithm attribution.** Algorithm attribution considered both pathogen-attributable fraction and longitudinal changes in the quantity of pathogen detected. Because the recorded start and end of symptoms are consistent across infection definitions, they are not separately reported.

| Pathogen | Age (months) | Median duration in days (interquartile range) | | |
|---|---|---|---|---|
| | | Unstratified | Stratified by infection history | |
| | | | No prior infections | ≥1 prior infection |
| Adenovirus 40/41 | 1-3 | 2.0 (1.0,4.5) | 2.0 (1.0,4.0) | 8.0 (5.5,9.5) |
| | 4-6 | 3.0 (1.0,4.5) | 3.0 (1.0,4.5) | 2.5 (1.0,4.5) |
| | 7-9 | 2.0 (1.0,5.0) | 3.0 (1.0,4.5) | 2.0 (1.0,5.0) |
| | 10-12 | 2.0 (1.0,4.0) | 2.0 (1.0,4.0) | 2.0 (1.0,3.8) |
| | 13-15 | 2.0 (1.0,4.0) | 2.0 (1.0,4.8) | 2.0 (1.0,3.0) |
| | 16-18 | 2.0 (1.0,3.0) | 2.0 (1.0,3.0) | 2.0 (1.0,3.0) |
| | 19-21 | 2.0 (1.0,3.0) | 3.5 (2.8,5.2) | 2.0 (1.0,3.0) |
| | 22-24 | 1.0 (1.0,3.0) | 2.5 (1.2,5.2) | 1.0 (1.0,3.0) |
| Astrovirus | 1-3 | 3.0 (1.0,8.0) | 3.0 (1.2,8.0) | 1.5 (0.8,2.5) |
| | 4-6 | 2.0 (1.0,5.0) | 2.0 (1.0,5.0) | 3.5 (2.0,5.8) |
| | 7-9 | 3.0 (2.0,5.5) | 3.0 (2.0,6.0) | 3.0 (1.0,4.0) |
| | 10-12 | 3.0 (2.0,4.0) | 3.0 (2.0,4.2) | 3.0 (2.0,4.0) |
| | 13-15 | 2.0 (1.0,4.0) | 4.0 (2.0,5.0) | 2.0 (1.0,3.5) |
| | 16-18 | 3.0 (1.0,4.0) | 2.5 (1.0,3.0) | 3.0 (1.0,4.0) |
| | 19-21 | 2.0 (1.0,3.0) | 2.0 (0.5,3.0) | 2.0 (1.0,3.0) |
| | 22-24 | 2.0 (1.0,3.0) | 2.0 (1.0,3.0) | 2.0 (1.0,3.2) |
| *Campylobacter jejuni* and *coli* | 1-3 | 3.0 (2.0,6.0) | 3.0 (1.8,6.0) | 2.0 (2.0,2.0) |
| | 4-6 | 3.0 (1.0,5.0) | 3.0 (1.0,5.0) | 3.0 (1.2,5.0) |
| | 7-9 | 3.0 (2.0,6.0) | 3.0 (2.0,6.0) | 3.0 (2.0,5.0) |
| | 10-12 | 3.0 (2.0,5.0) | 3.0 (1.0,4.0) | 3.0 (2.0,5.0) |
| | 13-15 | 3.0 (1.0,4.0) | 3.0 (2.0,7.0) | 2.0 (1.0,4.0) |
| | 16-18 | 2.0 (1.0,4.0) | 2.0 (1.2,3.8) | 2.0 (1.0,4.0) |
| | 19-21 | 2.0 (1.0,5.0) | 1.5 (1.0,2.8) | 2.0 (1.0,5.0) |
| | 22-24 | 2.0 (1.0,3.0) | 2.0 (1.0,3.0) | 2.0 (1.0,3.0) |
| Cryptosporidium | 1-3 | 7.0 (4.5,14.0) | 7.0 (4.5,14.0) | -- |
| | 4-6 | 3.0 (2.0,5.5) | 3.0 (2.0,5.5) | -- |

| | | | | |
|---|---|---|---|---|
| | 7-9 | 3.0 (2.0,6.0) | 3.0 (2.0,7.5) | 2.0 (2.0,2.0) |
| | 10-12 | 3.0 (1.0,4.0) | 2.5 (1.0,4.2) | 3.0 (1.0,4.0) |
| | 13-15 | 2.0 (1.0,5.0) | 3.5 (1.2,6.0) | 2.0 (1.0,3.0) |
| | 16-18 | 3.0 (1.0,6.0) | 3.0 (1.0,5.5) | 4.5 (2.2,6.0) |
| | 19-21 | 2.0 (1.0,3.0) | 2.0 (1.0,3.0) | 2.0 (1.5,3.0) |
| | 22-24 | 2.0 (1.0,3.0) | 2.0 (1.0,3.0) | 2.0 (1.0,3.0) |
| Norovirus GII | 1-3 | 3.0 (2.0,5.0) | 3.0 (2.0,5.0) | 4.0 (3.5,4.5) |
| | 4-6 | 3.0 (1.0,6.0) | 3.0 (1.0,6.0) | 3.0 (1.0,4.5) |
| | 7-9 | 3.0 (1.0,5.0) | 2.0 (1.0,5.5) | 3.0 (2.0,5.0) |
| | 10-12 | 2.0 (1.0,4.0) | 2.0 (2.0,4.0) | 2.0 (1.0,3.5) |
| | 13-15 | 2.0 (1.0,5.0) | 2.0 (2.0,4.5) | 2.0 (1.0,5.0) |
| | 16-18 | 2.0 (1.0,4.0) | 3.5 (1.2,4.0) | 2.0 (1.0,3.0) |
| | 19-21 | 2.5 (1.0,4.2) | 4.0 (1.8,12.0) | 2.5 (1.0,4.0) |
| | 22-24 | 2.0 (1.0,3.5) | 2.0 (1.0,2.0) | 2.0 (1.0,3.8) |
| Rotavirus | 1-3 | 3.0 (1.0,5.0) | 3.0 (1.0,5.5) | 2.5 (1.8,3.2) |
| | 4-6 | 4.0 (2.0,6.0) | 4.0 (2.0,6.0) | 2.5 (1.2,4.5) |
| | 7-9 | 3.0 (2.0,5.0) | 3.0 (2.0,5.0) | 2.0 (1.0,4.0) |
| | 10-12 | 3.0 (2.0,4.0) | 3.0 (2.0,4.0) | 2.0 (1.0,3.0) |
| | 13-15 | 3.0 (1.0,4.0) | 3.0 (1.0,4.0) | 2.0 (1.0,4.0) |
| | 16-18 | 3.0 (1.0,5.0) | 3.0 (1.0,5.0) | 2.5 (1.0,3.2) |
| | 19-21 | 3.0 (1.0,3.0) | 3.0 (3.0,4.0) | 2.0 (1.0,3.0) |
| | 22-24 | 1.0 (0.0,3.0) | 1.0 (0.5,2.5) | 1.5 (0.2,2.8) |
| Sapovirus | 1-3 | 7.0 (3.0,14.0) | 8.0 (5.5,14.0) | 3.0 (2.0,15.5) |
| | 4-6 | 3.0 (1.0,6.0) | 3.0 (1.0,5.0) | 5.5 (3.5,6.0) |
| | 7-9 | 3.0 (2.0,5.0) | 3.0 (1.0,4.0) | 4.0 (2.0,5.0) |
| | 10-12 | 3.0 (1.0,4.0) | 3.0 (1.0,5.0) | 3.0 (1.2,4.0) |
| | 13-15 | 2.0 (1.0,4.0) | 2.0 (1.0,3.2) | 2.0 (1.0,4.0) |
| | 16-18 | 2.0 (1.0,4.0) | 2.0 (1.0,6.0) | 2.0 (1.0,4.0) |
| | 19-21 | 2.0 (1.0,4.0) | 2.0 (2.0,4.0) | 2.0 (1.0,4.0) |
| | 22-24 | 2.0 (1.0,3.0) | 2.0 (1.0,3.5) | 2.0 (1.0,3.0) |
| Shigella | 1-3 | 3.0 (1.2,7.8) | 4.0 (2.0,9.0) | 1.0 (1.0,1.0) |
| | 4-6 | 3.0 (1.0,7.0) | 3.0 (1.0,7.0) | 0.0 (0.0,0.0) |

| | | | | |
|---|---|---|---|---|
| | 7-9 | 3.0<br>(1.0,6.0) | 3.0<br>(2.0,6.0) | 1.0<br>(1.0,3.0) |
| | 10-12 | 2.0<br>(1.0,4.0) | 3.0<br>(1.0,5.0) | 2.0<br>(1.0,3.0) |
| | 13-15 | 2.0<br>(1.0,5.0) | 3.0<br>(2.0,5.0) | 2.0<br>(1.0,4.2) |
| | 16-18 | 3.0<br>(1.0,6.0) | 3.0<br>(1.0,6.0) | 3.0<br>(1.0,6.0) |
| | 19-21 | 2.0<br>(1.0,4.0) | 3.0<br>(1.0,4.2) | 2.0<br>(1.0,3.0) |
| | 22-24 | 2.0<br>(1.0,4.0) | 3.0<br>(2.0,4.8) | 2.0<br>(1.0,3.0) |
| ST-ETEC | 1-3 | 4.0<br>(1.0,6.0) | 4.0<br>(1.0,6.5) | 4.0<br>(3.0,5.0) |
| | 4-6 | 3.0<br>(1.0,4.8) | 3.0<br>(1.0,5.5) | 3.0<br>(1.0,4.0) |
| | 7-9 | 2.0<br>(1.0,4.0) | 3.0<br>(1.0,5.0) | 2.0<br>(1.0,4.0) |
| | 10-12 | 2.0<br>(1.0,4.0) | 3.0<br>(1.0,5.0) | 2.0<br>(1.0,3.0) |
| | 13-15 | 2.0<br>(1.0,4.5) | 3.0<br>(2.0,7.0) | 2.0<br>(1.0,4.0) |
| | 16-18 | 2.0<br>(1.0,4.0) | 2.5<br>(1.0,4.2) | 2.0<br>(1.0,4.0) |
| | 19-21 | 2.0<br>(1.0,4.0) | 2.0<br>(1.5,4.5) | 2.0<br>(1.0,4.0) |
| | 22-24 | 2.0<br>(1.0,4.0) | 3.0<br>(1.0,4.0) | 2.0<br>(1.0,3.2) |
| tEPEC | 1-3 | 3.0<br>(2.0,10.0) | 3.0<br>(2.0,10.5) | 4.0<br>(2.2,6.5) |
| | 4-6 | 3.0<br>(1.0,6.0) | 3.0<br>(1.0,5.5) | 3.0<br>(2.0,7.5) |
| | 7-9 | 2.0<br>(1.0,6.0) | 2.0<br>(1.0,6.8) | 3.0<br>(1.5,4.5) |
| | 10-12 | 2.0<br>(1.0,5.0) | 2.0<br>(1.0,5.0) | 2.0<br>(1.0,4.0) |
| | 13-15 | 3.0<br>(1.0,5.0) | 3.0<br>(1.8,5.5) | 3.0<br>(1.0,5.0) |
| | 16-18 | 3.0<br>(1.0,5.0) | 2.0<br>(2.0,3.0) | 3.0<br>(1.0,5.5) |
| | 19-21 | 3.0<br>(1.8,5.2) | 3.0<br>(2.0,3.5) | 4.0<br>(2.0,6.0) |
| | 22-24 | 2.0<br>(1.0,3.0) | 3.0<br>(2.0,3.5) | 2.0<br>(1.0,3.0) |

**Supplemental Table 20. Infection incidence rates per 100 infant-months and exact 95% confidence intervals (95% CIs) by 3-month age bands using Ct <35 to define a positive detection, algorithm attribution, and different infection definitions (crude or consecutive detections).** Algorithm attribution considered both pathogen-attributable fraction and longitudinal changes in the quantity of pathogen detected. With the crude infection definition, all positives were unique infections unless positive asymptomatic and symptomatic detections were separated by <7 days. The consecutive detections definition considered both consecutive positive detections and time between consecutive positives.

| Pathogen | Age (months) | Algorithm attribution | | | |
|---|---|---|---|---|---|
| | | Crude | | Consecutive detections | |
| | | Infections (Infant-months) | Incidence (95% CI) | Infections (Infant-months) | Incidence (95% CI) |
| Adenovirus 40/41 | 1-3 | 429 (4,185.7) | 10.25 (9.30,11.27) | 394 (3,822.1) | 10.31 (9.32,11.38) |
| | 4-6 | 723 (4,448.5) | 16.25 (15.09,17.48) | 586 (3,948.9) | 14.84 (13.66,16.09) |
| | 7-9 | 896 (4,516.9) | 19.84 (18.56,21.18) | 705 (3,951.8) | 17.84 (16.55,19.21) |
| | 10-12 | 860 (4,523.9) | 19.01 (17.76,20.32) | 673 (3,958.8) | 17.00 (15.74,18.33) |
| | 13-15 | 850 (4,535.3) | 18.74 (17.50,20.05) | 675 (3,987.8) | 16.93 (15.67,18.25) |
| | 16-18 | 762 (4,523.8) | 16.84 (15.67,18.08) | 621 (4,009.9) | 15.49 (14.29,16.75) |
| | 19-21 | 733 (4,492.7) | 16.32 (15.16,17.54) | 592 (3,983.5) | 14.86 (13.69,16.11) |
| | 22-24 | 678 (4,520.5) | 15.00 (13.89,16.17) | 544 (4,082.1) | 13.33 (12.23,14.49) |
| Astrovirus | 1-3 | 423 (4,179.0) | 10.12 (9.18,11.13) | 381 (3,847.2) | 9.90 (8.93,10.95) |
| | 4-6 | 645 (4,454.8) | 14.48 (13.38,15.64) | 523 (4,046.6) | 12.92 (11.84,14.08) |
| | 7-9 | 683 (4,561.1) | 14.97 (13.87,16.14) | 554 (4,124.6) | 13.43 (12.34,14.60) |
| | 10-12 | 758 (4,532.1) | 16.73 (15.56,17.96) | 592 (4,020.3) | 14.73 (13.56,15.96) |
| | 13-15 | 698 (4,576.9) | 15.25 (14.14,16.43) | 535 (4,170.5) | 12.83 (11.76,13.96) |
| | 16-18 | 632 (4,548.0) | 13.90 (12.83,15.02) | 526 (4,128.0) | 12.74 (11.68,13.88) |
| | 19-21 | 541 (4,527.1) | 11.95 (10.96,13.00) | 451 (4,174.2) | 10.80 (9.83,11.85) |
| | 22-24 | 508 (4,540.4) | 11.19 (10.24,12.20) | 436 (4,217.2) | 10.34 (9.39,11.36) |
| *Campylobacter jejuni* and *coli* | 1-3 | 294 (4,220.2) | 6.97 (6.19,7.81) | 256 (3,883.0) | 6.59 (5.81,7.45) |
| | 4-6 | 741 (4,441.8) | 16.68 (15.50,17.93) | 514 (3,838.4) | 13.39 (12.26,14.60) |
| | 7-9 | 1189 (4,470.3) | 26.60 (25.11,28.15) | 814 (3,499.9) | 23.26 (21.69,24.91) |
| | 10-12 | 1317 (4,432.8) | 29.71 (28.13,31.36) | 796 (3,524.6) | 22.58 (21.04,24.21) |
| | 13-15 | 1207 (4,477.5) | 26.96 (25.46,28.52) | 727 (3,688.5) | 19.71 (18.30,21.20) |
| | 16-18 | 1014 (4,466.8) | 22.70 (21.32,24.14) | 645 (3,796.8) | 16.99 (15.70,18.35) |
| | 19-21 | 913 (4,448.6) | 20.52 (19.21,21.90) | 577 (3,836.2) | 15.04 (13.84,16.32) |
| | 22-24 | 821 (4,479.3) | 18.33 (17.10,19.63) | 526 (4,057.4) | 12.96 (11.88,14.12) |
| *Cryptosporidium* | 1-3 | 56 (4,259.4) | 1.31 (0.99,1.71) | 54 (4,209.8) | 1.28 (0.96,1.67) |

| | | | | | |
|---|---|---|---|---|---|
| | 4-6 | 144 (4,559.2) | 3.16 (2.66,3.72) | 113 (4,449.0) | 2.54 (2.09,3.05) |
| | 7-9 | 293 (4,643.8) | 6.31 (5.61,7.07) | 218 (4,422.7) | 4.93 (4.30,5.63) |
| | 10-12 | 454 (4,598.1) | 9.87 (8.99,10.83) | 334 (4,276.0) | 7.81 (7.00,8.70) |
| | 13-15 | 566 (4,602.0) | 12.30 (11.31,13.36) | 400 (4,201.2) | 9.52 (8.61,10.50) |
| | 16-18 | 602 (4,547.5) | 13.24 (12.20,14.34) | 424 (4,120.6) | 10.29 (9.33,11.32) |
| | 19-21 | 578 (4,516.8) | 12.80 (11.77,13.88) | 409 (4,122.1) | 9.92 (8.98,10.93) |
| | 22-24 | 579 (4,526.0) | 12.79 (11.77,13.88) | 425 (4,158.4) | 10.22 (9.27,11.24) |
| Norovirus GII | 1-3 | 294 (4,217.6) | 6.97 (6.20,7.81) | 254 (3,921.8) | 6.48 (5.70,7.32) |
| | 4-6 | 817 (4,439.0) | 18.41 (17.16,19.71) | 594 (3,860.4) | 15.39 (14.17,16.68) |
| | 7-9 | 1060 (4,505.0) | 23.53 (22.13,24.99) | 785 (3,787.7) | 20.72 (19.30,22.23) |
| | 10-12 | 909 (4,518.6) | 20.12 (18.83,21.47) | 703 (3,933.6) | 17.87 (16.57,19.24) |
| | 13-15 | 724 (4,576.6) | 15.82 (14.69,17.01) | 599 (4,085.2) | 14.66 (13.51,15.89) |
| | 16-18 | 615 (4,562.3) | 13.48 (12.44,14.59) | 528 (4,141.2) | 12.75 (11.69,13.89) |
| | 19-21 | 470 (4,541.0) | 10.35 (9.44,11.33) | 414 (4,200.6) | 9.86 (8.93,10.85) |
| | 22-24 | 424 (4,555.8) | 9.31 (8.44,10.24) | 373 (4,275.0) | 8.73 (7.86,9.66) |
| Rotavirus | 1-3 | 236 (2,849.3) | 8.28 (7.26,9.41) | 218 (2,660.7) | 8.19 (7.14,9.36) |
| | 4-6 | 308 (3,033.8) | 10.15 (9.05,11.35) | 260 (2,849.0) | 9.13 (8.05,10.31) |
| | 7-9 | 334 (3,086.7) | 10.82 (9.69,12.05) | 280 (2,890.8) | 9.69 (8.58,10.89) |
| | 10-12 | 277 (3,111.2) | 8.90 (7.89,10.02) | 249 (2,944.5) | 8.46 (7.44,9.57) |
| | 13-15 | 222 (3,129.1) | 7.09 (6.19,8.09) | 200 (2,997.9) | 6.67 (5.78,7.66) |
| | 16-18 | 148 (3,119.3) | 4.74 (4.01,5.57) | 137 (3,024.4) | 4.53 (3.80,5.36) |
| | 19-21 | 146 (3,079.9) | 4.74 (4.00,5.57) | 139 (2,977.6) | 4.67 (3.92,5.51) |
| | 22-24 | 125 (3,047.0) | 4.10 (3.41,4.89) | 118 (2,960.7) | 3.99 (3.30,4.77) |
| Sapovirus | 1-3 | 225 (4,226.1) | 5.32 (4.65,6.07) | 208 (4,028.6) | 5.16 (4.49,5.91) |
| | 4-6 | 492 (4,497.2) | 10.94 (9.99,11.95) | 406 (4,142.8) | 9.80 (8.87,10.80) |
| | 7-9 | 859 (4,539.3) | 18.92 (17.68,20.23) | 669 (3,944.9) | 16.96 (15.70,18.29) |
| | 10-12 | 1008 (4,492.0) | 22.44 (21.08,23.87) | 735 (3,827.3) | 19.20 (17.84,20.64) |
| | 13-15 | 939 (4,537.8) | 20.69 (19.39,22.06) | 713 (3,920.7) | 18.19 (16.87,19.57) |
| | 16-18 | 895 (4,502.9) | 19.88 (18.60,21.22) | 718 (3,886.7) | 18.47 (17.15,19.88) |
| | 19-21 | 760 (4,496.0) | 16.90 (15.72,18.15) | 596 (3,980.8) | 14.97 (13.79,16.22) |
| | 22-24 | 678 (4,511.8) | 15.03 (13.92,16.20) | 559 (4,077.5) | 13.71 (12.60,14.89) |
| *Shigella* | 1-3 | 81 (4,256.0) | 1.90 (1.51,2.37) | 79 (4,184.4) | 1.89 (1.49,2.35) |

| | | | | | |
|---|---|---|---|---|---|
| | 4-6 | 136<br>(4,568.2) | 2.98<br>(2.50,3.52) | 123<br>(4,457.2) | 2.76<br>(2.29,3.29) |
| | 7-9 | 339<br>(4,628.6) | 7.32<br>(6.56,8.15) | 257<br>(4,362.3) | 5.89<br>(5.19,6.66) |
| | 10-12 | 595<br>(4,576.0) | 13.00<br>(11.98,14.09) | 414<br>(4,143.5) | 9.99<br>(9.05,11.00) |
| | 13-15 | 684<br>(4,580.4) | 14.93<br>(13.83,16.10) | 492<br>(4,109.9) | 11.97<br>(10.94,13.08) |
| | 16-18 | 823<br>(4,516.8) | 18.22<br>(17.00,19.51) | 589<br>(3,879.2) | 15.18<br>(13.98,16.46) |
| | 19-21 | 958<br>(4,458.2) | 21.49<br>(20.15,22.89) | 629<br>(3,774.3) | 16.67<br>(15.39,18.02) |
| | 22-24 | 968<br>(4,456.5) | 21.72<br>(20.37,23.13) | 632<br>(3,938.4) | 16.05<br>(14.82,17.35) |
| ST-ETEC | 1-3 | 226<br>(4,227.1) | 5.35<br>(4.67,6.09) | 210<br>(4,031.1) | 5.21<br>(4.53,5.96) |
| | 4-6 | 521<br>(4,485.4) | 11.62<br>(10.64,12.66) | 443<br>(4,108.1) | 10.78<br>(9.80,11.84) |
| | 7-9 | 755<br>(4,551.7) | 16.59<br>(15.43,17.81) | 582<br>(4,018.4) | 14.48<br>(13.33,15.71) |
| | 10-12 | 907<br>(4,508.5) | 20.12<br>(18.83,21.47) | 703<br>(3,862.1) | 18.20<br>(16.88,19.60) |
| | 13-15 | 961<br>(4,522.7) | 21.25<br>(19.93,22.64) | 700<br>(3,888.0) | 18.00<br>(16.69,19.39) |
| | 16-18 | 963<br>(4,482.0) | 21.49<br>(20.15,22.89) | 719<br>(3,807.3) | 18.88<br>(17.53,20.32) |
| | 19-21 | 944<br>(4,439.4) | 21.26<br>(19.93,22.67) | 674<br>(3,785.0) | 17.81<br>(16.49,19.20) |
| | 22-24 | 884<br>(4,464.2) | 19.80<br>(18.52,21.15) | 657<br>(3,916.5) | 16.78<br>(15.52,18.11) |
| tEPEC | 1-3 | 263<br>(4,215.0) | 6.24<br>(5.51,7.04) | 218<br>(3,985.1) | 5.47<br>(4.77,6.25) |
| | 4-6 | 580<br>(4,472.3) | 12.97<br>(11.93,14.07) | 453<br>(4,015.4) | 11.28<br>(10.27,12.37) |
| | 7-9 | 843<br>(4,543.6) | 18.55<br>(17.32,19.85) | 624<br>(3,962.4) | 15.75<br>(14.54,17.03) |
| | 10-12 | 810<br>(4,531.9) | 17.87<br>(16.66,19.15) | 593<br>(4,001.5) | 14.82<br>(13.65,16.06) |
| | 13-15 | 729<br>(4,573.0) | 15.94<br>(14.81,17.14) | 560<br>(4,102.0) | 13.65<br>(12.54,14.83) |
| | 16-18 | 636<br>(4,551.9) | 13.97<br>(12.91,15.10) | 531<br>(4,089.1) | 12.99<br>(11.90,14.14) |
| | 19-21 | 631<br>(4,511.9) | 13.99<br>(12.92,15.12) | 518<br>(4,069.3) | 12.73<br>(11.66,13.87) |
| | 22-24 | 570<br>(4,531.2) | 12.58<br>(11.57,13.66) | 463<br>(4,162.9) | 11.12<br>(10.13,12.18) |

**Supplemental Table 21. Proportion of infections that were pathogen-attributable diarrhea and exact 95% confidence intervals (95% CIs) by 6-month age bands using Ct <35 to define a positive detection, algorithm attribution, and different infection definitions (crude or consecutive detections).** Algorithm attribution considered both pathogen-attributable fraction and longitudinal changes in the quantity of pathogen detected. With the crude infection definition, all positives were unique infections unless positive asymptomatic and symptomatic detections were separated by <7 days. The consecutive detections definition considered both consecutive positive detections and time between consecutive positives.

| Pathogen | Age (months) | Algorithm attribution | | | |
|---|---|---|---|---|---|
| | | Crude | | Consecutive detections | |
| | | Diarrhea (infections) | Proportion (95% CI) | Diarrhea (infections) | Proportion (95% CI) |
| Adenovirus 40/41 | 1-6 | 142 (1,152) | 0.12 (0.10,0.14) | 142 (980) | 0.14 (0.12,0.17) |
| | 7-12 | 228 (1,756) | 0.13 (0.11,0.15) | 228 (1,378) | 0.17 (0.15,0.19) |
| | 13-18 | 164 (1,612) | 0.10 (0.09,0.12) | 164 (1,296) | 0.13 (0.11,0.15) |
| | 19-24 | 106 (1,411) | 0.08 (0.06,0.09) | 106 (1,136) | 0.09 (0.08,0.11) |
| Astrovirus | 1-6 | 131 (1,068) | 0.12 (0.10,0.14) | 131 (904) | 0.14 (0.12,0.17) |
| | 7-12 | 169 (1,441) | 0.12 (0.10,0.14) | 169 (1,146) | 0.15 (0.13,0.17) |
| | 13-18 | 155 (1,330) | 0.12 (0.10,0.14) | 155 (1,061) | 0.15 (0.13,0.17) |
| | 19-24 | 107 (1,049) | 0.10 (0.08,0.12) | 107 (887) | 0.12 (0.10,0.14) |
| *Campylobacter jejuni* and *coli* | 1-6 | 105 (1,035) | 0.10 (0.08,0.12) | 105 (770) | 0.14 (0.11,0.16) |
| | 7-12 | 204 (2,506) | 0.08 (0.07,0.09) | 204 (1,610) | 0.13 (0.11,0.14) |
| | 13-18 | 127 (2,221) | 0.06 (0.05,0.07) | 127 (1,372) | 0.09 (0.08,0.11) |
| | 19-24 | 66 (1,734) | 0.04 (0.03,0.05) | 66 (1,103) | 0.06 (0.05,0.08) |
| *Cryptosporidium* | 1-6 | 18 (200) | 0.09 (0.05,0.14) | 18 (167) | 0.11 (0.07,0.16) |
| | 7-12 | 66 (747) | 0.09 (0.07,0.11) | 66 (552) | 0.12 (0.09,0.15) |
| | 13-18 | 84 (1,168) | 0.07 (0.06,0.09) | 84 (824) | 0.10 (0.08,0.12) |
| | 19-24 | 77 (1,157) | 0.07 (0.05,0.08) | 77 (834) | 0.09 (0.07,0.11) |
| Norovirus GII | 1-6 | 106 (1,111) | 0.10 (0.08,0.11) | 106 (848) | 0.12 (0.10,0.15) |
| | 7-12 | 254 (1,969) | 0.13 (0.11,0.14) | 254 (1,488) | 0.17 (0.15,0.19) |
| | 13-18 | 152 (1,339) | 0.11 (0.10,0.13) | 152 (1,127) | 0.13 (0.12,0.16) |
| | 19-24 | 67 (894) | 0.07 (0.06,0.09) | 67 (787) | 0.09 (0.07,0.11) |
| Rotavirus | 1-6 | 109 (544) | 0.20 (0.17,0.24) | 109 (478) | 0.23 (0.19,0.27) |
| | 7-12 | 202 (611) | 0.33 (0.29,0.37) | 202 (529) | 0.38 (0.34,0.42) |
| | 13-18 | 111 (370) | 0.30 (0.25,0.35) | 111 (337) | 0.33 (0.28,0.38) |
| | 19-24 | 50 (271) | 0.18 (0.14,0.24) | 50 (257) | 0.19 (0.15,0.25) |
| Sapovirus | 1-6 | 79 (717) | 0.11 (0.09,0.14) | 79 (614) | 0.13 (0.10,0.16) |

| | | | | | |
|---|---|---|---|---|---|
| | 7-12 | 267 (1,867) | 0.14 (0.13,0.16) | 267 (1,404) | 0.19 (0.17,0.21) |
| | 13-18 | 266 (1,834) | 0.15 (0.13,0.16) | 266 (1,431) | 0.19 (0.17,0.21) |
| | 19-24 | 145 (1,438) | 0.10 (0.09,0.12) | 145 (1,155) | 0.13 (0.11,0.15) |
| *Shigella* | 1-6 | 27 (217) | 0.12 (0.08,0.18) | 27 (202) | 0.13 (0.09,0.19) |
| | 7-12 | 150 (934) | 0.16 (0.14,0.19) | 150 (671) | 0.22 (0.19,0.26) |
| | 13-18 | 284 (1,507) | 0.19 (0.17,0.21) | 284 (1,081) | 0.26 (0.24,0.29) |
| | 19-24 | 309 (1,926) | 0.16 (0.14,0.18) | 309 (1,261) | 0.25 (0.22,0.27) |
| ST-ETEC | 1-6 | 87 (747) | 0.12 (0.09,0.14) | 87 (653) | 0.13 (0.11,0.16) |
| | 7-12 | 215 (1,662) | 0.13 (0.11,0.15) | 215 (1,285) | 0.17 (0.15,0.19) |
| | 13-18 | 211 (1,924) | 0.11 (0.10,0.12) | 211 (1,419) | 0.15 (0.13,0.17) |
| | 19-24 | 158 (1,828) | 0.09 (0.07,0.10) | 158 (1,331) | 0.12 (0.10,0.14) |
| tEPEC | 1-6 | 98 (843) | 0.12 (0.10,0.14) | 98 (671) | 0.15 (0.12,0.18) |
| | 7-12 | 99 (1,653) | 0.06 (0.05,0.07) | 99 (1,217) | 0.08 (0.07,0.10) |
| | 13-18 | 58 (1,365) | 0.04 (0.03,0.05) | 58 (1,091) | 0.05 (0.04,0.07) |
| | 19-24 | 42 (1,201) | 0.03 (0.03,0.05) | 42 (981) | 0.04 (0.03,0.06) |

**Supplemental Table 22. Infection incidence rates per 100 infant-months and exact 95% confidence intervals (95% CIs) by country using Ct <35 to define a positive detection, algorithm attribution, and different infection definitions (crude or consecutive detections).** Algorithm attribution considered both pathogen-attributable fraction and longitudinal changes in the quantity of pathogen detected. With the crude infection definition, all positives were unique infections unless positive asymptomatic and symptomatic detections were separated by <7 days. The consecutive detections definition considered both consecutive positive detections and time between consecutive positives.

| Pathogen | Country | Algorithm attribution | | | |
|---|---|---|---|---|---|
| | | Crude | | Consecutive detections | |
| | | Infections (Infant-months) | Incidence (95% CI) | Infections (Infant-months) | Incidence (95% CI) |
| Adenovirus 40/41 | Bangladesh | 1,387 (4,459.7) | 31.10 (29.49,32.78) | 1,049 (3,633.7) | 28.87 (27.15,30.67) |
| | Brazil | 137 (2,887.2) | 4.75 (3.98,5.61) | 133 (2,775.2) | 4.79 (4.01,5.68) |
| | India | 941 (4,838.6) | 19.45 (18.22,20.73) | 754 (4,146.5) | 18.18 (16.91,19.53) |
| | Nepal | 354 (5,173.0) | 6.84 (6.15,7.59) | 301 (4,922.1) | 6.12 (5.44,6.85) |
| | Peru | 1,401 (4,223.0) | 33.18 (31.46,34.96) | 1,050 (3,372.3) | 31.14 (29.28,33.08) |
| | Pakistan | 843 (5,327.0) | 15.83 (14.77,16.93) | 739 (4,750.1) | 15.56 (14.46,16.72) |
| | South Africa | 503 (4,554.8) | 11.04 (10.10,12.05) | 446 (4,154.1) | 10.74 (9.76,11.78) |
| | Tanzania | 365 (4,284.0) | 8.52 (7.67,9.44) | 318 (3,990.9) | 7.97 (7.12,8.89) |
| Astrovirus | Bangladesh | 1,201 (4,486.2) | 26.77 (25.28,28.33) | 912 (3,746.5) | 24.34 (22.79,25.98) |
| | Brazil | 52 (2,893.0) | 1.80 (1.34,2.36) | 52 (2,852.8) | 1.82 (1.36,2.39) |
| | India | 634 (4,873.1) | 13.01 (12.02,14.06) | 518 (4,421.2) | 11.72 (10.73,12.77) |
| | Nepal | 311 (5,190.6) | 5.99 (5.34,6.70) | 261 (4,979.7) | 5.24 (4.62,5.92) |
| | Peru | 1,109 (4,306.6) | 25.75 (24.26,27.31) | 879 (3,675.4) | 23.92 (22.36,25.55) |
| | Pakistan | 964 (5,290.1) | 18.22 (17.09,19.41) | 809 (4,666.4) | 17.34 (16.16,18.57) |
| | South Africa | 334 (4,574.8) | 7.30 (6.54,8.13) | 303 (4,311.6) | 7.03 (6.26,7.87) |
| | Tanzania | 283 (4,304.9) | 6.57 (5.83,7.39) | 264 (4,074.9) | 6.48 (5.72,7.31) |
| *Campylobacter jejuni and coli* | Bangladesh | 1,627 (4,404.8) | 36.94 (35.16,38.78) | 935 (3,336.7) | 28.02 (26.25,29.88) |
| | Brazil | 130 (2,882.8) | 4.51 (3.77,5.35) | 114 (2,779.3) | 4.10 (3.38,4.93) |
| | India | 705 (4,867.5) | 14.48 (13.43,15.59) | 508 (4,344.9) | 11.69 (10.70,12.75) |
| | Nepal | 1,092 (5,038.0) | 21.68 (20.41,23.00) | 716 (4,259.8) | 16.81 (15.60,18.09) |
| | Peru | 1,018 (4,331.0) | 23.51 (22.08,24.99) | 738 (3,708.2) | 19.90 (18.49,21.39) |
| | Pakistan | 1,038 (5,306.6) | 19.56 (18.39,20.79) | 774 (4,596.8) | 16.84 (15.67,18.07) |
| | South Africa | 280 (4,575.7) | 6.12 (5.42,6.88) | 250 (4,352.9) | 5.74 (5.05,6.50) |
| | Tanzania | 1,606 (4,030.9) | 39.84 (37.92,41.84) | 820 (2,746.3) | 29.86 (27.85,31.97) |
| *Cryptosporidium* | Bangladesh | 480 (4,623.3) | 10.38 (9.47,11.35) | 329 (4,321.0) | 7.61 (6.81,8.48) |

| | | | | | |
|---|---|---|---|---|---|
| | Brazil | 40 (2,896.1) | 1.38 (0.99,1.88) | 39 (2,864.5) | 1.36 (0.97,1.86) |
| | India | 315 (4,933.7) | 6.38 (5.70,7.13) | 261 (4,700.0) | 5.55 (4.90,6.27) |
| | Nepal | 356 (5,182.7) | 6.87 (6.17,7.62) | 229 (4,923.3) | 4.65 (4.07,5.29) |
| | Peru | 723 (4,387.7) | 16.48 (15.30,17.72) | 453 (3,943.8) | 11.49 (10.45,12.59) |
| | Pakistan | 482 (5,412.3) | 8.91 (8.13,9.74) | 379 (5,089.3) | 7.45 (6.72,8.24) |
| | South Africa | 254 (4,582.3) | 5.54 (4.88,6.27) | 219 (4,380.7) | 5.00 (4.36,5.71) |
| | Tanzania | 622 (4,234.5) | 14.69 (13.56,15.89) | 468 (3,737.1) | 12.52 (11.41,13.71) |
| Norovirus GII | Bangladesh | 812 (4,563.3) | 17.79 (16.59,19.06) | 642 (4,032.4) | 15.92 (14.71,17.20) |
| | Brazil | 139 (2,881.1) | 4.82 (4.06,5.70) | 129 (2,771.3) | 4.65 (3.89,5.53) |
| | India | 682 (4,872.8) | 14.00 (12.97,15.09) | 567 (4,368.1) | 12.98 (11.93,14.09) |
| | Nepal | 632 (5,126.6) | 12.33 (11.39,13.33) | 500 (4,686.2) | 10.67 (9.75,11.65) |
| | Peru | 890 (4,364.2) | 20.39 (19.08,21.78) | 676 (3,822.8) | 17.68 (16.38,19.07) |
| | Pakistan | 1,156 (5,278.8) | 21.90 (20.65,23.20) | 865 (4,495.1) | 19.24 (17.98,20.57) |
| | South Africa | 421 (4,573.1) | 9.21 (8.35,10.13) | 372 (4,237.6) | 8.78 (7.91,9.72) |
| | Tanzania | 581 (4,256.0) | 13.65 (12.56,14.81) | 499 (3,791.8) | 13.16 (12.03,14.37) |
| Rotavirus | Bangladesh | 594 (4,603.4) | 12.90 (11.89,13.98) | 524 (4,284.7) | 12.23 (11.20,13.32) |
| | India | 434 (4,900.7) | 8.86 (8.04,9.73) | 390 (4,579.1) | 8.52 (7.69,9.41) |
| | Nepal | 259 (5,199.7) | 4.98 (4.39,5.63) | 214 (5,041.2) | 4.25 (3.70,4.85) |
| | Pakistan | 282 (5,448.9) | 5.18 (4.59,5.82) | 257 (5,278.0) | 4.87 (4.29,5.50) |
| | Tanzania | 227 (4,303.6) | 5.27 (4.61,6.01) | 216 (4,122.7) | 5.24 (4.56,5.99) |
| Sapovirus | Bangladesh | 1,111 (4,506.1) | 24.66 (23.23,26.15) | 841 (3,805.7) | 22.10 (20.63,23.64) |
| | Brazil | 133 (2,883.1) | 4.61 (3.86,5.47) | 115 (2,777.6) | 4.14 (3.42,4.97) |
| | India | 818 (4,857.3) | 16.84 (15.71,18.04) | 639 (4,271.4) | 14.96 (13.82,16.17) |
| | Nepal | 680 (5,116.8) | 13.29 (12.31,14.33) | 487 (4,656.0) | 10.46 (9.55,11.43) |
| | Peru | 1,003 (4,343.9) | 23.09 (21.68,24.56) | 762 (3,744.3) | 20.35 (18.93,21.85) |
| | Pakistan | 1,111 (5,277.0) | 21.05 (19.83,22.33) | 867 (4,538.1) | 19.10 (17.85,20.42) |
| | South Africa | 518 (4,551.9) | 11.38 (10.42,12.40) | 455 (4,137.0) | 11.00 (10.01,12.06) |
| | Tanzania | 482 (4,266.9) | 11.30 (10.31,12.35) | 438 (3,879.2) | 11.29 (10.26,12.40) |
| *Shigella* | Bangladesh | 924 (4,545.6) | 20.33 (19.04,21.68) | 630 (3,963.1) | 15.90 (14.68,17.19) |
| | Brazil | 160 (2,873.6) | 5.57 (4.74,6.50) | 134 (2,747.8) | 4.88 (4.09,5.78) |
| | India | 718 (4,869.1) | 14.75 (13.69,15.87) | 488 (4,340.5) | 11.24 (10.27,12.29) |
| | Nepal | 390 (5,170.0) | 7.54 (6.81,8.33) | 244 (4,905.6) | 4.97 (4.37,5.64) |

| | | | | | |
|---|---|---|---|---|---|
| | Peru | 807<br>(4,377.9) | 18.43<br>(17.18,19.75) | 521<br>(3,872.0) | 13.46<br>(12.32,14.66) |
| | Pakistan | 455<br>(5,408.1) | 8.41<br>(7.66,9.22) | 395<br>(5,124.0) | 7.71<br>(6.97,8.51) |
| | South Africa | 339<br>(4,579.3) | 7.40<br>(6.64,8.23) | 284<br>(4,310.7) | 6.59<br>(5.84,7.40) |
| | Tanzania | 791<br>(4,217.3) | 18.76<br>(17.47,20.11) | 519<br>(3,585.4) | 14.48<br>(13.26,15.78) |
| ST-ETEC | Bangladesh | 1,903<br>(4,355.6) | 43.69<br>(41.75,45.70) | 1,244<br>(3,132.4) | 39.71<br>(37.54,41.98) |
| | Brazil | 73<br>(2,890.0) | 2.53<br>(1.98,3.18) | 72<br>(2,830.5) | 2.54<br>(1.99,3.20) |
| | India | 728<br>(4,852.4) | 15.00<br>(13.93,16.13) | 600<br>(4,322.6) | 13.88<br>(12.79,15.04) |
| | Nepal | 631<br>(5,126.1) | 12.31<br>(11.37,13.31) | 520<br>(4,681.1) | 11.11<br>(10.17,12.11) |
| | Peru | 759<br>(4,382.2) | 17.32<br>(16.11,18.60) | 637<br>(3,928.2) | 16.22<br>(14.98,17.53) |
| | Pakistan | 624<br>(5,379.7) | 11.60<br>(10.71,12.55) | 541<br>(4,976.2) | 10.87<br>(9.97,11.83) |
| | South Africa | 194<br>(4,580.2) | 4.24<br>(3.66,4.88) | 183<br>(4,427.8) | 4.13<br>(3.56,4.78) |
| | Tanzania | 1,249<br>(4,114.8) | 30.35<br>(28.69,32.09) | 891<br>(3,117.7) | 28.58<br>(26.73,30.52) |
| tEPEC | Bangladesh | 1,114<br>(4,496.4) | 24.78<br>(23.34,26.27) | 815<br>(3,766.0) | 21.64<br>(20.18,23.18) |
| | Brazil | 101<br>(2,892.2) | 3.49<br>(2.84,4.24) | 96<br>(2,813.9) | 3.41<br>(2.76,4.17) |
| | India | 883<br>(4,837.3) | 18.25<br>(17.07,19.50) | 684<br>(4,172.1) | 16.39<br>(15.19,17.67) |
| | Nepal | 412<br>(5,170.3) | 7.97<br>(7.22,8.78) | 324<br>(4,870.6) | 6.65<br>(5.95,7.42) |
| | Peru | 803<br>(4,368.1) | 18.38<br>(17.13,19.70) | 634<br>(3,874.6) | 16.36<br>(15.11,17.69) |
| | Pakistan | 714<br>(5,361.2) | 13.32<br>(12.36,14.33) | 593<br>(4,906.8) | 12.09<br>(11.13,13.10) |
| | South Africa | 213<br>(4,592.4) | 4.64<br>(4.04,5.30) | 197<br>(4,427.1) | 4.45<br>(3.85,5.12) |
| | Tanzania | 822<br>(4,213.0) | 19.51<br>(18.20,20.89) | 617<br>(3,556.5) | 17.35<br>(16.01,18.77) |

**Supplemental Table 23. Median and interquartile range for age at first infection, regardless of symptomology, overall and by country using Ct <35 to define a positive detection.** Because only the first positive detection mattered, results are identical regardless of infection definition Algorithm attribution considered both pathogen-attributable fraction (Afe) and longitudinal changes in the quantity of pathogen detected. With the crude infection definition, all positives were unique infections unless positive asymptomatic and symptomatic detections were separated by <7 days. The consecutive detections definition considered both consecutive positive detections and time between consecutive positives. Brazil, Peru, and South Africa had already introduced rotavirus vaccines and were not included in rotavirus analyses.

| Pathogen | Country | Age (days) at first infection | |
|---|---|---|---|
| | | Crude | Consecutive detections |
| Adenovirus 40/41 | All | 210.0 (119.0,336.0) | 210.0 (119.0,336.0) |
| | Bangladesh | 140.0 (88.0,198.0) | 140.0 (88.0,198.0) |
| | Brazil | 305.0 (162.0,488.0) | 305.0 (162.0,488.0) |
| | India | 151.0 (91.0,273.0) | 151.0 (91.0,273.0) |
| | Nepal | 334.0 (212.0,489.0) | 334.0 (212.0,489.0) |
| | Peru | 149.0 (89.0,213.8) | 149.0 (89.0,213.8) |
| | Pakistan | 212.5 (123.8,333.0) | 212.5 (123.0,333.0) |
| | South Africa | 272.0 (181.0,424.5) | 272.0 (181.0,424.5) |
| | Tanzania | 259.5 (151.0,423.2) | 259.5 (151.0,423.2) |
| Astrovirus | All | 191.5 (100.0,355.8) | 191.5 (100.0,355.8) |
| | Bangladesh | 149.0 (91.0,214.8) | 149.0 (91.0,214.8) |
| | Brazil | 460.0 (280.8,609.8) | 460.0 (280.8,609.8) |
| | India | 183.0 (92.5,304.5) | 183.0 (92.0,304.5) |
| | Nepal | 389.5 (272.2,498.8) | 389.5 (272.2,497.2) |
| | Peru | 153.0 (90.0,241.0) | 153.0 (90.0,239.5) |
| | Pakistan | 152.0 (79.0,242.0) | 152.0 (79.0,242.0) |
| | South Africa | 336.0 (185.0,485.0) | 336.0 (185.0,485.0) |
| | Tanzania | 212.0 (122.0,364.8) | 212.0 (122.0,364.8) |
| *Campylobacter jejuni* and *coli* | All | 215.0 (148.0,305.0) | 215.0 (148.0,305.0) |
| | Bangladesh | 211.0 (121.2,303.0) | 211.0 (121.2,303.0) |
| | Brazil | 309.0 (230.5,489.0) | 309.0 (230.5,489.0) |
| | India | 219.0 (151.0,333.8) | 219.0 (151.0,333.8) |
| | Nepal | 243.0 (180.0,334.0) | 243.0 (180.0,334.0) |
| | Peru | 237.0 (146.5,305.5) | 237.0 (146.5,305.5) |
| | Pakistan | 216.0 (149.0,303.0) | 216.0 (149.0,303.0) |

| | | | |
|---|---|---|---|
| | South Africa | 226.5 (124.2,335.0) | 226.5 (124.2,335.0) |
| | Tanzania | 180.0 (96.0,242.0) | 180.0 (96.0,242.0) |
| *Cryptosporidium* | All | 397.5 (274.8,532.8) | 397.0 (274.8,532.8) |
| | Bangladesh | 416.0 (275.0,538.0) | 416.0 (275.0,538.0) |
| | Brazil | 545.0 (399.0,677.0) | 545.0 (399.0,677.0) |
| | India | 402.0 (305.5,521.0) | 399.0 (305.5,521.0) |
| | Nepal | 546.0 (416.5,668.0) | 546.0 (416.5,668.0) |
| | Peru | 369.5 (268.0,487.8) | 369.5 (268.0,487.8) |
| | Pakistan | 364.0 (241.0,494.0) | 364.0 (241.0,494.0) |
| | South Africa | 425.0 (303.0,545.0) | 425.0 (303.0,545.0) |
| | Tanzania | 333.0 (243.0,429.8) | 333.0 (243.0,429.8) |
| Norovirus GII | All | 211.0 (127.0,303.0) | 211.0 (127.0,303.0) |
| | Bangladesh | 211.0 (151.0,272.0) | 211.0 (151.0,272.0) |
| | Brazil | 278.0 (211.5,424.0) | 278.0 (211.5,424.0) |
| | India | 212.0 (150.0,287.0) | 212.0 (150.0,287.0) |
| | Nepal | 245.0 (181.2,372.2) | 245.0 (181.2,372.2) |
| | Peru | 213.0 (152.0,274.0) | 212.5 (152.0,274.0) |
| | Pakistan | 152.0 (90.0,223.5) | 152.0 (90.0,223.5) |
| | South Africa | 244.5 (150.0,366.5) | 244.5 (150.0,366.5) |
| | Tanzania | 153.0 (118.0,240.8) | 153.0 (118.0,240.8) |
| Rotavirus | All | 213.0 (119.0,364.0) | 213.0 (119.0,364.0) |
| | Bangladesh | 149.5 (89.8,252.8) | 149.5 (89.8,252.8) |
| | India | 181.5 (89.8,318.5) | 181.5 (89.8,318.5) |
| | Nepal | 331.5 (213.2,474.0) | 331.5 (213.2,474.0) |
| | Pakistan | 247.5 (148.2,414.0) | 247.5 (148.2,414.0) |
| | Tanzania | 217.0 (152.0,365.0) | 217.0 (152.0,365.0) |
| Sapovirus | All | 246.0 (170.5,365.0) | 246.0 (170.5,365.0) |
| | Bangladesh | 211.0 (127.0,275.0) | 211.0 (127.0,275.0) |
| | Brazil | 427.0 (279.5,584.2) | 427.0 (279.5,584.2) |
| | India | 241.0 (155.0,307.0) | 241.0 (155.0,307.0) |
| | Nepal | 333.0 (226.5,437.0) | 333.0 (226.5,437.0) |
| | Peru | 251.0 (179.0,334.0) | 251.0 (179.0,334.0) |

| | | | |
|---|---|---|---|
| | Pakistan | 212.0 (149.0,309.0) | 212.0 (149.0,309.0) |
| | South Africa | 303.0 (181.0,457.0) | 303.0 (181.0,457.0) |
| | Tanzania | 246.5 (179.0,388.0) | 246.5 (179.0,388.0) |
| *Shigella* | All | 394.0 (273.0,518.0) | 394.0 (273.0,518.0) |
| | Bangladesh | 388.0 (279.5,495.0) | 388.0 (279.5,495.0) |
| | Brazil | 456.0 (310.0,548.0) | 456.0 (310.0,548.0) |
| | India | 335.0 (243.0,502.0) | 335.0 (243.0,502.0) |
| | Nepal | 481.5 (349.8,610.2) | 481.5 (349.8,610.2) |
| | Peru | 421.0 (304.0,517.0) | 421.0 (304.0,517.0) |
| | Pakistan | 398.0 (303.0,545.5) | 398.0 (303.0,545.5) |
| | South Africa | 369.0 (275.0,545.0) | 369.0 (275.0,545.0) |
| | Tanzania | 304.0 (216.5,396.5) | 304.0 (216.5,396.5) |
| ST-ETEC | All | 251.0 (153.0,395.0) | 251.0 (153.0,395.0) |
| | Bangladesh | 153.0 (90.0,220.0) | 153.0 (90.0,220.0) |
| | Brazil | 398.0 (188.2,578.0) | 398.0 (188.2,578.0) |
| | India | 271.0 (209.5,364.5) | 271.0 (209.5,364.5) |
| | Nepal | 304.5 (210.0,427.0) | 304.5 (210.0,427.0) |
| | Peru | 283.0 (159.0,387.0) | 283.0 (159.0,387.0) |
| | Pakistan | 301.5 (151.0,454.8) | 301.5 (151.0,454.8) |
| | South Africa | 425.0 (274.0,550.2) | 425.0 (274.0,550.2) |
| | Tanzania | 211.0 (123.0,275.2) | 211.0 (123.0,275.2) |
| tEPEC | All | 241.5 (152.0,341.5) | 241.5 (152.0,341.5) |
| | Bangladesh | 212.0 (152.0,275.0) | 212.0 (152.0,275.0) |
| | Brazil | 370.0 (245.0,538.0) | 370.0 (245.0,538.0) |
| | India | 212.0 (122.0,274.8) | 212.0 (122.0,274.8) |
| | Nepal | 334.5 (265.8,490.2) | 334.5 (265.8,490.2) |
| | Peru | 271.0 (181.0,340.0) | 271.0 (181.0,340.0) |
| | Pakistan | 181.0 (106.8,276.0) | 181.0 (106.8,276.0) |
| | South Africa | 318.5 (185.8,453.2) | 318.5 (185.8,453.2) |
| | Tanzania | 181.0 (124.0,242.0) | 181.0 (124.0,242.0) |
